# Ensemble Approaches for Robust and Generalizable Short-Term Forecasts of Dengue Fever. A retrospective and prospective evaluation study in over 180 locations around the world

**DOI:** 10.1101/2024.10.22.24315925

**Authors:** Skyler Wu, Austin Meyer, Leonardo Clemente, Lucas M. Stolerman, Fred Lu, Atreyee Majumder, Rudi Verbeeck, Serge Masyn, Mauricio Santillana

**Affiliations:** Department of Statistics, Harvard University, Cambridge, MA, USA; Machine Intelligence Group for the Betterment of Health and the Environment, Network Science Institute, Northeastern University, Boston, MA, USA; Beth Israel Deaconess Medical Center, Boston, MA, USA; Department of Mathematics, Oklahoma State University, Stillwater, OK, USA; Global Public Health, Janssen R&D, Beerse, Belgium; Harvard T.H. Chan School of Public Health, Boston, MA USA

## Abstract

Dengue fever, a tropical vector-borne disease, is a leading cause of hospitalization and death in many parts of the world, especially in Asia and Latin America. In places where timely and accurate dengue activity surveillance is available, decision-makers possess valuable information that may allow them to better design and implement public health measures, and improve the allocation of limited public health resources. In addition, robust and reliable near-term forecasts of likely epidemic outcomes may further help anticipate increased demand on healthcare infrastructure and may promote a culture of preparedness. Here, we propose ensemble modeling approaches that combine forecasts produced with a variety of independent mechanistic, statistical, and machine learning component models to forecast reported dengue case counts 1-, 2-, and 3-months ahead of current time at the province level in multiple countries. We assess the ensemble and each component models’ monthly predictive ability in a fully out-of-sample and retrospective fashion, in over 180 locations around the world — all provinces of Brazil, Colombia, Malaysia, Mexico, and Thailand, as well as Iquitos, Peru, and San Juan, Puerto Rico — during at least 2-3 years. Additionally, we evaluate ensemble approaches in a multi-model, real-time, and prospective dengue forecasting platform — where issues of data availability and data completeness introduce important limitations — during an 11-month time period in the years 2022 and 2023. We show that our ensemble modeling approaches lead to reliable and robust prediction estimates when compared to baseline estimates produced with available information at the time of prediction. This can be contrasted with the high variability in the forecasting ability of each individual component model, across locations and time. Furthermore, we find that no individual model leads to optimal and robust predictions across time horizons and locations, and while the ensemble models do not always achieve the best prediction performance in any given location, they consistently provide reliable disease estimates — they rank in the top 3 performing models across locations and time periods — both retrospectively and prospectively.

## 1 Introduction

Dengue fever is a tropical vector-borne disease threatening an estimated 3.9 billion people [62] over 141 countries [2], with cases doubling every ten years since 1990 [26]. Over 390 million infections arise each year worldwide, with severe dengue causing 25,000 deaths annually, mostly in children [2, 3]. For many parts of Asia and Latin America, dengue is a leading cause of hospitalization and death, especially among children [16]. It also causes more morbidity and mortality than any other arthropod-borne virus [9]. While dengue infections often presents with flu-like or otherwise mild symptoms [1], about 1 in 20 people infected with dengue will develop *severe dengue*, which can further develop into *dengue hemorrhagic fever* [3] — a very serious condition marked by capillary leakage leading to potentially significant organ impairment, multiorgan failure, and death [1, 3, 21, 28]. The ability to forecast dengue fever cases can provide public health officials with a more accurate picture of future disease dynamics, empowering decision-makers to implement public health measures and better allocate limited resources.

Over the past few decades, multiple approaches have been developed for dengue forecasting. Dynamic, mathematical models that incorporate knowledge of dengue virus transmission biology, historical incidence, and climatological factors have been developed to predict the evolution of dengue epidemics [11, 64, 20, 51]. However, the intricate nature of dengue dynamics poses a significant challenge, as mechanistic assumptions usually remain unclear — or hard to quantify —, and acquiring the data to parameterize models often proves impossible. More recently, data-driven methodologies to predict the severity of an upcoming seasonal outbreak — a classification problem — have experienced a surge in popularity due to the increasing availability of epidemiological and exogenous dengue-related data. These include methods such as *k*-Nearest-Neighbors, Logistic Regression, and various boosting methods [44]; or the identification of weather (temperature, rain frequency) patterns that may help anticipate years with high incidence [35, 57, 56]. Additionally, classical time series methods like SARIMA (and its variants) have been widely explored to forecast confirmed case counts over time[6, 18, 46, 59], as well as more complex, non-linear methods such as generalized additive models, artificial neural networks, and exponential smoothing approaches [5, 6, 29, 46]. Other studies have explored the feasibility of leveraging Internet-based data sources such as Dengue-related Google search data [27, 50, 62], social media data [17, 33], and Wikipedia access logs [33] as additional predictors of Dengue activity.

The abundance of dengue forecasting methodologies presents a significant challenge for decisionmakers and stakeholders who ultimately need a single set of reliable predictions to make informed decisions to protect their communities. Factors such as data availability, computational resources, and the desired level of accuracy further complicate the decision-making process.

Therefore, navigating the array of forecasting methodologies requires careful consideration and expertise to ensure effective dengue prediction and response. Additionally, each model has its own limitations [18, 43], including robustness to variability in data quality and sensitivity to different outbreak phases, which can lead to inconsistent predictions.

Ensemble methods that intelligently and adaptively combine the predictions of multiple component models into a single prediction may be more robust alternatives for disease forecasting. For example, previous studies have used averaged and weighted-averaged ensembles combining models such as SARIMA, vector autoregression, neural networks, and linear regression to nowcast and forecast dengue in Brazil and India [23, 53]. Similar ensembling approaches combining models such as the Method of Analogues, Holt-Winter models, and Bayesian generalized linear mixed models, among other historical models, have shown promising results in Iquitos, Peru [8], and Vietnam [12]. Chakraborty et al. directly combined ARIMA and neural networks to forecast dengue in San Juan, Iquitos, and the Philippines as a composition of linear and non-linear signals [10]. Mahajan et al. use a gradient-boosting super-ensemble to combine ARIMA, exponential smoothing, and neural network for forecasting dengue in Hong Kong [32]. Ensemble models intentionally involving strong and weak learners to reduce over-fitting have also shown good predictive performance in Bangkok and Chiang Mai, Thailand [24].

However, the studies referenced above are (1) individually limited in their geographic scope –to study a few locations within a country at a time–, (2) they only focus on a few model choices as potential ensemble components, and (3) they were in all cases implemented retrospectively. The latter implies that issues of data availability, data completeness, and other challenges that emerge in real-time forecasting efforts were not at all considered [34]. As such, the approaches described are not guaranteed to generalize as robust, ready-to-use methodologies globally, in real-time and prospective forecasting efforts.

We would like to highlight that the challenges of the real-time implementation of disease forecasting systems have been documented extensively, and they are still the backbone that motivate multiple research studies ([37, 31, 54]). In fact, public health systems typically experience severe lags in reporting [13, 36], and reported case counts for a given month may be updated many times in the following months (this is commonly referred to as “backfill”) [50]. While some methods for addressing these data quality and backfill issues have been proposed [15], for example — methods that attempt to learn backfill patterns [22, 37] and methods that introduce auxiliary data sources [45] to improve forecasts; we did not include in our real-time prediction pipeline a comprehensive set of approaches to address these issues. Instead, we evaluated the ability of our machine learning-based ensemble methods to lead to improved or consistent forecasts in the presence of these challenges.

The primary prediction task addressed by this manuscript is the short-term forecast of reported dengue fever case counts one to three months ahead in province-sized localities. The main contributions of this work are twofold. First, we retrospectively formulate a family of ensemble system pipelines — comprised of multiple structurally heterogeneous individual component models — that generate more robust and accurate forecasts compared to their individual component models. Specifically, we include the following 11 diverse classes of individual component models as potential inputs to our ensembling pipelines: autoregressive (AR); autoregressive with Google Trends data as exogenous covariates (ARGO [63]); three variants of vector autoregression (NetModel and two variants of VAR [41] — VAR (Reg.) and VAR (Clust., Reg.); a combination of ARGO and NetModel (ARGONet [31]); a novel, mechanistic, repurposed dynamically-trained SIR; a classical error-trend-seasonality model (ETS); a mini-ensemble of machine learning methods (Stacked ML); a baseline naive persistence model; and a baseline seasonal model. We exhaustively demonstrate across 180+ province-sized locations that our ensemble models not only incur lower percent absolute errors on average compared to their individual component models (though such improvements may be modest), but also, and more importantly, they produce top performing forecasts (typically in the top three) more consistently than any individual model.

The second contribution of this study is the evaluation of a real-time dengue activity forecasting platform implemented as a prospective tool to identify when and where an upcoming dengue outbreak would be experienced 1, 2 and 3 months into the future. These predictive efforts were implemented as a decision-making support tool to guide the allocation of resources for prospective clinical trials in all provinces of Brazil, Colombia, Malaysia, Mexico, and Thailand. The ensemble techniques were assessed using a different set of component models that were chosen based on their suitability to be implemented in the presence of multiple data availability and data incompleteness issues. These models included: KNN, VAR (Reg.), Support Vector Machines (SVM), and SARIMA. The scope of our study and the consistency of our analyzes suggests that our ensemble approaches are generalizable across geographically diverse locations and individual component models, and thus may become a reliable first choice for forecasting teams who are interested in communicating concisely with public health officials and other decision makers[54].

## Results

We evaluated the performances of eleven optimized component models and two selected ensemble models on forecasting reported dengue cases in over 180 province-sized locations worldwide in Brazil, Colombia, Malaysia, Mexico, and Thailand. Specifically, we retrospectively assessed each model’s ability to forecast reported dengue cases 1-, 2-, and 3-months ahead into the future, with performance quantified using Percent Absolute Error (see Supplementary Materials).

For each location, we partitioned the available data into distinct training and test periods. During the training phase, we engaged in a hyperparameter optimization for each component model, and two versions of optimal hyperparameters for our ensembles: a ‘Country’ set of hyperparameters (using the same set of hyperparameter with best on-average performance across all locations within a country), and a ‘Overall’ (using the set of hyperparameters that, on average, best performed within all the locations in the study). Following this stage, we proceeded to generate out-of-sample forecasts utilizing the designated test period. It is important to note that due to the varied availability of epidemiological data across different regions, the time periods for analysis varied from country to country. For specific details on the analysis period for each country, please consult Table 2.

We present our main results in the following way: First, we observe the heterogeneity in the performance of each of our component models in Figure 1, which presents a summary of the performance of each model across each country, and each horizon of prediction. Then, in Figure 2 we focus specifically on the capacity of the ensemble models to consistently succeed in generating reliable forecasts, independent of the location where they are trained on. Finally, we present an overview of the error reduction of each component model, emphasizing the consistency of the ensemble techniques to reduce error across locations.

**Figure 1:**
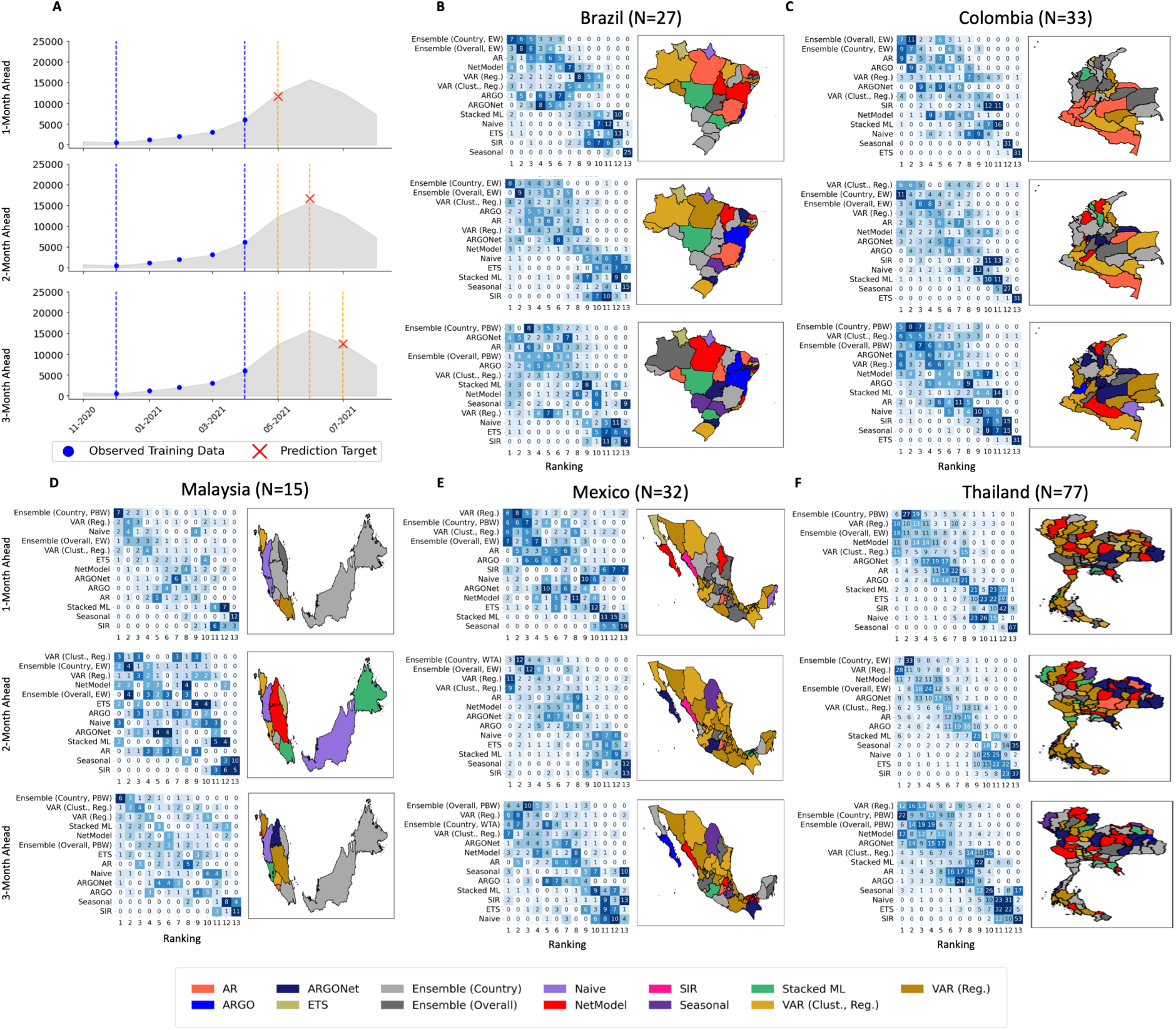
Country-specific optimized individual and ensemble model performance rankings. (A) Graphical representations of the 1-, 2-, and 3-month forecast horizons that we explore in this paper. The red *X* marks our forecasting target *n*-months ahead, the blue dots represent the historical cases that we are using as our observed training data (in this case, a 5-month sliding window), the vertical blue dotted lines represent the limits of our training data range, and the grey silhouette represents the ground truth reported case counts. (B) - (F) Within each country and forecast horizon, the heatmaps show the rankings distribution for each individual and ensemble model’s forecasts in terms of percent absolute error. The geographic maps next to each heatmap indicate the best-performing model in each province, color-coded by the legend at the bottom of the figure.

Component model performance were significantly dependent on the location. Our results, summarized in Figure 1, present a table with the percent absolute error ranking (PAE) distribution for each model within Sections B through F (one for each country). Each row within the table represents a distinct model, while columns denote their respective rankings (1st, 2nd, 3rd, etc.). For instance, for Brazil, the first entry of the table indicates that the model ‘Ensemble (Country, EW)’ secured the lowest PAE, achieving first place in 7 out of 27 locations across the country. The tables showcase the top-performing model in terms of this ranking, listed in descending order. We found that our ensembles, including the ‘Country’ and ‘Optimized’ versions of the ‘Equal Weights’ (EW) ensemble, and ‘Country’ and ‘Overall’ ‘Performance Based Weights’ (PBW) tended to be in the top positions within our ranking tables across all horizons for Brazil, Colombia, Mexico, and Thailand (see the Methods section for details on the ensemble approaches). In the case of Malaysia, the EW ensemble appeared in the 6*^th^* position (almost half of the participating models). The Vector Autoregression (VAR) also consistently appeared in top positions for each of the ranking matrices for each country. On the other hand, the positioning of each of our component models varied from country to country. A country-by-country description, along with additional results for San Juan, Puerto Rico, and Iquitos, Peru can be found in the Supplementary Materials. We relegate Puerto Rico and Peru to the Supplementary Materials, since we only have one location in each of these regions, which does not facilitate interprovince analyses.

### Ensemble models consistently achieve the most top performance rankings compared to any individual model

Our second main result is that the country-specific and overall ensembles consistently perform among the top 3 relative to the individual component models. Moreover, averaged across all locations, two ensemble variants incurred the lowest prediction error compared to all other component models. Fig. 2 shows the forecasting performance of both individual and ensemble models across all 187 tested locations. In panel (A), we provide one example emphasizing the advantages of using ensembles over component models. For the 1- and 2-month ahead horizons, the best country-specific ensemble used equal weights (EW) for all component models when producing an ensemble forecast at each timestep, while at the 3-month ahead horizon, the best country-specific ensemble used performance-based weights (PBW), where the weightings of the component models were determined based on which component models performed the best in the recent past. Specifically, at the 1- and 2-month ahead forecast horizons, the ensemble model predicted the ground truth (grey silhouette) much more closely and with less variance than that of its component models, which tended to display more significant oscillation relative to the ground truth. At the 3-month ahead horizon, the ensemble and its component models tended to produce less accurate predictions in this specific example. Given the broad geographical scope of our study, these findings suggest that ensemble models are a suitable default choice for generalizable and robust forecasts.

**Figure 2:**
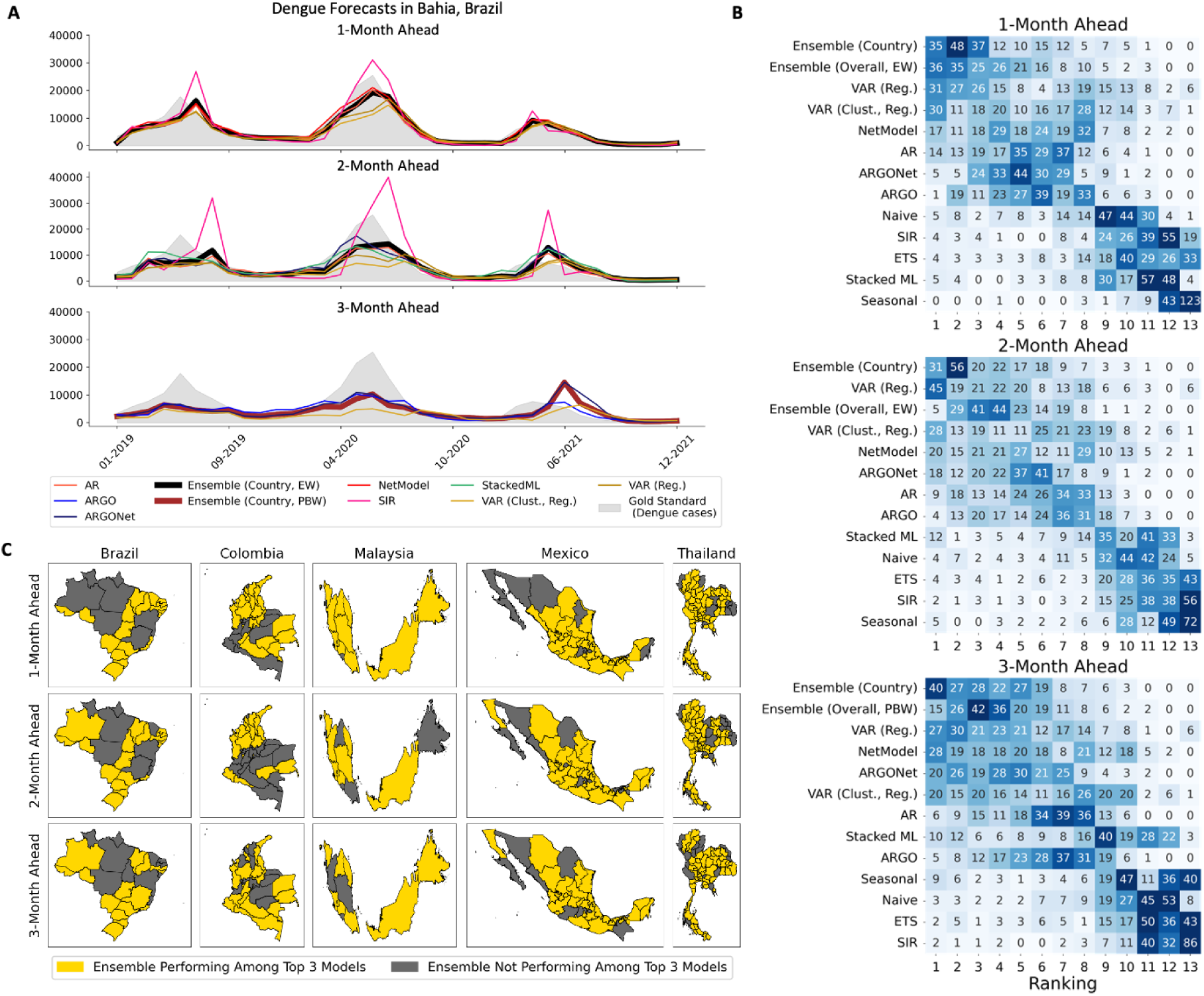
A summary of our prediction tasks and optimized model performances across 187 locations. **(A)** An example of our fine-tuned country-specific ensemble variants’ forecasts compared to their optimized component models in one selected location — Bahia, Brazil. The gold standard ground truth of reported dengue cases is shown as the grey silhouette. Ensemble predictions are shown in thick, bolded lines, while component models are shown in thinner, colored lines.**(B)** Heatmaps of the number of locations where each model attained a specific ranking in terms of mean absolute error with respect to the ground truth reported dengue case counts across all 187 locations. **(C)** Geographical maps of Brazil, Colombia, Malaysia, Mexico, and Thailand showing provinces where either the country-specific or overall ensemble performed in the top 3 rankings for each location (in yellow) and where they did not (in grey).

The heatmaps in Fig. 2 (B) show that the country-specific and overall ensembles had the most locations where they performed in the top 3 rankings in terms of percent absolute error, followed by regularized VAR and clustered + regularized VAR, out of the 187 tested locations. At the 2-month ahead horizon, country-specific ensembles still garnered the most top 3 rankings, but regularized VAR seemed to garner more top 3 rankings than the overall ensemble. At the 3-month ahead horizon, the country-specific and overall ensembles again attained the greatest number of top 3 rankings, followed by regularized VAR. While ensembles may not always be the absolute winner in every tested location, overall, they consistently placed on the top 3 rankings, being reliable and robust model choices. From the geographical maps in panel (C), ensemble models performed in the top 3 rankings in a vast majority of all 187 locations tested, as indicated by most of the maps being colored yellow. We found only a few consistent exceptions to this rule primarily in central Brazil and Colombia. These maps again corroborate the finding that ensembles are an effective option for stakeholders.

### Ensembling also yields improvement in error distributions across locations

We analyzed the potential error reduction of our ensemble models with respect to individual models. Our results are displayed in 3, which shows the percent absolute error distributions for the individual and the ensemble models in the 187 locations tested. Fig. 4 shows the same percent absolute error distributions of the individual and ensemble models within each country and forecast horizon.

### Overall performances

Despite the minor reduction in some cases, the country-specific and overall ensembles had the lowest mean percent absolute errors compared to all other models across all forecast horizons. Fig. 3 (A) to (C) displays the error distributions for each model, across the 3 different horizons of forecast. In all cases, we can observe that the Ensemble models were placed as the top performers, reaching values of 38.5%, 54.5% and 62.7% in terms of Percent Absolute Error (% AE). The next best model were there VAR variants (one incorporating regularization, and other implementing both regularization and clustering), reaching values of 40%, 55% 64%, for each respective task.

**Figure 3:**
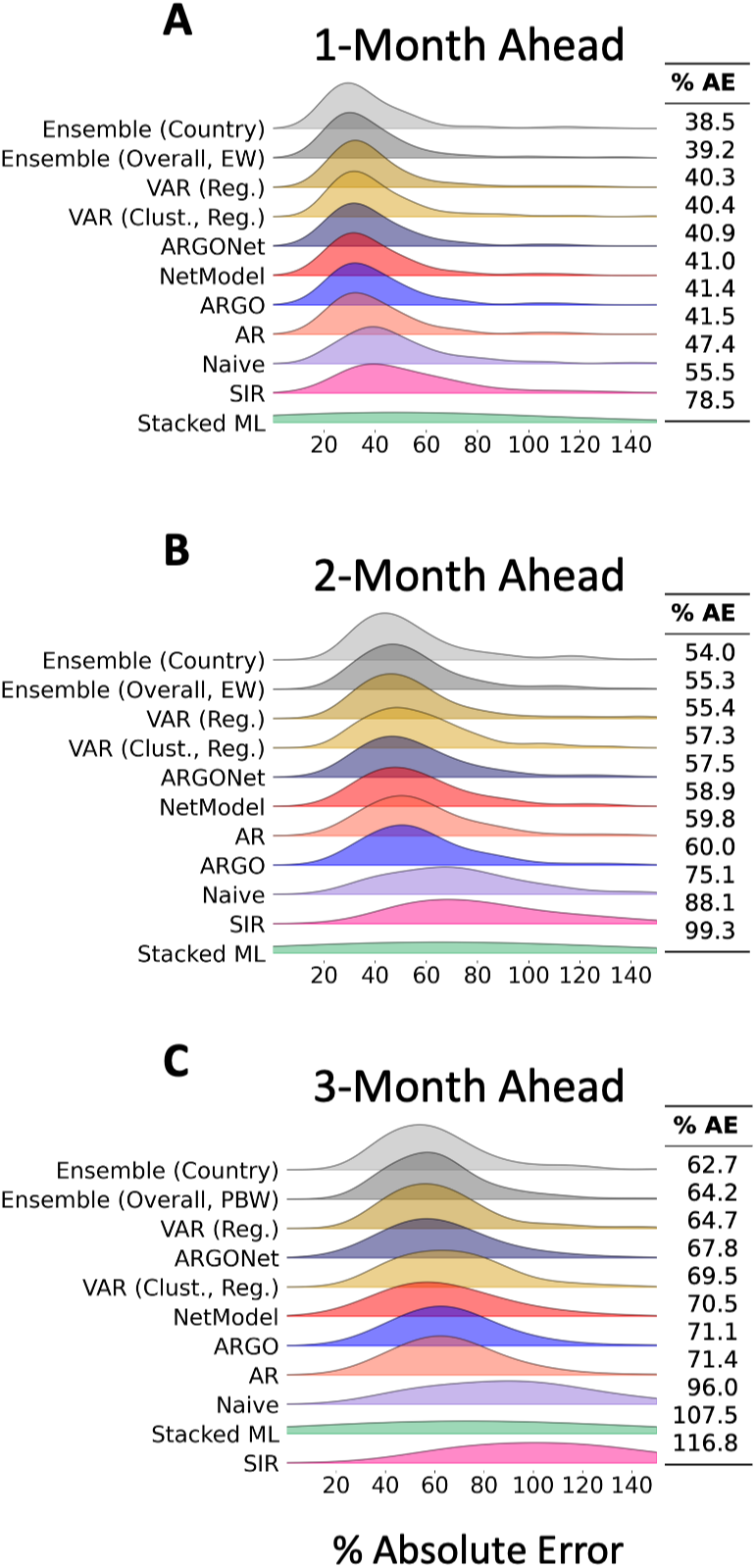
Overall error distributions for optimized individual and ensemble models across all 187 tested locations. (A) - (C) Ridgeline plots show percent absolute error distributions at the 1-month, 2-month, and 3-month horizons, respectively. Side tables record the mean percent absolute error incurred.

### Performance by country

Fig. 4 shows the performance per country. Notably, an ensemble variant achieved the lowest mean percent absolute error in 14 out of 15 tested combinations of forecast horizon and country. The sole exception was Colombia at 2-months ahead, with clustered + regularized VAR having achieved the lowest mean percent absolute error. However, the mean percent absolute errors and the shapes of the errors’ distributions are very similar for the top-performing models. Nonetheless, our analyses shown in this figure still confirm that the ensembles were greater than the sum of their component models, even when looking only within a particular country.

**Figure 4:**
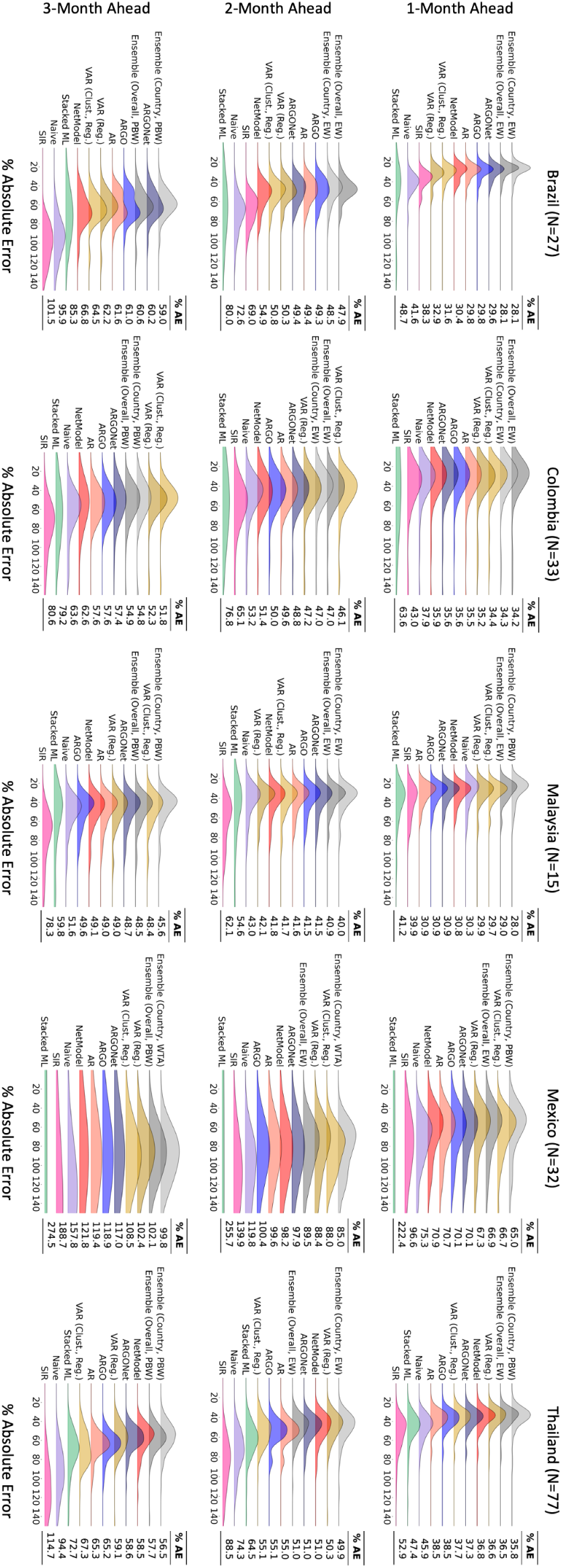
Country-specific error distributions for optimized individual and ensemble models, organized by country and forecast horizon. Ridgeline plots show percent absolute error

### Prospective Study: a real-life application and analysis of our methodology

In this section, we evaluate the performance of our forecasts generated by a real-time dengue activity forecasting platform. This platform was designed as a prospective tool to predict where dengue outbreak activity would occur 1, 2, and 3 months into the future. There are two primary differences between our retrospective and prospective studies. First, the prospective study was conducted in a real-world scenario where future ground truth data was unknown, whereas, in the retrospective study, we had access to the entire time series beforehand. Second, due to reporting delays in dengue case data, our prospective predictions did not utilize the most up-to-date epidemiological data. In contrast, the retrospective predictions were made using the most complete datasets available. This evaluation covered various locations in Brazil, Colombia, Mexico, Thailand, and Panama from May 2022 to July 2023. The forecasting models included Support Vector Machine (SVM), K-Nearest Neighbors (KNN), a regularized version of Vector Autoregression (VAR), and Seasonal Autoregressive Integrated Moving Average (SARIMA). Our ensemble models comprised a weighted ensemble, which assigned weights based on the mean squared error score of each model over the past six months, and a winner-takes-all ensemble, which selected the prediction from the ‘best’ base model with the least mean squared error over the past 3 months. Additionally, we used Persistence as our baseline model.

Figure 5 and Table 1 show a summary of the number of times our ensemble’s Mean Squared Error was among the top 3 best over all the locations within a Country (the analysis was repeated over each forecasting horizon). We also present the overall error reduction of each model with respect to persistence 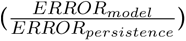 using a set of violin plots. We conducted the analysis for each location and each country (Figure 6).

**Table 1:** Summary of the number of times a model scored a Mean Squared Value among the top 3 best, across every location, and every horizon. Our results show that our weighted ensemble, VAR and Winner takes all approach are among the top performers.

| Top 3 overview |  |
| --- | --- |
| Weighted Ensemble | 3318 |
| VAR (regularized) | 2989 |
| Winner Takes All | 2310 |
| SARIMA | 1638 |
| KNN | 1519 |
| SVM | 1512 |
| Persistence | 658 |

#### Top 3 Analysis

Figure 5 shows the number of times a model reached within the top 3 Mean Squared Error reductions, for each country and each horizon. Each barplot represents the performance of a model within a Country, for a different prediction horizon. We can see VAR appearing eight times on the leftmost side (40%), and our Weighted Ensemble appearing seven times(35%). Our Winner-Takes-all approach appeared only two times on the leftmost side (10%), but had a comparable count with the top model whenever it was on second position (see Thailand in horizon 1, Panama in Horizon 1 and 4, Brazil in horizon 1, and Mexico in Horizon 1 and 2). Overall, most of our base models and ensembles had a higher count than persistence, with exception to Colombia in horizon 1, Panama in horizon 2, and Mexico in Horizon 1. Table 1 shows a total count over all locations, and all horizons. Our weighted ensemble had a total of 3318 counts, followed by VAR with 2989 and Winner-Takes-All with 2310 counts.

**Figure 5:**
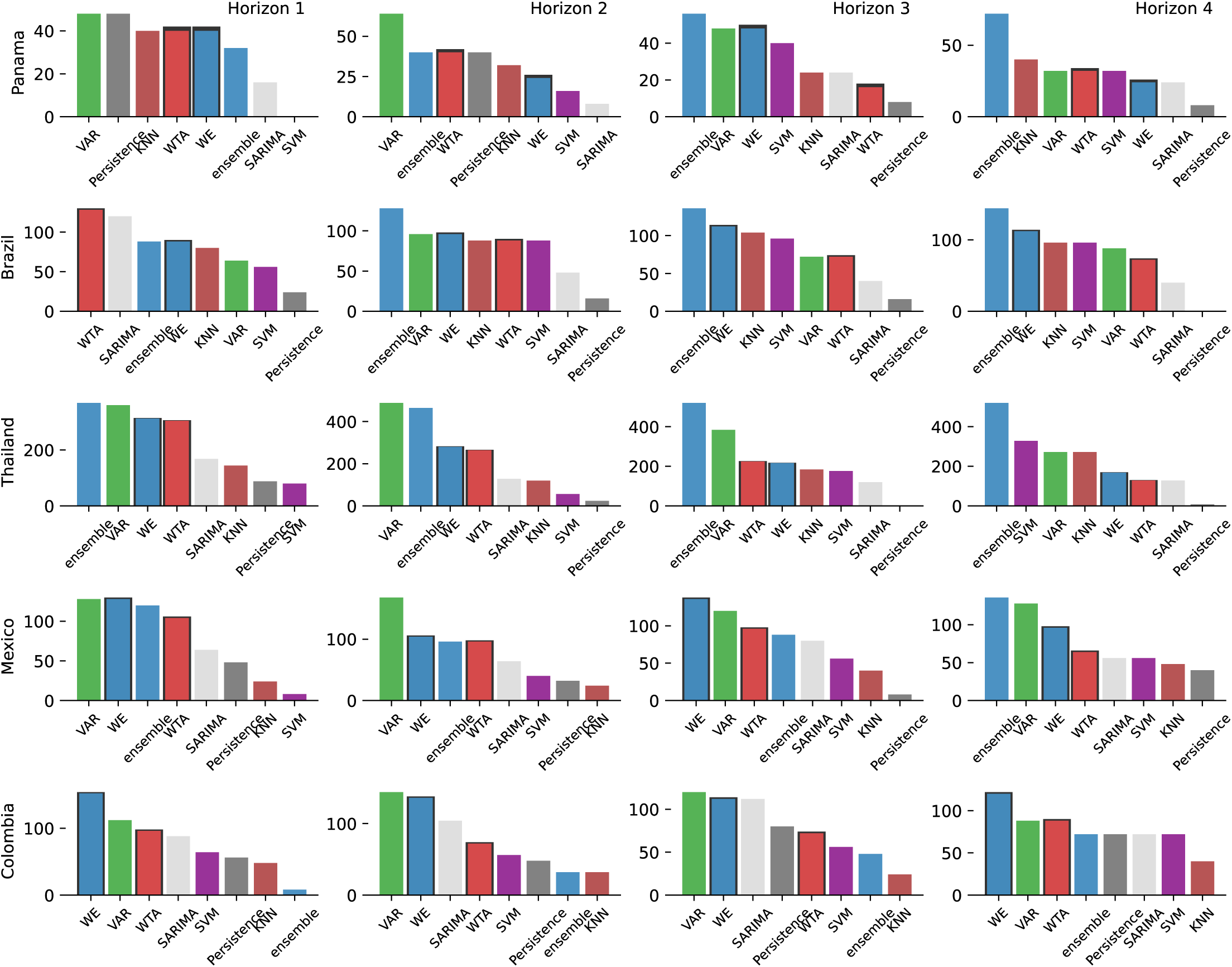
Top performer count. A visualization of the number of times a model scored a value of MSE rated among the top 3 best, ordered from left (models with the highest count) to right (models with the lowest count). Our results show that the Weighted Ensemble (blue), VAR (green), and the Winner-Takes-All approach are frequently among the top performers (leftmost side).

**Figure 6:**
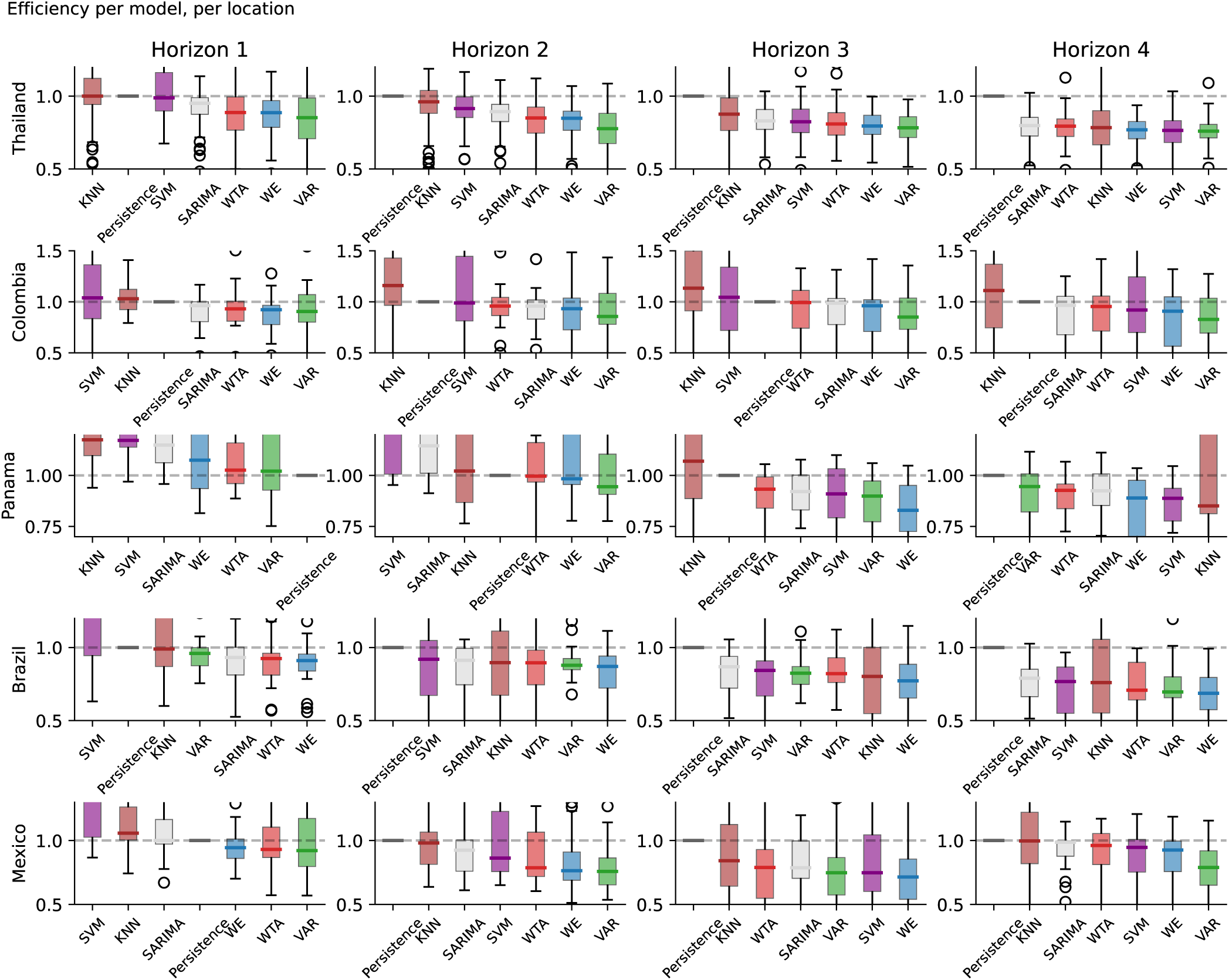
Error reduction with respect to the Persistence model. Summary of the error reduction of each model with respect to the Persistence baseline model. Each plot represents a different horizon (columns) and country (rows). Each violin plot visualizes a summary of the error reduction scores 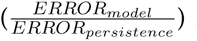 for a single model. Models are ordered from worst (left) to best (right). The gray dashed line represents the value of persistence and serves as a reference to validate if a model improved over the baseline.

#### 1.0.1 Overall Error reduction

Figure 6 exhibits a summary of the error reduction for the analyzed models with respect to the Persistence model 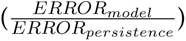. The violin plots are ordered so that the best model is to the rightmost side, and a dashed horizontal line plotted at *y* = 1 (*y* = 1 means the error of our model is equal to the error of Persistence) serves as a reference to know if a model consistently beat persistence or not.

The weighted ensemble (WE), vector autoregression and the winner-takes-all (WTA) ensemble were the three models that most frequently scored within top-3 error reduction. We observe that the weighted ensemble had median error reduction within the top 3 at every location and time horizon, except Panama in horizon 1. Regularized VAR scored the biggest error reduction in Colombia and Thailand for horizons 1,2, and 3, 4. Although less frequently, the Winner-Takes-All ensemble also remained within the top 3 performances with exception to Thailand in horizon 4, Panama in horizon 3 and 4, and Mexico in horizon 3 and 4.

#### Impact of reporting delays in our model’s performance

In conducting the prospective analysis, our forecasts were generated in a real-time scenario where the ground truth for each location was not fully reported at the time of prediction. The performance of our models were therefore likely affected by backfill issues in the data. Figure 7 illustrates our forecasts within the region of Sergipe, Brazil. At the time of prediction, the available information on confirmed cases (depicted in dark gray) differed from the most recently reported data (shown in light gray), which we employed as our ground truth for final metrics and error scores. Such backfill issues significantly impact real world applications as our models are trained solely on the information available at the given point in time.

**Figure 7:**
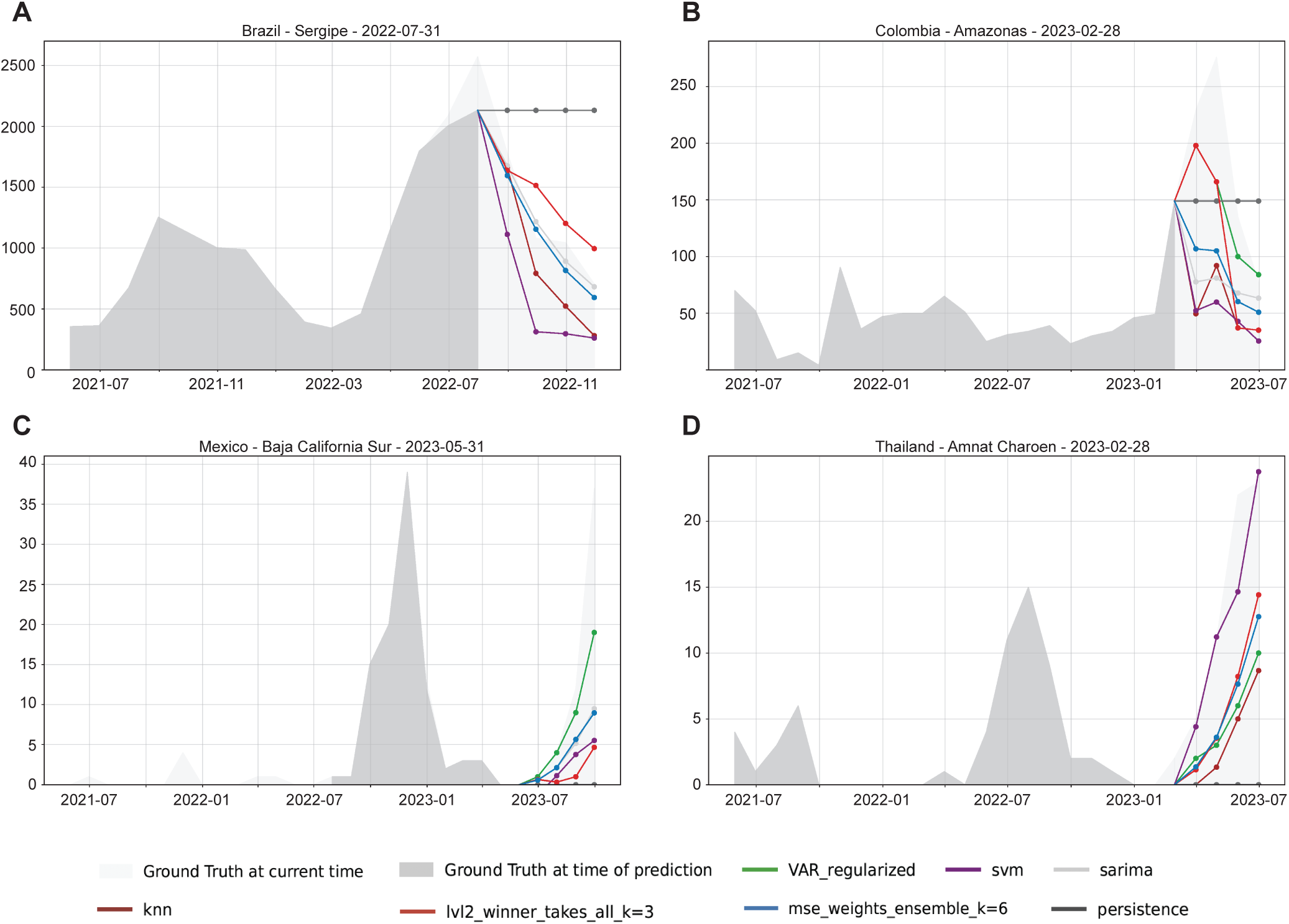
Visualization of the bias embedded in our models given reporting delays. The official reports known at the time of prediction, shown in dark gray, is the only information available to our models at the time of prediction. After several months, the ground truth changes due to backfill efforts based on the most recent reports are shown in light gray.

## Discussion

Dengue is a leading cause of hospitalization and death for many people around the world [16], and with cases doubling every ten years [26]. An essential component of dengue control is disease forecasting. Enhancing the accuracy and robustness of predictive models, particularly across multiple diverse geographical localities, empowers public health institutions to adopt a more proactive approach towards curbing the spread of the disease.

We tackled the task of predicting reported dengue fever case counts one, two, and three months ahead in various province-sized locations around the worldwide. Since individual model performances typically fluctuate across locations, there is a need for more robust and generalizable forecast models. In this context, our first contribution was the development of a family of ensemble models that retrospectively produced more accurate forecasts than their components across a broad range of geographically and socially diverse locations. Specifically, we investigated eleven types of data-driven, statistical, and mechanistic models as potential components of our ensemble. We also explored three ensembling mechanisms — performance-based weights, winner-takes-all, and simple average — and found that our ensemble models achieved lower percent absolute errors across 180+ geographically diverse locations. Our second contribution was a real-time dengue forecasting platform to predict when and where outbreaks will occur 1, 2, and 3 months in advance. Our forecasts were made in real-time without complete ground truth for each location. In fact, this task is not the same as performing retrospective studies, since dengue case data are typically updated months later (a problem commonly referred to as “backfill”). In this challenging scenario, our ensemble models still emerged as the top performers, producing better forecasts and reducing error compared to individual components.

Our ensemble models were robust predictors of dengue across the world. This fact is especially relevant because, as shown in Fig. 1, no individual model consistently achieved the lowest error. By contrast, while country-specific and overall ensembles were not always the best-performing models at a given location and forecast horizon, they almost always incurred the most top 3 performers compared to any of the component models. As shown in Fig. 3, our ensemble variants incurred the lowest error averaged across all 180+ tested locations in terms of percent absolute error compared to the component models. Even looking within a particular country and forecast horizon, as shown in Fig 4, ensemble models achieved the lowest error averaged across all locations within that country in 13 out of 15 combinations of country and forecast horizon. The two exceptions were Colombia at 2-months and 3-months ahead. Furthermore, in 9 out of 15 country-horizon combinations, both the country-specific and overall ensembles achieved the top 2 lowest errors averaged across locations. In our prospective study, the weighted ensemble (WE) model was overall the top performer in terms of low mean squared error, followed by VAR and the Winner Takes All (WTA) ensemble (Table 1). In terms of error reduction with respect to the persistence model, WE and WTA also consistently performed within top-3 performers across locations.

It is worth addressing the excellent performance of our VAR model in both retrospective and prospective studies. Examining our results more granularly, from Fig. 4, we observe that in Colombia, at the 2-month ahead horizon, VAR (Clust., Reg.) achieved more top 3 rankings than both ensemble variants. At the 3-month ahead horizon, VAR (Clust., Reg.) not only outperforms the overall ensemble in terms of the number of top 3 rankings but achieves more top 1 rankings than both ensemble variants. Similarly, at least one VAR model also outperforms at least one ensemble variant in terms of the number of top 3 rankings in all three forecast horizons of both Malaysia and Thailand. We hypothesize that VAR’s stellar performance in Colombia, Malaysia, and Thailand can be significantly attributed to the fact that these three countries are “province-dense” in the sense that individual provinces are relatively geographically small and, by extension, extremely close to each other. For example, Thailand has 77 provinces compacted into a relatively-small total surface area. In contrast, Brazil has 27 provinces spread out across a much larger area. The consequence of this geographical difference is that population centers between Brazilian provinces are much farther apart, and thus network effects are much weaker than their Thai counterparts. As such, VAR is much more effective in Thailand than Brazil because there are significantly stronger network effects between provinces to capture in our models.

We also investigated component models that were not exhaustively hyperparameter tuned but rather deployed straight out of the box, which we refer to as “standard” models. For details, we refer the reader to our Supplementary Information. As shown in Figs. 12 and 14, while our standard component models are nearly all unable to outperform our naive persistence baseline in terms of percent absolute error as averaged across locations, our ensemble models comprised of these standard component models outperformed the naive persistence baseline consistently. As such, our ensembling approach can take relatively weak, unoptimized learners and output a much stronger and more robust prediction. In this sense, the ensemble still performs better than its components.

From Tables 6 - 8 in the Supplemental Materials, we observe that when forecasting 1-month and 2-months ahead, the ensembling method most commonly employed (albeit plurality, not majority) was the equal weights method, followed by the performance-based weights model. At 3-months ahead, however, the performance-based weights mechanism was employed in most countries, including the overall ensemble. There does not appear to be a clear trend with respect to the ensemble training window sizes used to fit the performance-based weights and winner-takes-all ensembles.

Since the success of our forecasts is measured by achieving a lower percent absolute error than the naive persistence baseline model, ensembling enables us to include the naive persistence model itself as a component. As shown in our standard model results in the Supplementary Materials, we observe that when working with standard, non-fine-tuned component models, nearly all of the best country-specific and overall ensembles were comprised of the naive and seasonal basic models, coupled with one or two other models. From a bias-variance tradeoff perspective, the naive persistence model has very low variance, given its absence of tunable parameters. While other component models may overfit to noise and thus incur large errors, the naive persistence, by being simple, provides a stable component to the ensemble and thus allows the ensemble to outperform the other models, including its components.

One limitation of our work is that of the eight non-basic models that we include as potential components into the ensemble, six of them — AR, ARGO, ARGONet, NetModel, VAR (regularized), VAR (clustered + regularized) — can be interpreted as belonging to a common family tree of linear models involving autoregressive terms. In fact, we did not include many models in our analyses that were non-linear with respect to historical (logged) reported case counts, and our resultant ensembles may not be expressive enough. In the future, one could consider including more expressive but also more heavily-parameterized models such as Random Forests [65] and neural networks [4, 65] into our ensemble lineup to potentially increase performance. However, as explored in [4], heavily-overparameterized neural networks may underperform compared to simpler regression models at short-term disease forecasting tasks. One could also include additional traditional time series forecasting techniques like Holt-Winter, as explored in [52], into the ensemble lineup for extra non-linear models. With the exception of ARGO and ARGONet, all of our models were trained exclusively using historical dengue-reported case counts. Future work could include models that leverage climate data and earth observations into our ensemble lineup, as explored in [12].

Despite our efforts to forecast dengue cases in o ver 180 locations worlwide, there are still many other countries, especially in the Americas and Asia, where we can retrospectively and prospectively test our individual and ensemble models. Our methodology can also be easily extended to support uncertainty quantification. Please see our Supplementary Materials for additional details and proofs-of-concept of such an extension. Future studies could explore classification tasks of predicting whether a given location will experience an outbreak by thresholding our case count predictions. Methods like DT-SIR, while prone to over-predicting at outbreak peaks, are still excellent at capturing the outbreak progression trend. Another interesting research avenue involves combining regression and classification components together within ensembles. For example, one can consider an ensemble setup containing both regression models (predicting the number of dengue reported case counts) and classification models (predicting whether an outbreak will occur in the next months). Future work could also involve combining ensembles together into superensembles to further reduce variance.

## 2 Methods

### 2.1 Data Sources

In this section, we present our data collection and processing routines.

#### Reported Case Counts

We used two primary modalities of data to train our models. Raw weekly and monthly reported dengue case counts at the city and province levels were obtained from the following sources. For Brazil, data was obtained from SINAN (using the `Datasus` package in R) and from the Info Dengue website API (at https://info.dengue.mat.br). Data from both sources were reformatted and merged into a single data set. Area codes were translated to state and municipality names. For Thailand, National Disease Surveillance Reports were downloaded from http://doe.moph.go.th. Separate files are available for dengue fever, DHF, and DHF shock syndrome. Files were processed in R, reformatted and combined into a single data set. For all other countries, PDF-formatted reports were downloaded from the Ministry of Health websites (for Colombia: https://www.ins.gov.co; for Malaysia: https://www.moh.gov.my; for Mexico: https://www.gob.mx; for Peru: https://www.dge.gob.pe and for Puerto Rico: https://www.salud.gov.pr). Tables containing dengue case data on a regional level were identified and extracted using ABBYY FineReader PDF software, applying OCR where required, and saved to excel. Extracted tables were then processed in R, checking extracted region names and count values using regular expressions and verifying table totals where available. All tables were time-stamped with the date of the report they were extracted from, reformatted and combined into a single data set per country. Data at the city level were aggregated via summation into province-level resolution before input into our model training pipeline. All data at the weekly level were also aggregated via summation into monthly values before the start of model training.

#### Google Trends

We used the Google Health Trends API to obtain monthly dengue-related search terms’ frequencies at the province level for all of our locations. For a small number of locations where Google Trends data was not available at the provincial level, we used the Google Trends data of the dengue-related search terms at the country level as a proxy. Within each country, we used the same set of dengue-related search terms for each province. Country-specific lists of all the dengue-related search terms we used can be found in our Supplementary Materials.

To maintain consistency, we chose only the top 10 most useful dengue-related search terms in each country as input into the ARGO and ARGONet models that required Google Trends data. Specifically, we determined each term’s “usefulness” in a particular location by computing the Pearson correlation of these term’s search frequencies with the reported case counts on a time window directly preceding our model evaluation time window to avoid signal leakage. We ordered the Google Trends terms in decreasing order of Pearson correlation and used this ordered data as input into the ARGO and ARGONet models. Country-specific time windows for the Pearson correlation analyses can be found in our Supplementary Materials under the “Training Period” column of Table 2.

**Table 2:** Country-specific train and test periods. Specifically, the “Test Period” column denotes the range of dates for which predictions were generated by individual component models. Because ensemble models require an extra ensemble training window range, we generated ensemble model predictions starting one year after the individual component model start date. All metrics for both individual component and ensemble models presented in this paper are computed across the ensembles models’ test prediction ranges.

| Country | Training Period | Test Period |
| --- | --- | --- |
| Brazil | January 2010 - December 2017 | January 2018 - December 2021 |
| Colombia | January 2007 - December 2013 | January 2014 - December 2017 |
| Malaysia | January 2010 - December 2017 | January 2018 - December 2021 |
| Mexico | January 2013 - December 2017 | January 2018 - December 2021 |
| Peru | January 2001 - December 2008 | January 2009 - December 2012 |
| Puerto Rico | January 1991 - December 2008 | January 2009 - December 2012 |
| Thailand | January 2003 - December 2017 | January 2018 - December 2021 |

### 2.2 Fitting Methods

Fig. 8 illustrates our two methods for fitting our individual models at each timestep.

**Figure 8:**
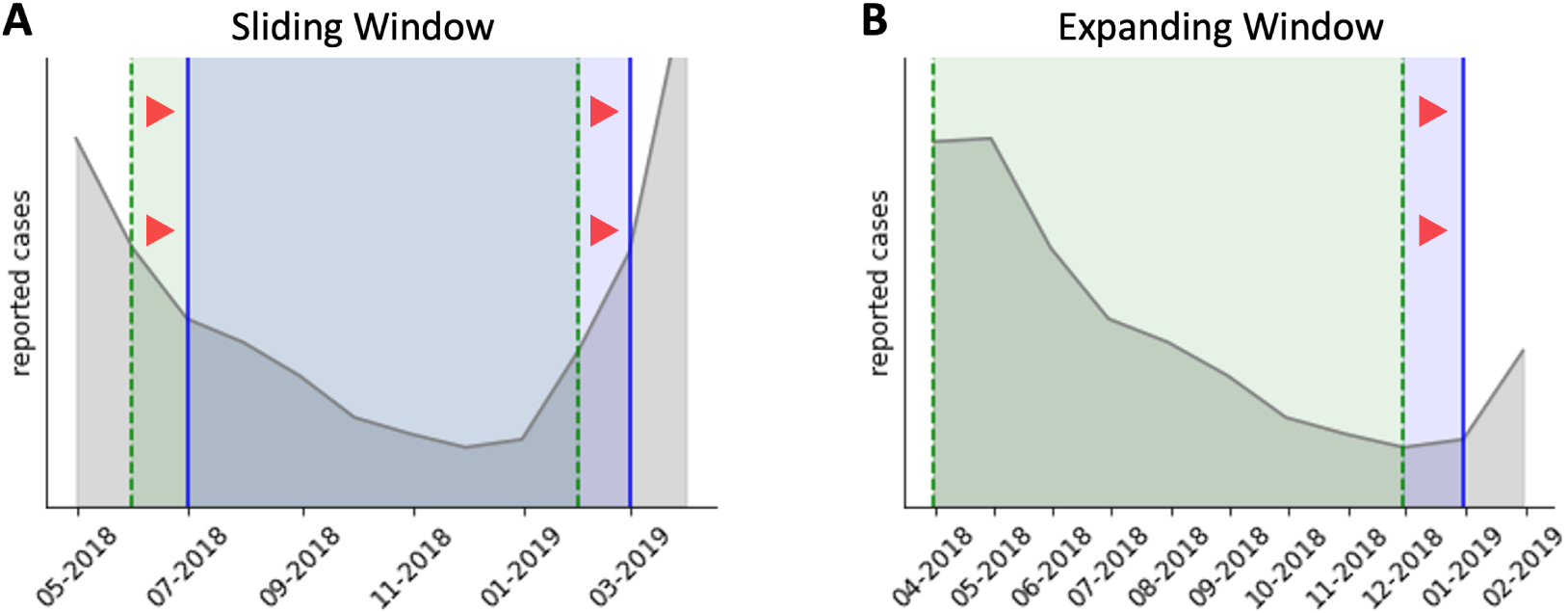
Schematics of model-fitting techniques. **(A)** Sliding window model-fitting. **(B)** Expanding window model-fitting.

Fig. 8 shows our two mechanisms for model-fitting. Panel (A) shows the sliding window mechanism for model fitting. For example, if the current time is February 2019, our prediction task is to forecast one month ahead — March 2019. And suppose, for example, that we are using a 9-month sliding window. Then, when fitting a given model to forecast for March 2019, a 9-month sliding window fitting method implies that we will only use the reported case counts data between June 2018 and February 2019 (inclusive) as target variables in our training set, as indicated by the green shaded region inside of the two dotted green lines. After March 2019, our new prediction task will be to forecast for April 2019, and we will *slide* our fitting window to fit our model using only the reported case counts data from July 2018 to March 2019 (now observed) as the target variables in our training set, as indicated by the blue shaded region inside of the two solid blue lines. To emphasize, because of the “sliding” operation, June 2018 is no longer contributing to our model fit. The assumption behind the sliding window mechanism is that infectious disease dynamics change across time, and thus, relationships between reported case counts in the distant past are likely not very informative of the current dynamics of the disease.

In contrast, panel (B) shows the expanding window mechanism for model fitting. Suppose that the earliest date in our dataset of reported case counts for which we can assemble a full set of covariate features is April 2018. Suppose that the current time is December 2018, and our goal is to forecast one month ahead — January 2019. Then, when fitting the model using an expanding window mechanism, we will use all of the reported case counts between April 2018 and December 2018 as target variables in our training set, as indicated by the region shaded in green inside of the two green dotted lines. After January 2019, our new prediction task will be to forecast for February 2019, and we will “expand” our fitting window to use all reported case counts from April 2018 to January 2019 as target variables in the training set, with the expansion region shaded in blue and bound by the solid blue line. In contrast to the sliding window, April 2018 is still in our training set. The underlying assumption behind the expanding window mechanism is that there exists some stationary distribution / ground-truth autoregressive data-generating process that holds across all time.

### 2.3 Individual Models

In this section, we describe the individual, fine-tuned component models that we will later ensemble together for more robust forecasts.

#### 2.3.1 Autoregression (AR)

As explained by [55], a *k*-month-ahead autoregressive model reported case counts in a specific location at month *t* + *k* as a linear combination of reported case counts at months *t* through *t* − *L* + 1 in said location, with a bias term. The hyperparameter *L* is the number of lags that we are using when forecasting. Initial experiments suggested that autoregressive models experienced significant performance boosts when working with log-transformed case counts, so we define *y_t_* as the log-transformed reported case counts of dengue in a given location at month *t*. An *AR*(*L*) model for *k*-month-ahead forecasting can thus be expressed as the following, where *ɛ* is an irreducible error term:

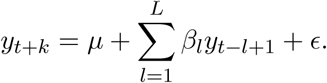

##### Standard Variant

For the standard variant, at all forecast horizons and locations, we used a generic *AR*(4) model fitted using Ordinary Least Squares, with no log-transformation of the reported case counts. We use an expanding window for model-fitting.

##### Optimized Variant

For the optimized variant, at all forecast horizons, we found that setting *L* = 24 yielded the best performance. We also trained all AR models in all locations and horizons using an expanding window approach. Computationally, we used the `glmnet` package [14] to minimize the L2 error subject to LASSO regularization (with regularization strength determined using cross-validation) to fit our model at each simulated month. From initial testing, we did not regularize the first two lags. For 2- and 3-months ahead forecasting, in addition to our choice of *L* = 24, we manually chose the specific lags of 1, 2, 3, 12, 13, 14, 15, and 24.

#### 2.3.2 AutoRegression with Google Search Data (ARGO)

As introduced in [63], ARGO builds on the classical AR model and incorporates the most-recently-available Google Trends search frequencies of dengue-related keywords as covariates into the linear model architecture. While ARGO was originally designed for flu incidence forecasting, it has also been adopted by [27] for dengue forecasting in 20 Brazilian cities. Let *x_m,t_* be the log-transformed Google Trends search frequency of term *m* (for *m* = 1 to *m* = 10) at month *t*. Then, the ARGO model with *L* epidemiological lags can be given by

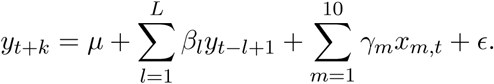

##### Standard Variant

For all forecast horizons, we used *AR*(4) coupled with the most recently observed Google Trends data as our features. We did use LASSO regularization via `glmnet` [14], but we did not protect any features from regularization. We also did not log-transform either the autoregressive features or the Google Trends features. We used an expanding window approach for model fitting.

##### Optimized Variant

Just as in AR, for all forecast horizons, we set *L* = 24 for maximum performance and use the same expanding window approach. We also use `glmnet` [14] with the same settings for model-fitting at each simulated month, with the first two epidemiological lags not subject to regularization. Unlike AR, we did not perform any manual selection of autoregressive lags and simply used all *L* = 24 lags as covariates.

#### 2.3.3 NetModel

To model the network effects of dengue spreading between nearby provinces, we extend the original AR linear model by modeling log-transformed reported case counts for location *j* during month *t* + *k* as a linear combination of only recent reported case counts in location *j*, but also recently reported case counts for all provinces *j*^′^ ∈ *J*, where *J* is the set of all provinces in a given country. A similar approach was implemented for flu forecasting in [31].

Let *L_a_* be the number of lags from location *j* itself (local lags) and *L_b_* be the number of lags from each of the other locations *j*^′^ ∈ *J* (neighbor lags) that we will be adding into our linear model. Let *y_j,t_* be the log-transformed reported case counts in location *j* at month *t*. Then, our NetModel is given by

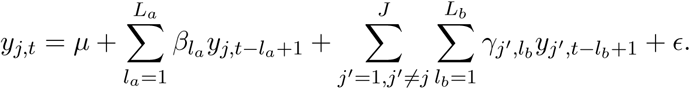

##### Standard Variant

For all forecast horizons, we used *L_a_* = 4 and *L_b_* = 1. We used LASSO via `glmnet` for model- fitting, but did not protect any lags from regularization. We used the expanding window fitting scheme for all locations and horizons. We did not log-transform the local lags nor the neighbor lags.

##### Optimized Variant

For 1-month ahead forecasting, we chose *L_a_* = 24, with manual feature-selection of the lags 1, 2, 3, 12, 13, 14, 15, and 24, and chose *L_b_* = 1, log-transforming both the local and neighbor lag features. We used `glmnet` with LASSO to fit our model, using an expanding window scheme, and refrained from regularizing the first two local lags.

For 2-months ahead forecasting, we chose *L_a_* = 12 and *L_b_* = 1, log-transforming both the local and neighbor lag features. We also used `glmnet` with LASSO to fit our model, using an expanding window scheme, and refrained from regularizing the first two local lags.

For 3-months ahead forecasting, we chose *L_a_* = 12, with manual feature-selection of the lags 1, 2, 3, and 12, and chose *L_b_* = 2, log-transforming both the local and neighbor lag features. We used `glmnet` with LASSO to fit our model, using an expanding window scheme, and refrained from regularizing only the first local lags.

#### 2.3.4 ARGONet

As introduced in [31], we also include ARGONet with two-component models — ARGO and NetModel — into our list of component models. Specifically, across all forecast horizons and locations, ARGONet returns the mean of the individual ARGO and NetModel predictions as its ensemble prediction. While ARGONet was also implemented in [31] with a winner-takes-all approach, we found that for the prediction task of forecasting monthly dengue-reported case counts, such a scheme was not as effective as simply returning the mean.

We implemented the standard / optimized variants of ARGONet by taking the mean of the corresponding standard / optimized ARGO and NetModel predictions, respectively.

#### 2.3.5 Exponential Smoothing

To implement exponential smoothing (ETS), we used the `sktime` [30] toolbox in Python. We used the pipeline functionality available within `sktime` with three different input-transforming preprocessors. The preprocessors we used were a power transformation, a robust scaler, and a min-max scaler. We then used AutoETS with a period of 12 (the number of months in a season) as the model in the pipeline. The pipeline was grid searched for the optimal input transformation using cross-validation with an expanding window splitter on the training data. After the pipeline hyperparameters were trained, it was used to make out-of-sample predictions in the usual manner with sequential addition of data, retraining parameters (not hyperparameters), and subsequent prediction.

#### 2.3.6 Vector Autoregression

We implemented multiple vector autoregression (VAR) models with varying degrees of regularization and clustering. Prior to modeling, the data were transformed with a standard log transformation. As with other components of the manuscript, we tested several transformation approaches, but none were consistently better than a log transformation. As the nature of VAR requires that data be available for all time points in every time series included in the model, we decided to individually implement a different VAR model for each country in the data. This allowed us to use nearly the entire time series available in each country. The alternative approach would be to combine all countries into a single model. Unfortunately, this would have seriously compromised the length of the time series in several countries as historical data varies considerably by country.

For modeling, we began with the most common method implemented in R through the `VARS` package [49]. Unfortunately, with the amount of data available, a standard VAR model in some countries (e.g., Thailand) would not converge even with a lag order of 1. However, much of the diminished performance could be resolved by implementing geographic clustering (discussed below), suggesting that the issue was primarily a result of some degree of underdetermination.

Nevertheless, the regularized models uniformly outperformed their unregularized counterparts, whether clustered or unclustered, so we proceeded with exclusively regularized models. In an effort to simplify the process of lag selection, we decided to standardize all regularized VAR models to use a lag order of 4. Any selection greater than 4 seemed to make no difference to the out-of-sample accuracy of the model.

For regularized VAR, we used primarily the `BigVAR` package made available in R [42]. We trialed several regularization frameworks, including (in the terminology of the package) Basic Lasso, Basic Elastic Net, Lag, Own/Other, Sparse Lag, Sparse Own/Other, Hierarchical Componentwise, Hierarchical Elementwise, and Hierarchical Own/Other. We used the standard rolling cross-validation method, expanding window, and adjusted the train/test window. However, there was no benefit to altering these from the default selections. We tested several lambda grid depth values and selected 100 as a good tradeoff between sufficient depth and reasonable compute time. We refer to this model in our figures and tables as “VAR (Reg.).”

#### 2.3.7 Vector Autoregression with Geographic Clustering

Next, we investigated to what extent geographic clustering could improve predictions. To that end, we obtained the latitude and longitude of every city in the data. We used the `sp` [7, 47] and `geosphere` packages available in R to compute the within-country distance matrix between all cities using the Haversine distance. The cities were then grouped with hierarchical clustering. The tree was produced using `hclust` and the clusters were produced using `cutree` as the highest number of clusters that allowed two or more cities in every cluster (which is a requirement for the application of VAR.

After within-country geographic clustering, VAR and regularized VAR were applied in the same manner as above. In all figures and tables, we refer to this model using the abbreviation “VAR (Clust., Reg.).”

#### 2.3.8 Stacked Machine Learning

We produced a stacked regression (stackedML) model using `sktime` [30] and `scikit-learn` [48]. We constructed it as an ensemble of several available machine-learning models in the toolboxes.

Given the challenges posed by underdetermination for these data when using higher lag orders on univariate time series, before implementing the model, we utilized a simple pipeline with a preprocessor and an elastic network (EN) base model that we optimized for the best L1 ratio. We then used this model to trim higher lag orders that seemed to improve this simple model.

After trimming the higher lag orders, we implemented the stackedML model again as a pipeline to allow comprehensive hyperparameter selection via grid-searched cross-validation. For preprocessing, we included the standard scaler, the min-max scaler, and a log transformation. For base models, we included an EN model, a k-nearest neighbors model (KNN), a support vector machine model (SVM), and a gradient-boosted machine (GBM) model. The pipeline was ensembled with an independently cross-validated elastic network model allowing only positive covariates. Hyperparameters in the base models were optimized via a grid search of the entire pipeline at once; the hyperparameters that we tuned included the number of neighbors in the KNN base model, the C-parameter in the SVM base model, and the number of leaves in the base GBM model.

After extensive testing, there were a few obvious limitations. First, optimizing the pipeline was extremely computationally intensive and became exponentially complex as more hyperparameters were searched. Second, the first issue was particularly challenging when combined with rolling optimization and leave-one-out (out-of-sample) cross-validation. Third, the data of a single-city univariate time series was clearly insufficient to tune an extremely large hyperparameter space. As a result, we pared the base models to remove the GBM. The GBM alone requires tuning of so many hyperparameters for optimal predictions that it was not improving the model with its inclusion. Then, we observed that the KNN was essentially never used by the ensembling model, so it was removed as well. As a result, we were left with an EN and an SVM model stacked together.

#### 2.3.9 Dynamically-Trained SIR

We introduce a novel, dynamically-trained SIR (DT-SIR) interpolator model for forecasting reported dengue cases. This model borrows the mathematical behaviors and properties of the traditional SIR dynamical system as introduced by Kermack and McKendrick [25] but re-purposes it for forecasting tasks.

As presented in [39], the traditional SIR model is governed by the following system of differential equations, parameterized by time *t*, where *S* is the number of susceptible individuals, *I* is the number of currently-infected individuals, *R* is the number of recovered (including deceased) individuals in a given population:

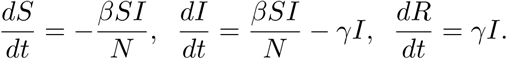

Here, *N* is the total number of people in the population, which we assume to be fixed. The parameter *β* governs the rate at which susceptible individuals (in *S*) become infected, and the parameter *γ* governs the rate at which infected individuals (in *I*) become recovered (or deceased). Mathematically, we can define any set of solution trajectory curves for *S*(*t*)*, I*(*t*)*, R*(*t*) for any interval of time starting with *t* = 0 by specifying *β* and *γ*, as well as specifying our initial conditions *S*_0_ and *I*_0_ for the numbers of susceptible and infected individuals at our initial point of reference *t* = 0. Since the SIR model assumes for all timesteps *t* that *S* + *I* + *R* = *N*, *R*_0_ is always uniquely determined by *S*_0_ and *I*_0_.

We acknowledge that the SIR model, originally designed for modeling direct transmission-type diseases, is an oversimplified model for dengue, given the complex combination of mosquito vector-borne transmission dynamics and intricate systems of partial immunity acquisition to different serotypes. However, for the sole purpose of forecasting case counts, our empirical results suggest that the basic SIR model is still sufficient. In fact, the mechanisms of the mosquito intermediary between infected humans for dengue transmission can be absorbed into / suitably approximated by the basic SIR model. It is also worth noting that using more complex models with more compartments and parameters risks over parameterization and overfitting, which could be undesirable regarding bias-variance tradeoff considerations.

It is important to note that our public health data provides *new* infected cases per month and not the total infected number of infected people within a population at a given time. However, because the recovery period of dengue is almost always less than a month [1], we can assume that all individuals who become newly infected in month *m* will also become recovered in the same month *m*. With this train of thought, it follows that we can treat *new* infected cases at month *m* as interchangeable with total infected people within the population at month *m*. However, from previous works in the literature, we know that dengue case reporting rates are very low. This is due in part to the reality that dengue fever oftentimes manifests no symptoms and that even when symptoms are present, they are oftentimes similar to that of the common flu. As such, reporting rates tend to be low. To accommodate this underreporting, we introduce a learnable report rate parameter *r* ∈ (0, 1) that captures the proportion of infected individuals who are recorded by public health authorities. Given *S*(*t*)*, I*(*t*)*, R*(*t*), let us define *C*(*t*) = *r*×*I*(*t*) to represent the number of *reported* infectious people within the population at month *t*, which we clarified above is interchangeable with the reported number of new infections at month *t*.

For each month *m*, DT-SIR outputs *reported* case count predictions for the month *m* + *h* at a single location through the following algorithm. For simplicity, suppose we are forecasting 1- month ahead, with *h* = 1. But, we can output forecasts for the 2- and 3-month-ahead horizons analogously.

1. We query the historical dengue reported case counts for the past *T* months: *m* − *T* + 1*, m* − *T* + 2*, …, m* − 1, and *m*. In practice, we found through extensive testing that *T* = 5 performed the best across all locations and horizons, and thus we set *T* = 5 for both our standard and optimized DT-SIR variants. Let us denote these historical case counts as *y_m_*_−_*_T_* _+1_*, y_m_*_−_*_T_* _+2_*, …, y_m_*_−1_, and *y_m_*.
2. Using the `scipy.integrate.odeint` numerical integration package from `SciPy` [60] and the non-linear least-squares curve-fitting package `lmfit` [40], we find the best set of parameters *β, γ, S*_0_*, I*_0_*, r* such that the resultant integrated *C*(*t*) curve (always calibrated to start mathematically from *t* = 0, corresponding to month *m* − *T* + 1) best fits the historical dengue reported case counts in the past *T* months. To emphasize, the solution curves to our SIR differential equations systems will always be plotted mathematically starting with *t* = 0, regardless of what month *m* we are in. We can do this because our initial conditions of *S*_0_ and *I*_0_ render our resultant trajectory curves agnostic to the realtime month/phase of an outbreak that we are in. We define “best fit” as minimizing the RMSE of the resultant *C*(*t*) curve (evaluated at *t* = 0, 1, 2*, …, T* − 1) with respect to the historical dengue case counts observed at months *m* − *T* + 1*, m* − *T* + 2*, …, m* − 1, and *m*. Because this objective function is almost certainly non-convex, we cannot guarantee that our curve-fitting algorithm will find the global minima. The best we can do is find a very good local optimum.
3. Given that we are only using the most recent *T* months’ observations as input data for fitting our SIR system parameters, in other words, we are using a sliding window approach and shifting our sliding window after each prediction timestep. The reason for using a sliding window as opposed to an expanding window that we use for the other component models is because the SIR model, by nature, can only model one outbreak peak at a time. If we used an expanding window approach, we may have multiple peaks in our fitting data, and our resultant SIR parameter fit would be very poor. The sliding window approach is especially attractive if we accept the assumption that disease outbreak dynamics vary significantly across time and various historical outbreak cycles. As such, we must re-estimate our model parameters at each timestep to remain up-to-date with current transmission dynamics.
4. When fitting our *C*(*t*) curve to the observed monthly data, it should be noted that we limit the plausible range for *β* ∈ [0, 500] and *γ* ∈ [0, 8.5]. We limited *β* to a still sizeable range mainly for practicality and reproducibility. From existing literature [1], we know that the average human infectious period for dengue fever is 4-5 days. Canonically, we know that *γ* represents 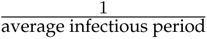. As such, converting to months, we find corresponding *γ* values of 6 and 7.5. We expand the upper bound of our parameter interval by 1 to account for some possible anomalies. Of course, we also restrict *S*_0_ and *I*_0_ to never exceed *N*, the total population. We restrict *r* ∈ [0, 1].
5. To avoid being confined to one local optima, we perform 500 independent fits for *β, γ, S*_0_*, I*_0_*, r*, selecting the estimated parameters corresponding to our ”best” fit (in terms of RMSE) for our forecasting purposes. For 250 of these fits, we randomly initialize our parameter guesses across their entire permitted intervals. For the other 250 fits, we initialize the starting parameter guesses to be distributed uniformly on an interval that is within 20% of the previous timestep’s fitted optimal parameter values, to enforce some “continuity” of our disease dynamic parameters over time. For *S*_0_ and *I*_0_, we initialize *S*_0_ during each fit to a starting guess of *N*, the total population, because intuitively, the proportion of people with dengue in a population is relatively low.
6. Using our best-estimated parameters of *β*, *γ*, *S*_0_, and *I*_0_, and *r*, we evaluate *C*(*t*) at *t* = *T* − 1 + *h* to produce our provisional prediction for the number of reported dengue case counts at month *m* + *h*. Let us call our provisional prediction *ŷ_m_*_+_*_h_*.
7. However, as observed in [58], SIR-type models are prone to “overshooting,” or significantly overestimating the number of reported dengue cases at outbreak peaks. Our DTSIR, without modification, also experiences such limitations. To mitigate this potential inaccuracy, we implement an anti-overshooting mechanism at each prediction timestep *m*, comprising of a ”threshold” and a ”compensator”. We will explain this anti-overshooting mechanism for the 1-month-ahead forecasting task (i.e., *h* = 1), but the 2-month and 3- month-ahead setups are analogous.

(a) Threshold: We calculate *y_t_* − *y_t_*_−_*_h_*, the observed *h*-month-apart differences between reported dengue case counts, for the past *n* months. Next, we calculate the mean observed historical differences in these past *n* months and add *s* standard deviations to this value. This computed value is our threshold *k*. Formally, we have

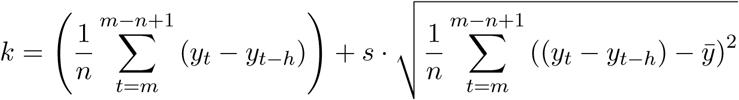

where *ȳ* is the sample mean of the *y_t_*−*y_t_*_−_*_h_*, for *t* = *m* to *t* = *m*−*n*+1. From extensive testing, we found that *n* = 24 and *s* = 4 were the most suitable and generalizable hyperparameter settings across all of our locations and forecast horizons.

(b) Compensator: If the difference between our provisional prediction for month *m* + *h*, *ŷ_m_*_+_*_h_*, and the true reported case count for our most recently observed month, *y_m_*, is greater than our threshold *k*, then our threshold is triggered and we adjust our prediction. Define *ȳ*^+^ to be the mean *positive h*-month-apart differences between reported dengue case counts for the past *n* months:

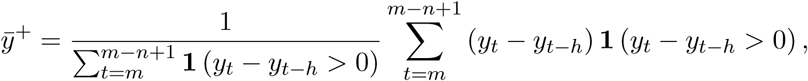

where **1** is an indicator function. Our compensator value is formulated as the sum of *ȳ*^+^ and *s* times the standard deviation of the *positive h*-month-apart differences between reported dengue case counts for the past *n* months. Let *δ* be our compensator value, which is formally defined as

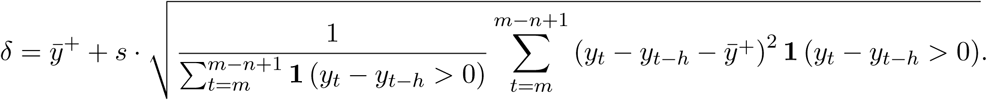

With our compensator value computed, we output the following adjusted prediction for month *m* + *h*, *ŷ*^′^_m+h_:

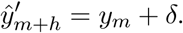

Intuitively, it would make sense to set *δ* = *k*. However, this alternative adjustment is suboptimal because setting it renders it very difficult for our model to predict an outbreak that is truly significantly more intense (in terms of peak reported case counts) than it has historically seen in its training data. In contrast, using our compensator formulation with the historical *positive* differences allows us to better capture the intuition that cases will indeed increase rapidly during outbreak peaks.

4. Finally, to predict reported case counts for the next month, month *m* + *h* + 1, we slide our fitting window forward by one month and repeat the steps enumerated above.

We use similar DT-SIR model settings in both our standard and optimized ensembling tests, for all locations and all forecast horizons — specifically, we use the most recent *T* = 5 months of observations as our sliding training window. However, for the standard model setting, we disable the anti-overshooting mechanism.

#### 2.3.10 Basic Models

In addition to the more complex individual models described above, we also include two relatively-simple baselines as potential component models in our ensemble.

#### Naive Persistence

If we are simulating being in month *t* and forecasting *k* months ahead, the naive persistence model will return the number of reported cases currently observed in month *t* as its forecast for month *t* + *k*. We use the same naive persistence model in both our standard and optimized ensembling tests.

#### Seasonal

Suppose we are simulating being in month *t* and forecasting *k* months ahead. Without loss of generality, suppose that month *t* + *k* is January. Then, the seasonal model will query our historical reported case counts for all observed January reported case counts, and return the mean of all the historical January dengue case counts as its forecast for month *t* + *k*. The idea behind the seasonal model is that dengue has been found to be seasonal in many locations around the world (see [19], [38], [61]). We use the same seasonal model in both our standard and optimized ensembling tests.

### 2.4 Ensemble Systems

Fig. 9 illustrates the three ensembling methods that we tested. Let *ŷ_i,t_* refer to the prediction generated by model *i* for time *t*, **ŷ***_t_* represent our component models’ predictions for time *t* stored as a vector, and *y_t_* be the ground-truth reported case counts at time *t*. Panel (B) illustrates the Performance-Based Weights (PBW) ensemble. In our toy example shown in Fig. 9, for this prediction timestep and our choice of a 5-month ensemble fitting window, the PBW ensemble finds the optimal set of non-negative weights **w** that minimizes the following objective function, if we calibrate *t* = 5 to correspond to the last observed timestep of April 2021: 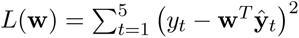, subject to the constraint that **1***^T^* **w** = 1. Such optimization was performed using the `scikit-learn` [48] package. To make our ensemble prediction for time *t*+1, we output said prediction as **w***^T^* **ŷ***_t_*_+1_, where **ŷ***_t_*_+1_ is the vector of our individual models’ predictions for time *t* + 1. Panel (C) illustrates the Winner-Takes-All (WTA) ensemble. Extending the notation from our discussion of the PBW ensemble, the WTA ensemble outputs *ŷ_i_*_∗_*_,t_*_+1_ as our ensemble prediction, where 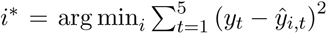 encodes the index corresponding to the component model that outputted the most accurate predictions during the ensemble fitting window. Panel (D) represents the Equal Weights (EW) ensemble, which simply outputs the unweighted mean of the components’ predictions at time *t* + 1 as the ensemble prediction.

**Figure 9:**
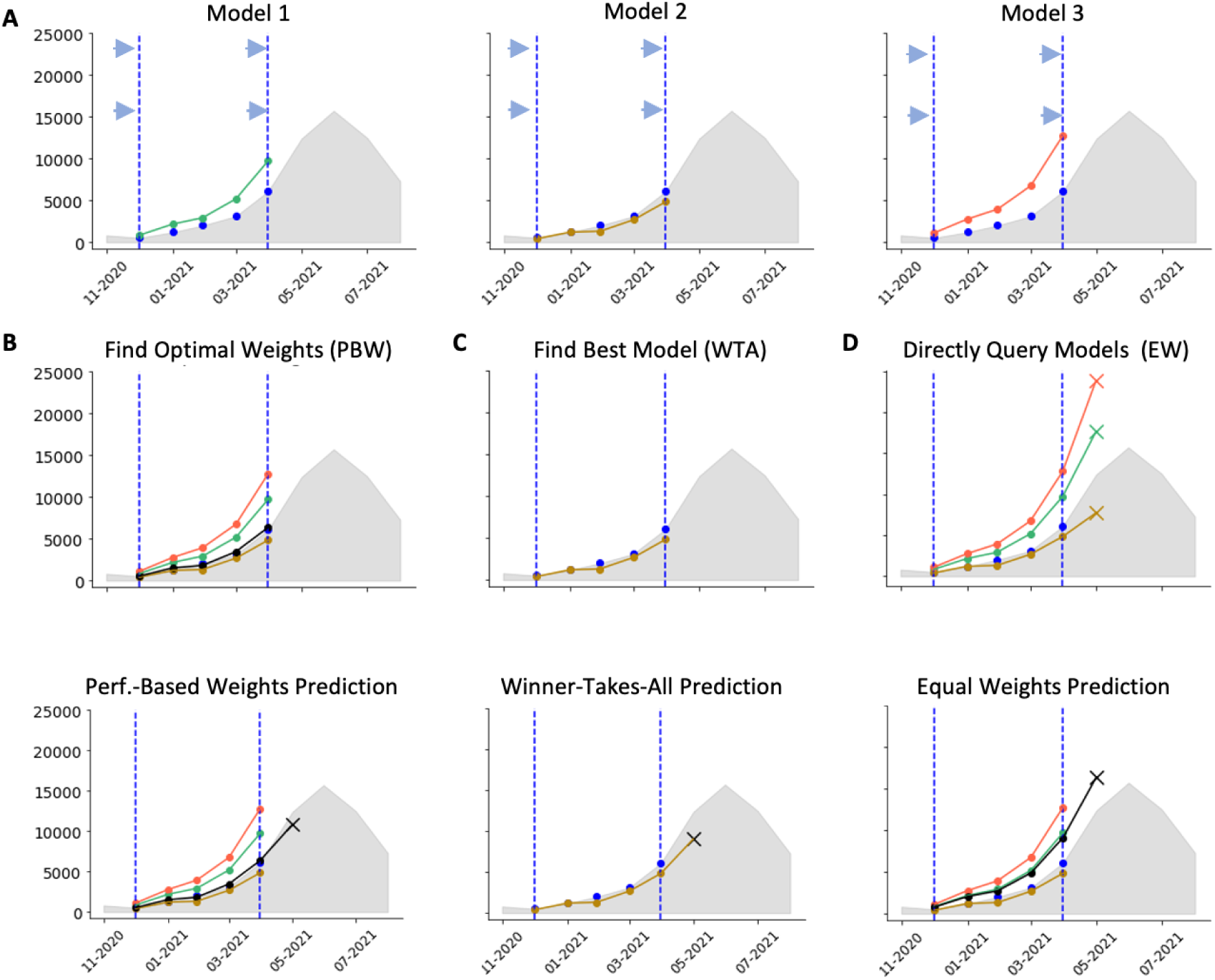
Schematics of the three ensembling methods. **(A)** Graphical representations of three individual models’ predictions during the ensemble fitting window. The ground truth reported case counts during the ensemble fitting window are depicted using the blue points. Each individual model’s predictions during the ensemble fitting window are shown with the connected lines. **(B)** Performance-Based Weights (PBW) ensemble fitting and prediction output. **(C)** Winner-Takes-All (WTA) ensemble fitting and prediction output. **(D)** Equal Weights (EW) ensemble prediction output.

**Figure 10:**
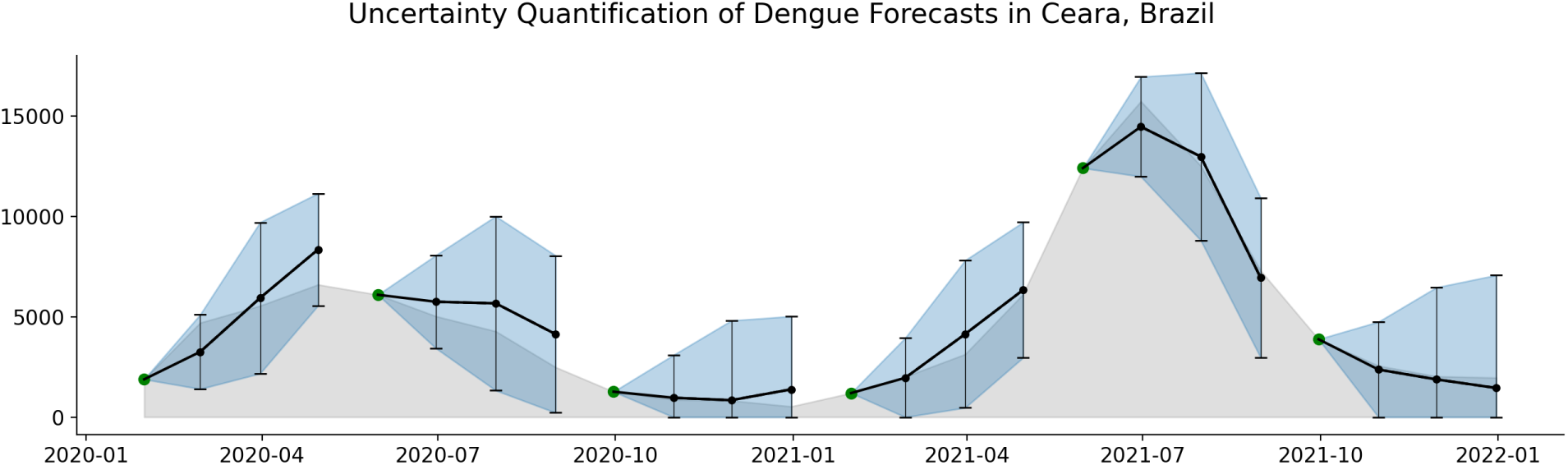
Demonstration of uncertainty quantification interval algorithm in Ceara, Brazil. The green points indicate the reference months *m*, and each set of subsequent three black points marks the 1-, 2-, and 3-month ahead point forecasts. The error bars corresponding to each point forecast quantify our 95% approximate predictive intervals. The blue cones emphasize how the uncertainty in our forecasts evolves over forecast horizon. The grey silhouette shows the ground truth reported dengue case counts.

Please see Tables 3-5 and 6-8 in the Supplementary Materials for the best ensemble variants using the standard and optimized individual models, respectively, at each forecast horizon and within each country.

**Table 3:** Best ensemble variants in each country for forecasting 1-month ahead using standard component models. “ETW” refers to ensemble training window.

| Country | Ensembling Method | ETW | Component Models |
| --- | --- | --- | --- |
| Brazil | Winner-Takes-All | 2 | AR, Naive, NetModel |
| Colombia | Performance-Based Weights | 3 | AR, Naive |
| Malaysia | Performance-Based Weights | 1 | ARGO, Naïve, NetModel |
| Mexico | Performance-Based Weights | 2 | AR, Naive, Seasonal |
| Peru | Performance-Based Weights | 3 | Naive, Seasonal |
| Puerto Rico | Equal Weights | 0 | AR, SIR |
| Thailand | Performance-Based Weights | 1 | AR, ARGONet, Naive, NetModel, Seasonal |
| Overall | Performance-Based Weights | 2 | AR, Naive, NetModel |

**Table 4:** Best ensemble variants in each country for forecasting 2-months ahead using standard component models. “ETW” refers to ensemble training window.

| Country | Ensembling Method | ETW | Component Models |
| --- | --- | --- | --- |
| Brazil | Winner-Takes-All | 1 | AR, Naive |
| Colombia | Winner-Takes-All | 4 | AR, Naive |
| Malaysia | Performance-Based Weights | 4 | AR, Naive |
| Mexico | Performance-Based Weights | 1 | AR, Naive, Seasonal |
| Peru | Performance-Based Weights | 1 | SIR, Seasonal |
| Puerto Rico | Equal Weights | 0 | SIR, Seasonal |
| Thailand | Performance-Based Weights | 3 | Naive, Seasonal |
| Overall | Performance-Based Weights | 1 | AR, Naive, Seasonal |

**Table 5:** Best ensemble variants in each country for forecasting 3-months ahead using standard component models. “ETW” refers to ensemble training window.

| Country | Ensembling Method | ETW | Component Models |
| --- | --- | --- | --- |
| Brazil | Performance-Based Weights | 6 | Naive, Seasonal |
| Colombia | Performance-Based Weights | 3 | Naive, NetModel |
| Malaysia | Winner-Takes-All | 2 | AR, Naive |
| Mexico | Performance-Based Weights | 1 | ARGONet, Naive, Seasonal |
| Peru | Performance-Based Weights | 1 | Naïve, SIR, Seasonal |
| Puerto Rico | Performance-Based Weights | 1 | SIR, Seasonal |
| Thailand | Performance-Based Weights | 2 | Naive, Seasonal |
| Overall | Performance-Based Weights | 2 | Naive, Seasonal |

**Table 6:** Best ensemble variants in each country for forecasting 1-month ahead using optimized component models. “ETW” refers to ensemble training window.

| Country | Ensembling Method | ETW | Component Models |
| --- | --- | --- | --- |
| Brazil | Equal Weights | 0 | AR, NetModel, SIR, VAR (Clust., Reg.), VAR (Reg.) |
| Colombia | Equal Weights | 0 | ARGONet, SIR, VAR (Clust., Reg.), VAR (Reg.) |
| Malaysia | Performance-Based Weights | 1 | ARGO, ARGONet, Naïve, Seasonal, VAR (Clust., Reg.), VAR (Reg.) |
| Mexico | Performance-Based Weights | 1 | ARGO, VAR (Clust., Reg.), VAR (Reg.) |
| Peru | Performance-Based Weights | 5 | ARGO, NetModel, Seasonal |
| Puerto Rico | Equal Weights | 0 | SIR, StackedML |
| Thailand | Performance-Based Weights | 12 | NetModel, VAR (Clust., Reg.), VAR (Reg.) |
| Overall | Equal Weights | 0 | ARGO, NetModel, SIR, VAR (Clust., Reg.), VAR (Reg.) |

**Table 7:** Best ensemble variants in each country for forecasting 2-months ahead using optimized component models. “ETW” refers to ensemble training window.

| Country | Ensembling Method | ETW | Component Models |
| --- | --- | --- | --- |
| Brazil | Equal Weights | 0 | AR, ARGONet, SIR, StackedML, VAR (Clust., Reg.), VAR (Reg.) |
| Colombia | Equal Weights | 0 | ARGONet, SIR, VAR (Clust., Reg.), VAR (Reg.) |
| Malaysia | Equal Weights | 0 | AR, ARGO, ARGONet, ETS, Naïve, NetModel, SIR, VAR (Clust., Reg.) |
| Mexico | Winner-Takes-All | 4 | ARGONet, VAR (Clust., Reg.), VAR (Reg.) |
| Peru | Performance-Based Weights | 1 | SIR, Seasonal |
| Puerto Rico | Equal Weights | 0 | SIR, StackedML |
| Thailand | Equal Weights | 0 | NetModel, VAR (Reg.) |
| Overall | Equal Weights | 0 | ARGONet, VAR (Clust., Reg.), VAR (Reg.) |

**Table 8:** Best ensemble variants in each country for forecasting 3-months ahead using optimized component models. “ETW” refers to ensemble training window.

| Country | Ensembling Method | ETW | Component Models |
| --- | --- | --- | --- |
| Brazil | Performance-Based Weights | 12 | ARGO, ARGONet, VAR (Clust., Reg.) |
| Colombia | Performance-Based Weights | 12 | SIR, VAR (Clust., Reg.), VAR (Reg.) |
| Malaysia | Performance-Based Weights | 4 | NetModel, StackedML, VAR (Clust., Reg.) |
| Mexico | Winner-Takes-All | 1 | VAR (Clust., Reg.), VAR (Reg.) |
| Peru | Performance-Based Weights | 1 | Naïve, SIR, Seasonal |
| Puerto Rico | Winner-Takes-All | 12 | SIR, StackedML |
| Thailand | Performance-Based Weights | 6 | NetModel, StackedML, VAR (Reg.) |
| Overall | Performance-Based Weights | 1 | ARGONet, VAR (Reg.) |

### 2.5 Model Evaluation

All individual and ensemble models were evaluated on the time periods under “Test Period” in Table 2 in the Supplementary Materials. While available data timeframes differed significantly across countries, we aimed to secure at least 3 years (the most recent years) of evaluation data points for each country. The remainder of the data were binned as training data, whether that be used directly for model-fitting, and/or for feature engineering.

To compare model performances, we used percent absolute error (PAE) on the predicted versus ground-truth reported case counts as our evaluation metrics. The “percent” implies that we divided the raw mean absolute errors for each model in a given location by the mean number of cases present in said location during the evaluation period.

## **3** Supplementary Materials

### 3.1 Extension of methodology for uncertainty quantification

Though not the focus of our main manuscript, our methodology can be easily extended to incorporate uncertainty quantification via approximate 95% predictive intervals for our forecasts. Our predictive interval generation algorithm is as follows:

1. Suppose today is month *m* and we would like to provide uncertainty quantification intervals for our *h*-month-ahead ensemble forecast. In practice, the ensemble can be replaced with any individual component model, too.
2. We can compute the predictive residuals *ɛ_t_* = *ŷ_t_* − *y_t_* for previously-observed months *t* = 1 through *t* = *m*, where *y_t_* was the true reported dengue case count at month *t* and *ŷ_t_* was our *h*-month-ahead dengue forecast for that month (i.e., generated in month *t* − *h*). Let us name our expanding-each-month vector of residuals at month *m* as ***ɛ_m_***.
3. Using our ensemble, we can generate our point-forecast for month *m* + *h*, and denote it as *ŷ_m_*_+_*_h_*. To provide uncertainty quantification, we can compute the standard deviation of this model’s historical residuals stored in ***ɛ_m_*** and, assuming approximate Normality, multiply by 1.96 to obtain the width of an approximate 95% predictive interval.
4. Then, our uncertainty-quantified ensemble forecast for month *m* + *h* would be

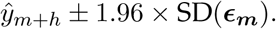

5. Because case counts cannot be negative, we may also clip our forecast intervals to be strictly non-negative.

Below, we demonstrate this uncertainty quantification algorithm on 1-, 2-, and 3-month country-specific ensemble forecasts in Ceara, Brazil, with approximate 95% predictive intervals generated for January, May, and September of 2020 and 2021.

#### No one individual model is best across all locations: model performance is significantly dependent on location

Fig. 1 summarizes the comparative performances of our optimized individual and ensemble models’ within each of the five countries, tested across the three prediction tasks of forecasting 1-, 2-, and 3-months ahead. The reader can find additional details on our fine-tuning and optimization processes in the Supplementary Information.

Panel (A) illustrates our three forecasting tasks of forecasting 1-, 2-, and 3-months ahead. Panels (B) to (F) show the models’ performances within each country in two ways. On the left of each panel, we present a heatmap where each row represents a model, and each column encodes the number of locations within the country of interest where a model achieved a certain rank in terms of percent absolute error (PAE) compared to the other individual component and ensemble models (1st through 13th rankings). The models in each heatmap are listed in decreasing order by the sum of the number of locations where each model performed in the first, second, or third ranks. As an example, “Ensemble (Country, EW)” having a value of 7 corresponding to Ranking 1 in Brazil (1-Month Ahead) means that the country-specific ensemble incurred the lowest (best) PAE compared to all other individual and ensemble models in 7 out of the 27 provinces of Brazil. On the right of each panel, we have a geographical map where each province is colored according to the model that incurred the lowest PAE in that province, with the legend displayed at the bottom of the overall figure. Overall, ensemble models were robust within any of our tested countries. They demonstrated the most or nearly the most top-3 rankings compared to other models.

Formally, we define percent absolute error (PAE) on the predicted versus ground-truth reported case counts as the raw mean absolute error divided by the mean number of monthly reported cases observed in a given location during the evaluation period. Mathematically, let *y*_1_ *… y_T_* be the ground-truth reported case counts, and *ŷ*_1_ *… ŷ_T_* be our predicted case counts. Then,

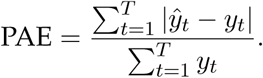

On the right of panels (B) - (F), we plot a map of each country along with the best-performing model for each province. We observed that no particular model consistently performs the best across all locations and prediction tasks within a country. Our findings thus suggest that, in practice, it is not feasible to train, evaluate, and find the best-performing models for every single province in every single country. Instead, using an out-of-the-box ensemble setup would require significantly less computation and optimization while still effectively guaranteeing strong performance. In what follows, we present the results for each country.

##### Brazil

Fig. 1 (B) shows that, within the 27 provinces of Brazil, the country-specific ensemble achieved the most top 3 rankings compared to any other model, with the overall ensemble having reached the second most top 3 rankings at the 1-month and 2-month horizons. However, at the 3-month horizon, ARGONet and AR outperformed the overall ensemble in the number of top 3 rankings. From the accompanying color-coded geographical maps, we observed that at any forecast horizon for Brazil, the ensemble did not rank first in many locations - the winning model varied widely across sites.

##### Colombia

Fig. 1 (C) shows our 13 models’ ranking in the 33 provinces of Colombia. While the country-specific and overall ensembles achieved the most top 3 rankings at the 1-month ahead forecast horizon, clustered + regularized VAR achieved the most top 3 rankings, superseding the two ensemble variants, at 2-month ahead. At 3-months ahead, the country-specific ensemble achieved the top 3 rankings, but the overall ensemble was still outperformed by clustered + regularized VAR.

##### Malaysia

Fig. 1 (D) shows our results for the 15 provinces in Malaysia. While the country-specific ensemble achieved the most top 3 rankings at the 1-month and 3-month ahead horizons, clustered + regularized VAR achieved the most top 3 rankings at the 2-month horizon. Notably, the overall ensemble did not generalize very well in Malaysia, being ranked even below the naive persistence baseline at the 1-month ahead horizon. We note, however, that the two easternmost (and largest) Malaysian provinces both saw the country-specific ensemble performing the best out of all 13 models at the 1-month and 3-month horizons.

##### Mexico

Fig. 1(E) shows our 13 models’ rankings in the 32 provinces of Mexico. Regularized VAR achieved the top 3 rankings at the 1-month ahead forecast horizon, followed by the country-specific ensemble. At the 2-month ahead horizon, the two ensembles achieved the top 3 rankings, though at 3-months ahead. In contrast, the overall ensemble maintained the top 3 rankings, regularized VAR overtook the country-specific ensemble for the second most top 3 rankings. From the geographical maps, we see that no individual model achieved the top rank across most of locations.

##### Thailand

In Fig. 1(F), we show the results in the 77 provinces of Thailand. From the heatmaps, we observed that the country-specific ensemble achieved the most top 3 rankings at the 1-month and 2-month horizons. By comparison, regularized VAR superseded both ensembles for the most top 3 rankings at the 3-month horizon. Notably, at the 2-month horizon, regularized VAR achieved more first-place rankings than both ensembles. At the 1-month horizon, regularized VAR achieved the same number of first-place rankings as the overall ensemble and significantly more first-place rankings than the country-specific ensemble. The geographical maps show that no particular model performed the best across most Thailand provinces.

#### No one individual model is best across all locations: model performance is significantly dependent on location

Fig. 1 summarizes the comparative performances of our optimized individual and ensemble models’ within each of the five countries, tested across the three prediction tasks of forecasting 1-, 2-, and 3-months ahead. The reader can find additional details on our fine-tuning and optimization processes in the Supplementary Information.

Panel (A) illustrates our three forecasting tasks of forecasting 1-, 2-, and 3-months ahead. Panels (B) to (F) show the models’ performances within each country in two ways. On the left of each panel, we present a heatmap where each row represents a model, and each column encodes the number of locations within the country of interest where a model achieved a certain rank in terms of percent absolute error (PAE) compared to the other individual component and ensemble models (1st through 13th rankings). The models in each heatmap are listed in decreasing order by the sum of the number of locations where each model performed in the first, second, or third ranks. As an example, “Ensemble (Country, EW)” having a value of 7 corresponding to Ranking 1 in Brazil (1-Month Ahead) means that the country-specific ensemble incurred the lowest (best) PAE compared to all other individual and ensemble models in 7 out of the 27 provinces of Brazil. On the right of each panel, we have a geographical map where each province is colored according to the model that incurred the lowest PAE in that province, with the legend displayed at the bottom of the overall figure. Overall, ensemble models were robust within any of our tested countries. They demonstrated the most or nearly the most top-3 rankings compared to other models.

Formally, we define percent absolute error (PAE) on the predicted versus ground-truth reported case counts as the raw mean absolute error divided by the mean number of monthly reported cases observed in a given location during the evaluation period. Mathematically, let *y*_1_ *… y_T_* be the ground-truth reported case counts, and *ŷ*_1_ *… ŷ_T_* be our predicted case counts. Then,

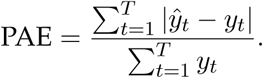

On the right of panels (B) - (F), we plot a map of each country along with the best-performing model for each province. We observed that no particular model consistently performs the best across all locations and prediction tasks within a country. Our findings thus suggest that, in practice, it is not feasible to train, evaluate, and find the best-performing models for every single province in every single country. Instead, using an out-of-the-box ensemble setup would require significantly less computation and optimization while still effectively guaranteeing strong performance. In what follows, we present the results for each country.

##### Brazil

Fig. 1 (B) shows that, within the 27 provinces of Brazil, the country-specific ensemble achieved the most top 3 rankings compared to any other model, with the overall ensemble having reached the second most top 3 rankings at the 1-month and 2-month horizons. However, at the 3-month horizon, ARGONet and AR outperformed the overall ensemble in the number of top 3 rankings. From the accompanying color-coded geographical maps, we observed that at any forecast horizon for Brazil, the ensemble did not rank first in many locations - the winning model varied widely across sites.

##### Colombia

Fig. 1 (C) shows our 13 models’ ranking in the 33 provinces of Colombia. While the country-specific and overall ensembles achieved the most top 3 rankings at the 1-month ahead forecast horizon, clustered + regularized VAR achieved the most top 3 rankings, superseding the two ensemble variants, at 2-month ahead. At 3-months ahead, the country-specific ensemble achieved the top 3 rankings, but the overall ensemble was still outperformed by clustered + regularized VAR.

##### Malaysia

Fig. 1 (D) shows our results for the 15 provinces in Malaysia. While the country-specific ensemble achieved the most top 3 rankings at the 1-month and 3-month ahead horizons, clustered + regularized VAR achieved the most top 3 rankings at the 2-month horizon. Notably, the overall ensemble did not generalize very well in Malaysia, being ranked even below the naive persistence baseline at the 1-month ahead horizon. We note, however, that the two easternmost (and largest) Malaysian provinces both saw the country-specific ensemble performing the best out of all 13 models at the 1-month and 3-month horizons.

##### Mexico

Fig. 1(E) shows our 13 models’ rankings in the 32 provinces of Mexico. Regularized VAR achieved the top 3 rankings at the 1-month ahead forecast horizon, followed by the country-specific ensemble. At the 2-month ahead horizon, the two ensembles achieved the top 3 rankings, though at 3-months ahead. In contrast, the overall ensemble maintained the top 3 rankings, regularized VAR overtook the country-specific ensemble for the second most top 3 rankings. From the geographical maps, we see that no individual model achieved the top rank across most of locations.

##### Thailand

In Fig. 1(F), we show the results in the 77 provinces of Thailand. From the heatmaps, we observed that the country-specific ensemble achieved the most top 3 rankings at the 1-month and 2-month horizons. By comparison, regularized VAR superseded both ensembles for the most top 3 rankings at the 3-month horizon. Notably, at the 2-month horizon, regularized VAR achieved more first-place rankings than both ensembles. At the 1-month horizon, regularized VAR achieved the same number of first-place rankings as the overall ensemble and significantly more first-place rankings than the country-specific ensemble. The geographical maps show that no particular model performed the best across most Thailand provinces.

### 3.2 Forecasting performance of ensemble models built from standard non-optimized individual components

It is not always feasible to fine-tune the hyperparameters of each individual component model into their most optimized, highest-performing variants, as we did in this study. In many situations, the lack of high-quality epidemiological data and/or computational resources may impose significant challenges, likely forcing users to apply standard, off-the-shelf models and systems without significant fine-tuning. In this section, we demonstrate that even in such a situation, ensembling is a very effective and robust solution for short-term forecasting.

We present the performances of 11 standard component models and two ensemble variants on forecasting dengue in our tested province-level locations. The forecasting tasks, evaluation ranges and metrics, data sources, and training processes are identical to those of the results that we present in the main manuscript for the optimized models. We also refer the reader to our Methods section and the Additional Methods Details in our Supplementary Information for specific details on our standard individual component models, intended to replicate off-the-shelf deployment.

Our key findings from this section not only corroborate but also, in fact, enhance our reported findings in the main manuscript. First, we find that there does not exist one standard model that consistently outperforms all others in all locations, reinforcing our corresponding finding in the main manuscript with optimized models. Second, and most importantly, even though our standard individual component models are overall markedly inferior to the naive persistence baseline model, combining such weak individual component models together produces ensemble models that consistently and significantly outperform the naive persistence baseline. Indeed, even when given weak individual learners, our ensemble methods are robust and generalizable forecasting tools. Fig. 11 summarizes our standard individual and ensemble models’ comparative performances within each of the five tested countries, across our three prediction tasks of forecasting 1-, 2-, and 3-months ahead. The individual models presented here are deployed with standard off-the-shelf settings and are not fully-optimized.

**Figure 11:**
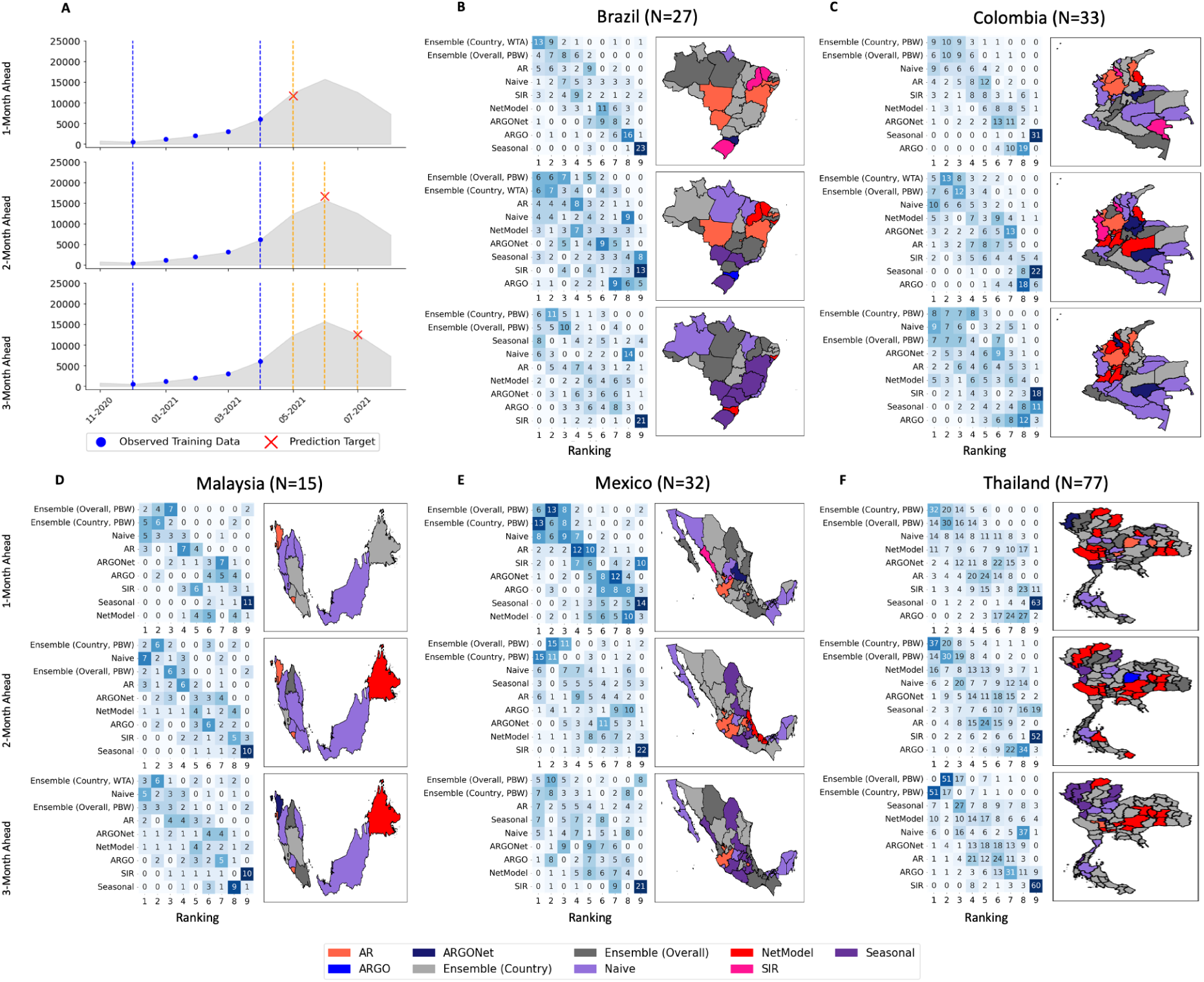
Country-specific standard individual and ensemble model performance rankings. **(A)** Graphical representations of the 1-, 2-, and 3-month forecast horizons. The red *X* marks our forecasting target *n*-months ahead, the blue dots represent the historical cases that we are using as our observed training data (in this case, a 5-month window), the vertical blue dotted lines represent the limits of our training data range. The grey silhouette represents the ground truth reported case counts. **(B) - (F)** Within each country and forecast horizon, the heatmaps show the rankings distribution for each individual and ensemble model’s forecasts in terms of percent absolute error. The geographic maps next to each heatmap indicate the best-performing model in each province, color-coded by the legend at the bottom of the figure.

The heatmaps on Fig. 11 represent the distribution of rankings for each model in each country, with models listed in decreasing order by the number of top 3 rankings accrued. We observe that in all combinations of country and forecast horizon presented, an ensemble model achieved the most top 3 rankings. This corroborates our finding in the main manuscript that the ensembles, while not always the top 1 ranked model, are generally very high-performing and robust.

From the geographical color-coded maps encoding the top 1 model in each province, we observe that no one model — standard individual nor ensemble — consistently outperformed the rest of the models. This result mirrors our finding in the main manuscript that no individual model performs the best across all locations consistently. Finally, the heatmaps and geographical maps emphasize that our standard individual component models are indeed very weak learners, being mostly outperformed by the naive persistence model.

Fig. 12 shows the forecasting performances of our standard individual and ensemble models across all 187 tested locations. Panel (A) emphasizes the primary advantage of ensemble models over their individual component models: while the individual component models fluctuate wildly in underpredicting and overpredicting, the ensemble model is much more invariant to such fluctuations and much more closely matches the ground truth.

From panel (B), we observe that for all three forecast horizons, both ensemble models achieved the most top 3 rankings compared to any other model, including the naive persistence baseline, which is ranked higher than all other standard individual component models. Indeed, one main advantage of the ensemble models is that they can take in the naive persistence models’ inputs as a component model, absorbing the robustness of the naive persistence model in times when the more complex data-driven models fail to perform well.

Panel (C) corroborates the main message in panel (B): the presence of only a few sparse patches of grey indicates that the ensemble models performed in the top 3 rankings for almost all tested locations. It should be mentioned that there are more yellow patches on this grid of maps than the corresponding grid presented in the main manuscript (see Fig. 2) — which is to be expected, as the standard individual models are consistently weaker than their optimized counterparts.

Fig. 13 displays the percent absolute error distributions for all of our standard individual component and ensemble models across all 187 tested locations. The ensemble models incurred the best mean percent absolute error across all locations compared to all other models. If compared to the optimized model results in the main manuscript, the improvements from individual component models to ensemble models were much larger when working with standard models. Finally, while all of the individual component models, at any forecast horizon, incurred significantly worse errors than the naive persistence model, the resultant ensemble models — taking as input these very same weak learners — were generally able to outperform the naive persistence baseline.

**Figure 12:**
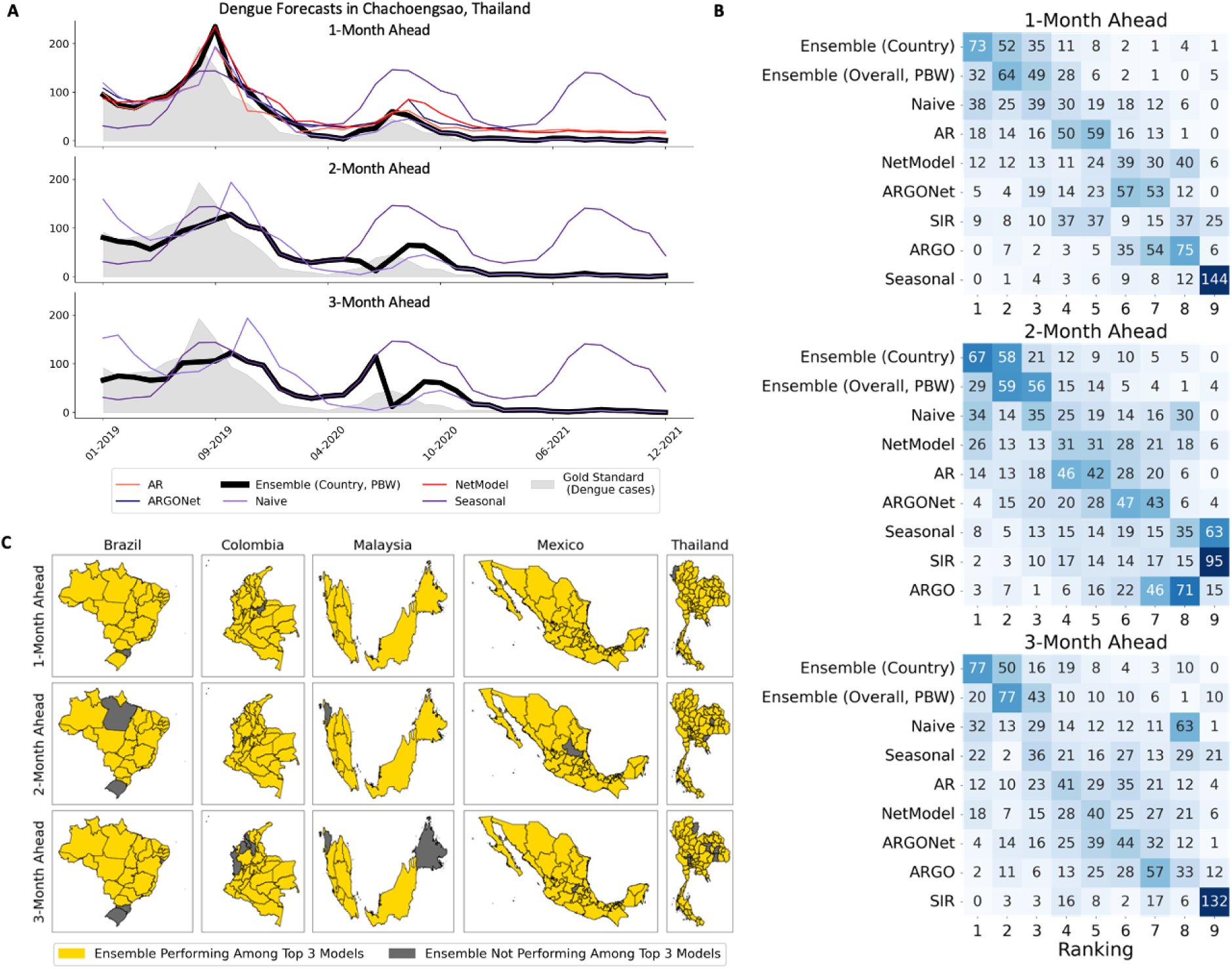
A summary of our prediction tasks and standard models’ overall performances across 187 locations. **(A)** An example of our standard country-specific ensemble variants’ forecasts compared to their standard, non-optimized component models in one selected location — Chachoengsao, Thailand. The gold standard ground truth of reported dengue cases is shown as the grey silhouette. Ensemble predictions are shown in thick, bolded lines, while standard component models are shown in thinner, colored lines.**(B)** Heatmaps of the number of locations where each model attained a specific ranking in terms of mean absolute error with respect to the ground truth reported dengue case counts across all 187 locations. **(C)** Geographical maps of Brazil, Colombia, Malaysia, Mexico, and Thailand showing provinces where either the country-specific or overall ensemble performed in the top 3 rankings for each location (in yellow) and where they did not (in grey).

**Figure 13:**
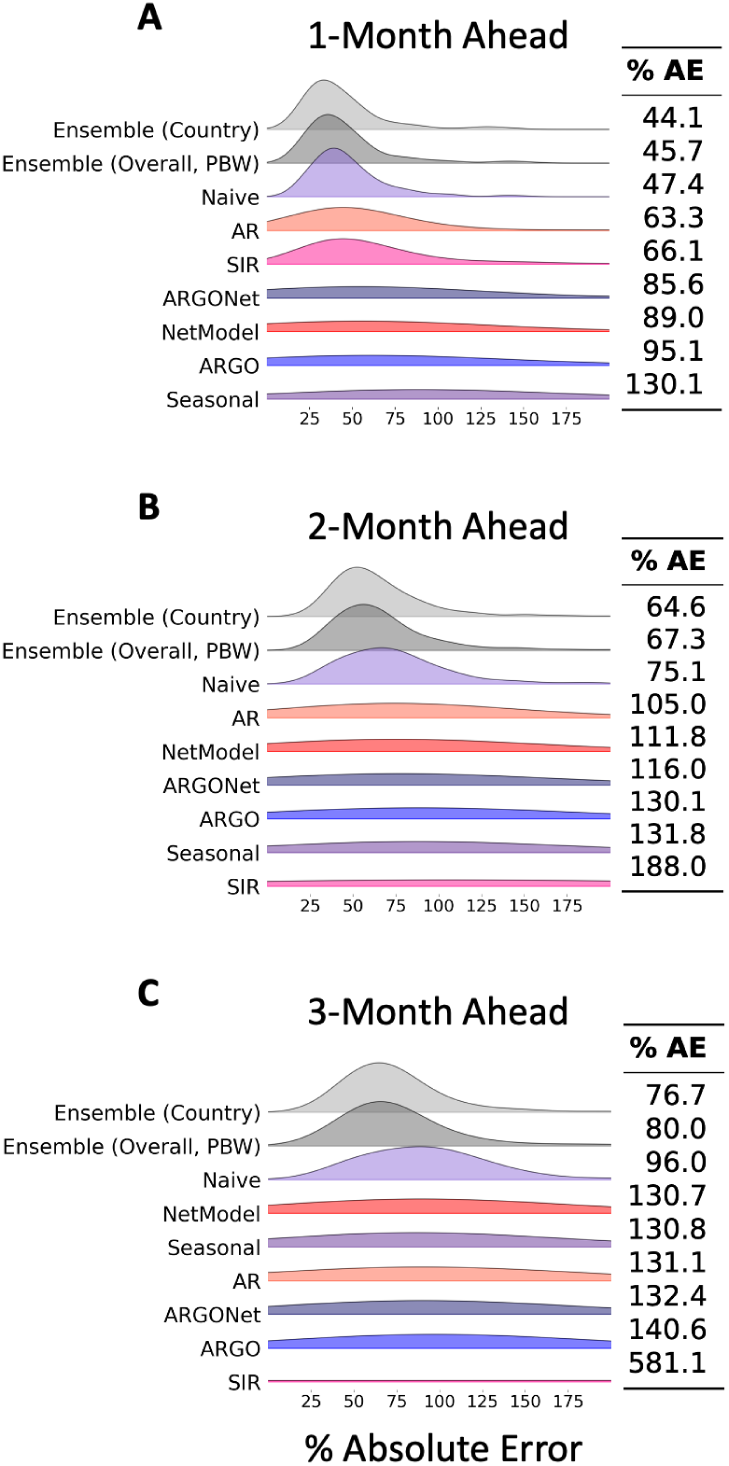
Overall error distributions for standard individual and ensemble models across all 187 tested locations. **(A) - (C)** Ridgeline plots show percent absolute error distributions at the 1-month, 2-month, and 3-month horizons, respectively. Side tables record the mean percent absolute error incurred.

Fig. 14 provides more granular, country-specific summaries of the standard individual and component models’ percent absolute error distributions within each country and forecast horizon. Corroborating our findings in the main manuscript, an ensemble model achieved the lowest mean percent absolute error in 12 out of 15 tested combinations of forecast horizon and country. The only exceptions were Malaysia at 1- and 2-months ahead, and Mexico at 1-month ahead, where the ensembles were still unable to outperform the naive persistence model in terms of mean percent absolute error. Nonetheless, even when the naive persistence model ranks higher, the error distributions of the two ensembles are visually very similar to that of the naive persistence.

**Figure 14:**
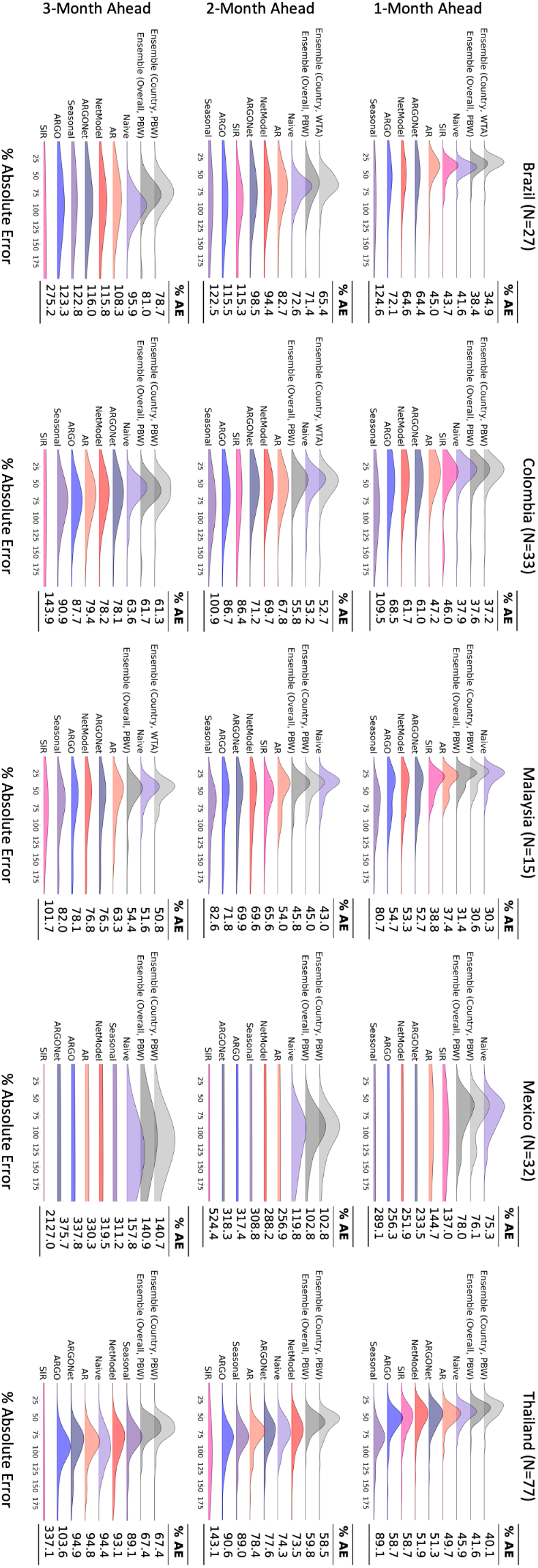
Country-specific error distributions for standard individual and ensemble models, organized by country and forecast horizon. Ridgeline plots show percent absolute error distributions at the 1-month, 2-month, and 3-month horizons. Side tables record the mean per-

Compared to Fig. 4, we observe that the differences in variance between the two ensembles and the standard individual component models, as measured through the spread of their ridgeline distributions, were much more pronounced. This corroborates our continual finding that ensembles yield relatively larger performance improvements when provided with weaker learners as input.

### 3.3 Additional Methods Details

**Figure 15:**
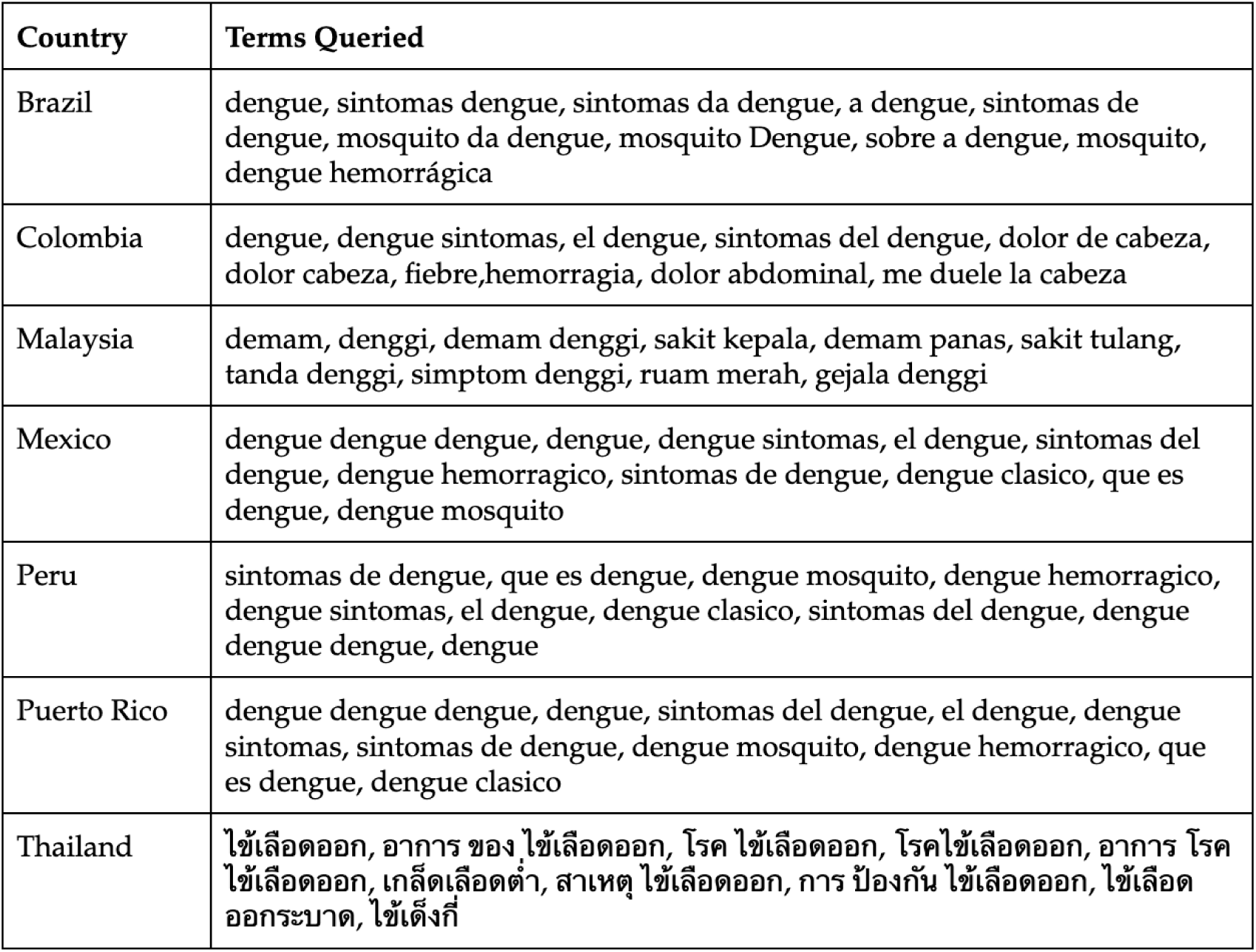
Country-specific Google Trends search terms.

### 3.4 Optimized Models’ PAE by Country

#### 3.4.1 1-Month Ahead

**Table 9:**
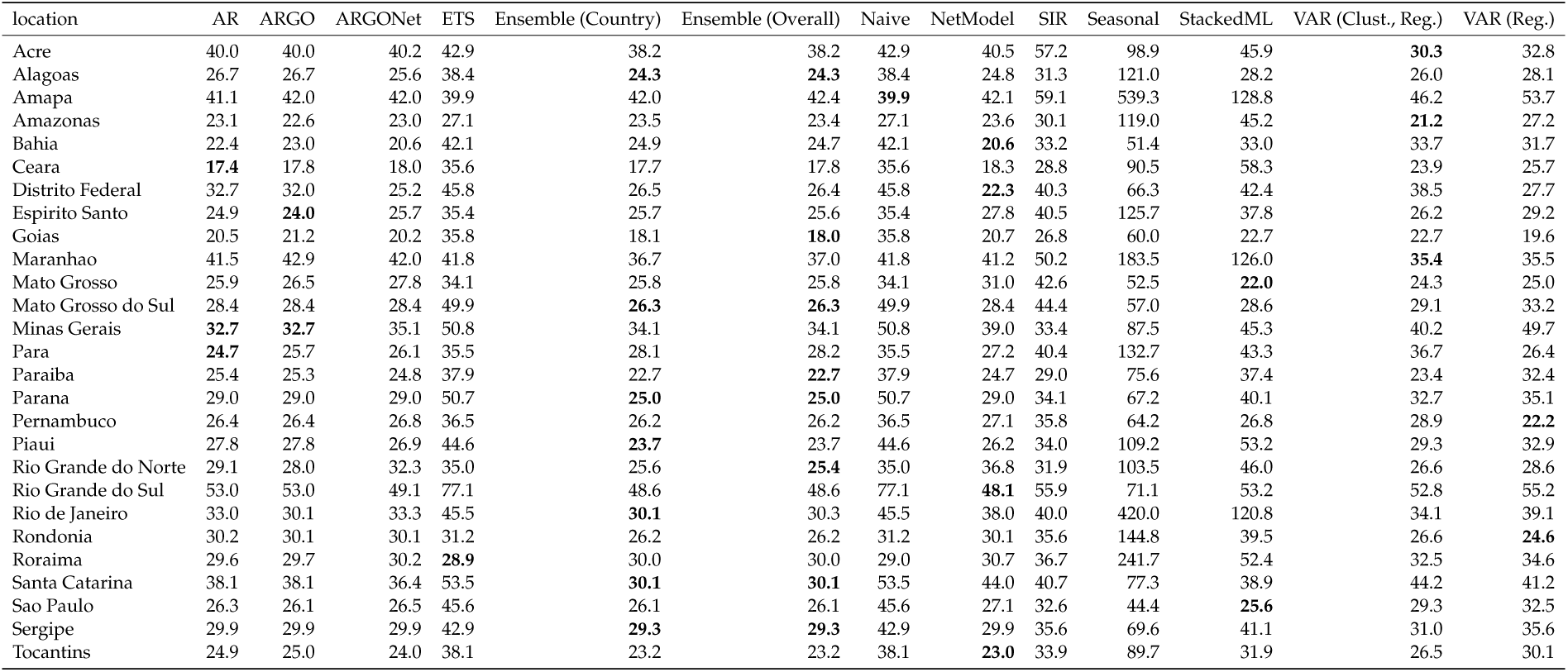
1-month ahead percent absolute errors in Brazil for optimized models. Best performing models in each location are bolded. Lower values indicate stronger performance.

**Table 10:**
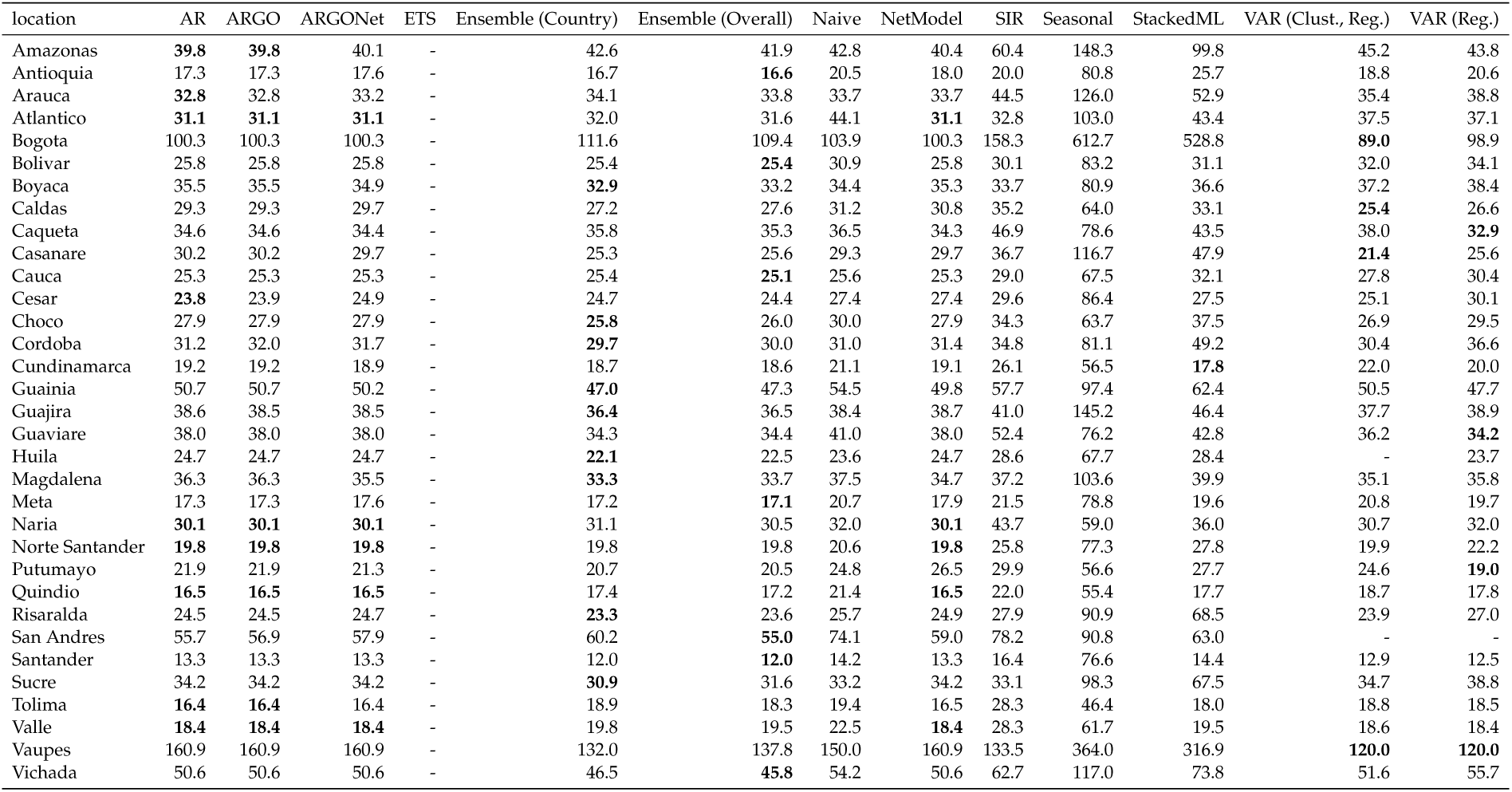
1-month ahead percent absolute errors in Colombia for optimized models. Best performing models in each location are bolded. Lower values indicate stronger performance.

**Table 11:** 1-month ahead percent absolute errors in Malaysia for optimized models. Best performing models in each location are bolded. Lower values indicate stronger performance.

| location | AR | ARGO | ARGONet | ETS | Ensemble (Country) | Ensemble (Overall) | Naive | NetModel | SIR | Seasonal | StackedML | VAR (Clust., Reg.) | VAR (Reg.) |
| --- | --- | --- | --- | --- | --- | --- | --- | --- | --- | --- | --- | --- | --- |
| Johor | 24.3 | 24.3 | 24.3 | 24.2 | 21.5 | 22.4 | 21.8 | 24.3 | 28.4 | 52.6 | 29.9 | 22.0 | **21.4** |
| Kedah | 26.0 | 26.0 | 26.0 | 29.7 | 25.8 | 24.0 | 28.2 | 26.0 | 36.4 | 49.9 | 36.9 | **23.5** | 24.4 |
| Kelantan | 36.7 | 36.7 | 36.1 | 41.8 | **33.2** | 38.1 | 41.8 | 35.6 | 51.1 | 82.8 | 59.8 | 40.4 | 40.1 |
| Kuala Lumpur and Putrajaya | 24.9 | 24.9 | 24.9 | 25.1 | **21.8** | 21.9 | 23.4 | 24.9 | 28.3 | 58.9 | 29.8 | 22.8 | 22.3 |
| Labuan | 56.8 | 56.8 | 56.8 | **53.9** | 58.6 | 61.2 | 65.8 | 56.8 | 71.9 | 109.9 | 103.4 | 60.5 | 66.0 |
| Malacca | - | - | - | 25.0 | 20.8 | 20.5 | 21.0 | - | - | 54.8 | 24.7 | 21.7 | **19.4** |
| Negeri Sembilan | 21.3 | 21.3 | 21.3 | 21.9 | **19.9** | 21.1 | 20.4 | 21.3 | 24.9 | 47.6 | 25.1 | 26.1 | 25.0 |
| Pahang | 25.1 | 25.0 | 24.8 | 25.9 | **19.7** | 21.7 | 20.3 | 24.6 | 30.0 | 64.3 | 29.0 | 23.3 | 23.8 |
| Perak | 22.9 | 22.9 | 22.9 | 20.4 | 19.9 | 21.0 | **19.4** | 22.9 | 23.2 | 103.3 | 34.1 | 23.8 | 24.1 |
| Perlis | 44.7 | 44.7 | 44.7 | 44.5 | 46.9 | 35.6 | 52.5 | 44.7 | 66.1 | 116.8 | 55.3 | **31.6** | 34.1 |
| Pulau Pinang | - | - | - | 28.8 | **21.3** | 28.7 | 27.4 | - | - | 106.3 | 33.4 | 29.1 | 28.2 |
| Sabah | 32.0 | 32.0 | 32.0 | 31.8 | **27.5** | 34.4 | 31.6 | 31.9 | 53.4 | 48.7 | 34.0 | 31.0 | 30.9 |
| Sarawak | 24.8 | 24.8 | 24.8 | 22.6 | **19.5** | 23.6 | 20.6 | 24.9 | 27.6 | 85.0 | 26.8 | 28.7 | 29.0 |
| Selangor | 24.4 | 24.4 | 24.4 | 23.2 | 21.7 | 22.7 | **20.8** | 24.4 | 32.5 | 57.6 | 31.2 | 22.4 | 21.6 |
| Terengganu | 38.5 | 38.5 | 38.5 | 38.8 | 41.4 | 37.7 | 39.0 | 38.5 | 45.2 | 172.5 | 64.1 | 38.3 | 38.0 |

**Table 12:** 1-month ahead percent absolute errors in Mexico for optimized models. Best performing models in each location are bolded. Lower values indicate stronger performance.

| location | AR | ARGO | ARGONet | ETS | Ensemble (Country) | Ensemble (Overall) | Naive | NetModel | SIR | Seasonal | StackedML | VAR (Clust., Reg.) | VAR (Reg.) |
| --- | --- | --- | --- | --- | --- | --- | --- | --- | --- | --- | --- | --- | --- |
| Aguascalientes | 103.2 | 103.2 | 103.2 | 97.6 | **84.7** | 95.4 | 100.8 | 103.3 | 88.1 | 153.9 | 129.4 | 94.1 | 88.2 |
| Baja California | 212.6 | 212.6 | 212.6 | **111.1** | 164.2 | 179.1 | 111.1 | 212.6 | 192.3 | 1911.7 | 1622.6 | 148.1 | 166.7 |
| Baja California Sur | 68.5 | 68.5 | 60.1 | 79.4 | 76.3 | 81.9 | 78.8 | **54.3** | 135.3 | 2126.3 | 1606.1 | 89.0 | 77.3 |
| Campeche | 68.3 | 68.3 | 71.0 | 70.8 | 66.7 | 68.5 | 74.2 | 73.8 | 91.8 | 418.7 | 101.5 | **60.7** | 61.3 |
| Chiapas | 46.8 | 46.8 | 46.9 | 49.6 | 39.2 | 45.3 | 49.7 | 48.5 | 71.7 | 139.8 | 68.3 | **36.5** | 44.5 |
| Chihuahua | 110.0 | 110.0 | 110.0 | 137.2 | 111.1 | 132.2 | 140.7 | 110.0 | 253.6 | 137.3 | 132.6 | **89.6** | 102.1 |
| Coahuila | 56.0 | 58.1 | 59.0 | 90.6 | **48.0** | 58.0 | 90.6 | 60.0 | 64.0 | 76.8 | 79.1 | 70.9 | 72.0 |
| Colima | 54.2 | 54.2 | 54.2 | 62.0 | 51.9 | 52.1 | 62.0 | 54.2 | 76.4 | 195.3 | 71.5 | 50.3 | **48.2** |
| Durango | 100.0 | 100.0 | 100.0 | 106.0 | 118.1 | 102.2 | 98.8 | 100.0 | 125.9 | 282.8 | 324.7 | **78.6** | 130.5 |
| Guanajuato | 71.2 | 71.2 | 71.1 | 93.7 | **55.1** | 63.1 | 86.4 | 71.1 | 98.0 | 308.3 | 398.6 | 66.8 | 62.2 |
| Guerrero | 51.7 | 51.7 | 51.9 | 56.0 | 53.0 | **44.2** | 56.0 | 52.1 | 56.0 | 151.6 | 90.2 | 47.6 | 55.9 |
| Hidalgo | 75.4 | 75.4 | 75.6 | 87.2 | 60.8 | 67.5 | 87.2 | 75.8 | 112.6 | 96.6 | 91.0 | 70.9 | **59.9** |
| Jalisco | 44.5 | 40.1 | 35.8 | 55.2 | 33.8 | **32.0** | 55.2 | 44.8 | 67.3 | 88.1 | 58.4 | 44.6 | 38.7 |
| Mexico | 117.9 | 118.1 | 118.0 | 118.7 | 119.5 | 115.7 | 136.3 | 117.9 | 185.1 | 145.6 | 173.5 | **115.5** | 117.3 |
| Mexico City | - | - | - | - | - | - | - | - | - | - | - | - | - |
| Michoacan | 42.2 | 31.9 | 36.9 | 39.2 | **26.3** | 35.9 | 39.2 | 44.0 | 50.3 | 77.9 | 50.8 | 40.7 | 42.2 |
| Morelos | 48.7 | 47.6 | 48.3 | 61.2 | **46.9** | 48.7 | 61.3 | 49.2 | 73.6 | 62.6 | 48.2 | 47.0 | 46.9 |
| Nayarit | 62.1 | 62.1 | 62.1 | 64.0 | 49.8 | 57.1 | 64.3 | 62.1 | 80.3 | 95.3 | 73.0 | 48.5 | **47.7** |
| Nuevo Leon | 48.5 | 47.8 | 45.0 | 87.8 | 50.9 | 60.8 | 87.4 | **44.4** | 110.6 | 122.8 | 93.7 | 51.2 | 67.5 |
| Oaxaca | 54.3 | 54.3 | 53.3 | 58.8 | 47.9 | **44.9** | 58.7 | 53.2 | 83.4 | 100.4 | 61.1 | 54.1 | 50.6 |
| Puebla | 52.9 | 52.7 | 51.1 | 58.0 | 48.7 | **45.7** | 58.1 | 50.6 | 57.8 | 78.2 | 94.9 | 53.4 | 45.9 |
| Queretaro | 73.6 | 73.6 | 82.9 | 76.5 | 76.7 | **73.6** | 76.0 | 92.3 | 118.0 | 110.4 | 110.7 | 81.9 | 75.9 |
| Quintana Roo | 45.5 | 45.5 | 45.5 | 41.6 | 47.0 | 42.7 | **41.6** | 45.5 | 46.0 | 142.1 | 67.4 | 52.2 | 42.5 |
| San Luis Potosi | 62.9 | 62.9 | 62.9 | 78.8 | **54.5** | 59.5 | 78.7 | 62.9 | 100.2 | 90.4 | 93.5 | 56.7 | 56.5 |
| Sinaloa | 54.2 | 53.6 | 54.0 | 63.7 | 49.7 | 50.0 | 63.7 | 54.6 | **32.2** | 89.0 | 57.8 | 67.1 | 70.6 |
| Sonora | 53.9 | 53.9 | 54.6 | 72.6 | 58.6 | 54.0 | 72.5 | 55.2 | 75.7 | 507.9 | 412.1 | 63.5 | **52.6** |
| Tabasco | 53.1 | 53.1 | 53.2 | 54.2 | 52.5 | 53.1 | 54.2 | 53.3 | 66.6 | 233.6 | 57.8 | **50.4** | 51.1 |
| Tamaulipas | 63.4 | 63.4 | 63.4 | 62.8 | 58.6 | **50.8** | 62.8 | 63.5 | 68.9 | 147.1 | 132.1 | 55.8 | 55.1 |
| Tlaxcala | - | - | - | - | - | - | - | - | - | - | - | - | - |
| Veracruz | 48.9 | 48.4 | 47.1 | 52.1 | 44.4 | 43.3 | 52.0 | 45.9 | 49.6 | 104.6 | 73.2 | 49.5 | **36.6** |
| Yucatan | 61.4 | 61.4 | 58.9 | 53.1 | 52.7 | 53.2 | 53.2 | 56.6 | 51.1 | 285.2 | 93.3 | 60.9 | **42.7** |
| Zacatecas | 113.5 | 113.5 | 113.7 | 105.9 | 102.3 | **96.2** | 107.8 | 113.8 | 214.1 | 192.2 | 206.0 | 103.7 | 109.7 |

**Table 13:** 1-month ahead percent absolute errors in Peru for optimized models. Best performing models in each location are bolded. Lower values indicate stronger performance.

| location | AR | ARGO | ARGONet | ETS | Ensemble (Country) | Ensemble (Overall) | Naive | NetModel | SIR | Seasonal | StackedML | VAR (Clust., Reg.) | VAR (Reg.) |
| --- | --- | --- | --- | --- | --- | --- | --- | --- | --- | --- | --- | --- | --- |
| Iquitos | 67.5 | 67.7 | 67.6 | 79.8 | **53.6** | - | 79.2 | 67.5 | 119.3 | 80.6 | 74.4 | - | - |

**Table 14:** 1-month ahead percent absolute errors in Puerto Rico for optimized models. Best performing models in each location are bolded. Lower values indicate stronger performance.

| location | AR | ARGO | ARGONet | ETS | Ensemble (Country) | Ensemble (Overall) | Naive | NetModel | SIR | Seasonal | StackedML | VAR (Clust., Reg.) | VAR (Reg.) |
| --- | --- | --- | --- | --- | --- | --- | --- | --- | --- | --- | --- | --- | --- |
| San Juan | 27.4 | 28.4 | 27.9 | 30.7 | **16.8** | - | 30.7 | 27.4 | 22.2 | 60.7 | 22.5 | - | - |

**Table 15:**
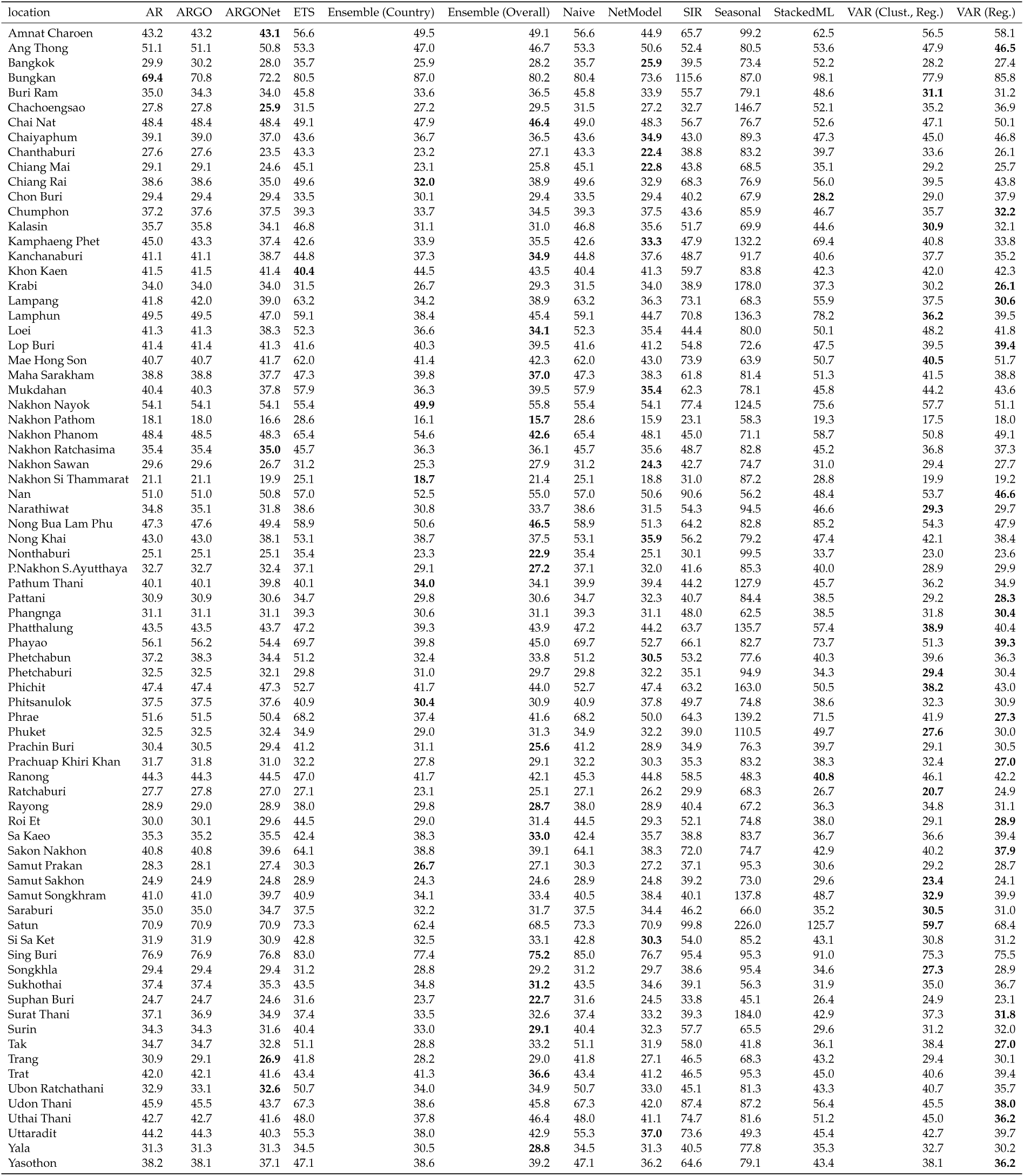
1-month ahead percent absolute errors in Thailand for optimized models. Best performing models in each location are bolded. Lower values indicate stronger performance.

#### 3.4.2 2-Month Ahead

**Table 16:**
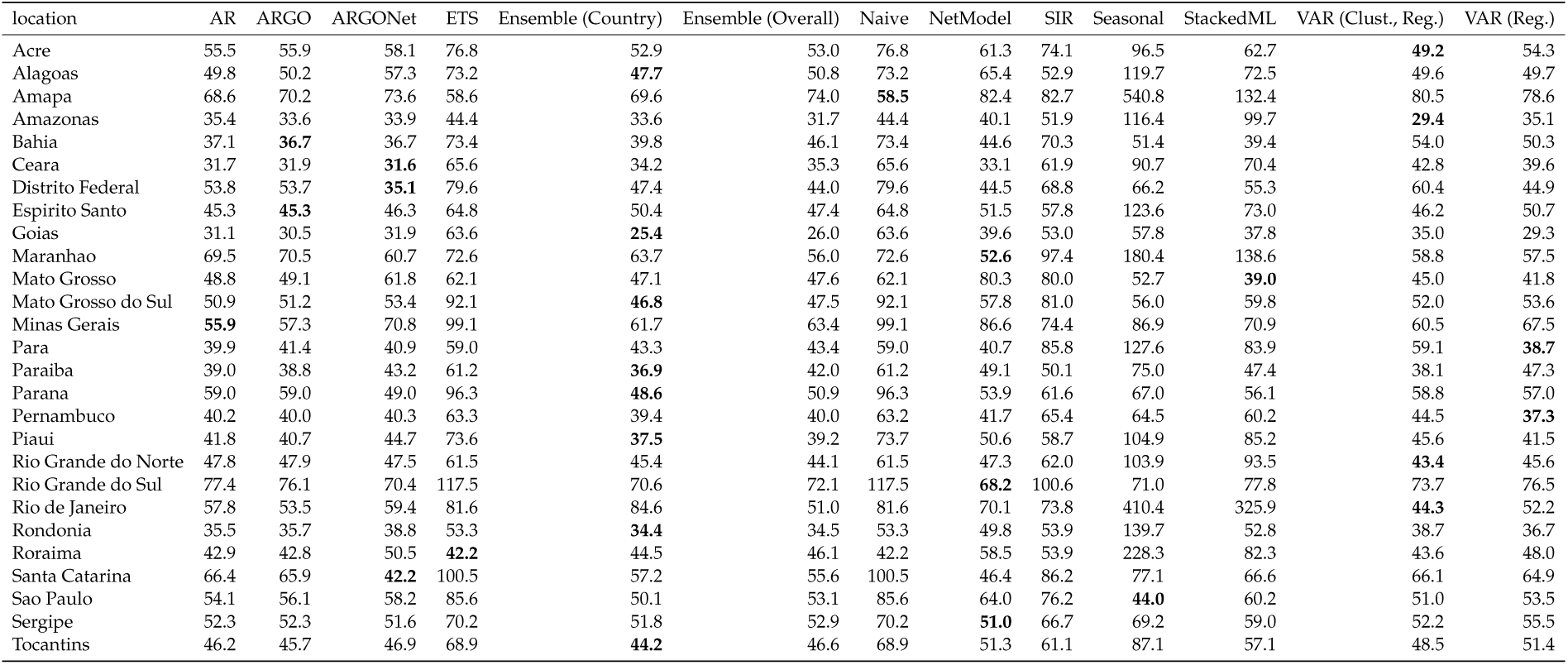
2-month ahead percent absolute errors in Brazil for optimized models. Best performing models in each location are bolded. Lower values indicate stronger performance.

**Table 17:** 2-month ahead percent absolute errors in Colombia for optimized models. Best performing models in each location are bolded. Lower values indicate stronger performance.

| location | AR | ARGO | ARGONet | ETS | Ensemble (Country) | Ensemble (Overall) | Naive | NetModel | SIR | Seasonal | StackedML | VAR (Clust., Reg.) | VAR (Reg.) |
| --- | --- | --- | --- | --- | --- | --- | --- | --- | --- | --- | --- | --- | --- |
| Amazonas | **49.4** | **49.4** | 58.1 | - | 51.3 | 52.9 | 55.0 | 69.4 | 68.1 | 119.2 | 77.6 | 50.3 | 50.6 |
| Antioquia | 32.9 | 32.9 | 37.7 | - | **28.5** | 33.9 | 37.5 | 51.2 | 32.9 | 76.3 | 40.5 | 32.0 | 35.7 |
| Arauca | 42.8 | 42.8 | **41.6** | - | 48.4 | 45.4 | 46.4 | 41.9 | 71.0 | 128.5 | 88.7 | 45.9 | 51.2 |
| Atlantico | 53.4 | 53.6 | 54.1 | - | **51.6** | 56.8 | 74.4 | 55.1 | 68.5 | 90.7 | 55.6 | 62.3 | 55.1 |
| Bogota | 108.6 | 111.4 | 118.8 | - | 115.2 | 103.5 | 96.1 | 126.2 | 163.8 | 431.6 | 368.3 | **87.1** | 104.5 |
| Bolivar | 45.3 | 46.3 | 49.6 | - | **41.2** | 48.8 | 50.9 | 54.5 | 47.2 | 79.2 | 45.8 | 48.0 | 49.1 |
| Boyaca | 46.8 | 46.5 | 46.8 | - | 43.0 | 45.9 | 45.9 | 51.5 | 54.7 | 78.0 | **42.7** | 47.3 | 48.3 |
| Caldas | 36.0 | 36.0 | 31.6 | - | 31.0 | **29.0** | 39.6 | 30.5 | 45.9 | 59.9 | 41.1 | 29.2 | 30.1 |
| Caqueta | 51.8 | 51.8 | **36.8** | - | 47.1 | 42.2 | 59.4 | 37.8 | 83.1 | 75.8 | 71.9 | 52.4 | 42.3 |
| Casanare | 53.0 | 53.0 | 40.5 | - | 37.0 | 33.5 | 46.6 | 36.8 | 64.4 | 134.3 | 57.5 | **30.2** | 37.5 |
| Cauca | **38.1** | **38.1** | 42.2 | - | 41.6 | 44.0 | 39.8 | 47.5 | 54.8 | 65.9 | 42.1 | 42.2 | 48.2 |
| Cesar | 41.2 | 41.2 | 45.8 | - | 42.2 | 44.3 | 44.7 | 53.9 | 57.8 | 84.6 | 50.5 | **39.1** | 48.7 |
| Choco | 40.9 | 40.9 | 40.4 | - | 36.0 | 36.3 | 42.3 | 39.9 | 52.1 | 61.5 | 44.3 | **33.9** | 39.1 |
| Cordoba | 46.3 | 51.5 | 49.9 | - | **43.0** | 47.9 | 49.2 | 52.5 | 46.2 | 77.7 | 118.5 | 43.7 | 51.5 |
| Cundinamarca | 32.1 | 32.1 | 27.9 | - | 28.4 | 29.5 | 35.7 | **27.8** | 42.6 | 50.0 | 32.7 | 33.2 | 29.7 |
| Guainia | 60.6 | 60.6 | **59.7** | - | 63.0 | 61.9 | 82.7 | 62.2 | 86.0 | 97.6 | 84.4 | 66.5 | 62.6 |
| Guajira | 57.2 | 56.6 | 58.3 | - | **54.9** | 58.8 | 58.4 | 62.4 | 67.9 | 129.4 | 97.0 | 59.1 | 59.4 |
| Guaviare | 52.9 | 52.9 | 50.0 | - | **45.9** | 48.4 | 59.3 | 51.7 | 70.8 | 73.3 | 50.0 | 50.8 | 48.1 |
| Huila | 41.0 | 43.5 | 38.9 | - | 37.9 | 37.6 | 40.0 | **35.9** | 55.1 | 66.4 | 44.1 | - | 37.8 |
| Magdalena | 58.9 | 58.9 | 59.3 | - | **51.9** | 55.5 | 62.0 | 60.9 | 66.1 | 103.3 | 63.5 | 55.2 | 54.7 |
| Meta | 32.0 | 32.0 | 27.7 | - | 26.9 | 29.8 | 34.6 | **24.9** | 38.8 | 75.7 | 44.4 | 33.8 | 29.9 |
| Naria | 36.3 | 36.3 | 33.2 | - | 36.7 | **31.7** | 40.9 | 38.6 | 58.4 | 58.9 | 59.2 | 32.9 | 33.4 |
| Norte Santander | 32.1 | 32.1 | 32.8 | - | **29.7** | 31.6 | 32.1 | 34.6 | 44.1 | 71.8 | 36.7 | 29.9 | 32.9 |
| Putumayo | 40.7 | 40.7 | 33.5 | - | 31.7 | 32.1 | 43.1 | 32.2 | 52.9 | 58.1 | 50.5 | 39.1 | **26.3** |
| Quindio | 29.9 | 29.6 | 29.8 | - | 28.8 | 28.7 | 35.2 | 30.7 | 38.9 | 53.2 | 43.8 | 30.4 | **27.0** |
| Risaralda | 38.2 | 37.4 | 43.5 | - | **37.3** | 40.2 | 42.8 | 49.7 | 44.2 | 86.0 | 67.5 | 37.9 | 39.9 |
| San Andres | 67.1 | 67.9 | 67.3 | - | 89.9 | 67.3 | 110.4 | **66.8** | 125.2 | 90.8 | 91.2 | - | - |
| Santander | 22.6 | 22.6 | 21.0 | - | **18.2** | 19.5 | 22.3 | 21.4 | 25.7 | 72.6 | 25.8 | 19.4 | 18.9 |
| Sucre | 50.8 | 51.0 | 50.6 | - | **41.7** | 49.1 | 51.4 | 51.8 | 47.9 | 82.5 | 67.7 | 46.9 | 50.2 |
| Tolima | 28.8 | 28.8 | 28.0 | - | 29.2 | **27.1** | 30.8 | 28.1 | 50.8 | 41.7 | 32.4 | 27.8 | 27.3 |
| Valle | 33.1 | 33.1 | 32.1 | - | 34.0 | 32.4 | 37.6 | 36.2 | 54.8 | 61.8 | 37.2 | 34.5 | **31.3** |
| Vaupes | 176.2 | 176.2 | 160.5 | - | 147.1 | 145.3 | 141.7 | 171.6 | 168.0 | 381.9 | 360.5 | **133.8** | 141.7 |
| Vichada | 61.0 | 61.0 | 60.6 | - | 61.9 | 59.1 | 66.8 | 60.3 | 99.7 | 109.9 | 102.0 | **55.0** | 67.0 |

**Table 18:** 2-month ahead percent absolute errors in Malaysia for optimized models. Best performing models in each location are bolded. Lower values indicate stronger performance.

| location | AR | ARGO | ARGONet | ETS | Ensemble (Country) | Ensemble (Overall) | Naive | NetModel | SIR | Seasonal | StackedML | VAR (Clust., Reg.) | VAR (Reg.) |
| --- | --- | --- | --- | --- | --- | --- | --- | --- | --- | --- | --- | --- | --- |
| Johor | 36.0 | 36.0 | 36.1 | 37.9 | 36.8 | 34.6 | 38.0 | 36.2 | 51.7 | 53.1 | **32.6** | 33.8 | 34.0 |
| Kedah | 36.3 | 35.8 | 35.3 | 41.5 | 35.9 | 34.0 | 42.3 | 34.9 | 60.6 | 53.9 | 35.8 | **33.8** | 34.7 |
| Kelantan | 51.6 | 50.8 | 50.1 | 62.6 | 54.9 | 54.1 | 62.6 | **49.9** | 93.7 | 82.2 | 68.1 | 56.7 | 59.5 |
| Kuala Lumpur and Putrajaya | 35.7 | 35.8 | 35.5 | 38.0 | 34.5 | 32.7 | 35.8 | 35.2 | 51.0 | 59.1 | 36.0 | **32.5** | 32.9 |
| Labuan | 73.6 | 74.1 | 76.0 | **66.4** | 77.7 | 78.0 | 84.8 | 78.9 | 106.5 | 151.3 | 148.7 | 82.7 | 78.8 |
| Malacca | - | - | - | 37.3 | 34.1 | 31.3 | 34.6 | - | - | 56.8 | 36.8 | 31.4 | **31.3** |
| Negeri Sembilan | 29.1 | 29.1 | 29.0 | 30.1 | **28.9** | 29.3 | 30.0 | 29.0 | 38.2 | 48.4 | 32.8 | 29.4 | 30.8 |
| Pahang | 33.9 | 33.8 | 33.1 | 37.8 | 32.9 | 34.7 | 35.9 | **32.5** | 50.9 | 62.7 | 40.0 | 36.0 | 36.1 |
| Perak | 31.2 | 31.2 | 30.8 | 30.1 | 30.1 | 35.4 | **29.1** | 30.3 | 39.5 | 95.1 | 52.4 | 37.5 | 39.7 |
| Perlis | 62.7 | 62.7 | 61.6 | 62.9 | **47.7** | 53.5 | 65.7 | 60.7 | 90.2 | 122.4 | 96.7 | 51.5 | 51.4 |
| Pulau Pinang | - | - | - | 38.6 | 36.1 | 36.9 | **35.5** | - | - | 95.0 | 46.3 | 36.6 | 37.1 |
| Sabah | 36.7 | 36.4 | 37.9 | 40.3 | 40.6 | 37.6 | 42.6 | 39.5 | 73.5 | 48.8 | **35.6** | 38.0 | 37.5 |
| Sarawak | 33.1 | 33.1 | 34.1 | 31.2 | 30.3 | 37.3 | **27.3** | 35.9 | 43.8 | 81.7 | 39.8 | 39.7 | 39.2 |
| Selangor | 33.1 | 32.7 | 33.1 | 34.3 | 33.2 | 32.6 | 34.0 | 33.4 | 50.4 | 57.5 | 34.6 | **32.1** | 33.0 |
| Terengganu | 48.3 | 48.5 | 47.3 | **45.6** | 46.7 | 52.1 | 47.1 | 46.4 | 57.3 | 171.2 | 83.2 | 54.2 | 55.0 |

**Table 19:** 2-month ahead percent absolute errors in Mexico for optimized models. Best performing models in each location are bolded. Lower values indicate stronger performance.

| location | AR | ARGO | ARGONet | ETS | Ensemble (Country) | Ensemble (Overall) | Naive | NetModel | SIR | Seasonal | StackedML | VAR (Clust., Reg.) | VAR (Reg.) |
| --- | --- | --- | --- | --- | --- | --- | --- | --- | --- | --- | --- | --- | --- |
| Aguascalientes | 132.8 | 132.8 | 129.1 | 109.4 | 123.4 | 115.6 | 169.7 | 125.4 | 158.8 | 153.9 | 182.4 | 111.8 | **105.9** |
| Baja California | 359.1 | 359.1 | 350.1 | 173.3 | 104.7 | 212.5 | 144.4 | 341.1 | 183.3 | 2130.2 | 1957.2 | **103.2** | 206.3 |
| Baja California Sur | 92.4 | 92.5 | **86.8** | 134.9 | 115.2 | 107.4 | 135.3 | 89.8 | 183.7 | 2255.8 | 1891.9 | 128.3 | 130.1 |
| Campeche | 79.9 | 79.9 | 85.3 | 80.8 | **74.4** | 81.4 | 75.2 | 90.7 | 83.8 | 421.1 | 112.8 | 81.2 | 80.4 |
| Chiapas | 57.9 | 57.5 | 54.7 | 70.2 | 49.4 | 50.3 | 70.3 | 54.6 | 89.3 | 128.0 | 72.9 | **43.3** | 54.8 |
| Chihuahua | 129.8 | 130.2 | 127.5 | 181.6 | 115.8 | 119.0 | 185.4 | 124.9 | 294.2 | 146.6 | 159.1 | **109.5** | 120.1 |
| Coahuila | 86.6 | 87.0 | 88.5 | 143.6 | 88.4 | 89.2 | 143.6 | 91.1 | 127.7 | **76.5** | 82.8 | 94.8 | 95.2 |
| Colima | 71.2 | 71.2 | 69.3 | 89.1 | 63.8 | 63.5 | 89.1 | 70.0 | 97.9 | 194.5 | 90.7 | **61.3** | 64.9 |
| Durango | 138.6 | 138.8 | 135.3 | 162.2 | 129.7 | 133.9 | 188.2 | 131.8 | 218.5 | 317.5 | 320.9 | **124.4** | 146.6 |
| Guanajuato | 194.8 | 193.2 | 185.7 | 276.7 | 156.7 | 173.3 | 267.3 | 181.5 | 268.1 | 539.7 | 626.3 | 188.1 | **148.3** |
| Guerrero | 67.4 | 66.9 | 66.8 | 76.8 | 66.4 | **63.9** | 76.9 | 68.1 | 86.2 | 143.7 | 82.7 | 64.9 | 66.1 |
| Hidalgo | 93.7 | 93.8 | 94.2 | 128.2 | 80.9 | 81.0 | 128.2 | 94.8 | 151.2 | 96.4 | 102.4 | 84.3 | **70.7** |
| Jalisco | 70.5 | 85.7 | 68.2 | 102.3 | 61.7 | 59.7 | 102.2 | 60.0 | 105.9 | 87.3 | 73.8 | 71.3 | **58.5** |
| Mexico | 113.9 | 113.7 | 113.4 | 118.5 | 115.5 | 112.9 | 159.4 | 113.1 | 250.0 | 138.7 | 153.2 | **108.0** | 117.5 |
| Mexico City | - | - | - | - | - | - | - | - | - | - | - | - | - |
| Michoacan | 63.7 | 62.3 | **56.0** | 71.0 | 57.9 | 57.4 | 71.0 | 62.3 | 79.8 | 75.6 | 60.1 | 61.3 | 61.6 |
| Morelos | 69.8 | 70.0 | 69.9 | 95.8 | 62.6 | 64.1 | 95.8 | 69.9 | 105.6 | **61.8** | 68.4 | 64.8 | 62.6 |
| Nayarit | 79.0 | 79.0 | 73.6 | 90.0 | **62.8** | 66.6 | 90.0 | 72.4 | 120.1 | 95.0 | 83.5 | 65.2 | 64.1 |
| Nuevo Leon | 85.3 | 80.5 | 95.2 | 143.7 | 91.1 | 83.7 | 143.3 | 118.3 | 147.0 | 128.1 | 118.4 | **78.6** | 84.2 |
| Oaxaca | 71.5 | 80.1 | 69.4 | 93.0 | 58.3 | 59.8 | 92.9 | 62.4 | 108.4 | 96.2 | 99.0 | 68.9 | **57.2** |
| Puebla | 76.5 | 76.4 | 71.8 | 110.5 | 69.2 | 71.2 | 110.5 | 67.6 | 121.0 | 79.4 | 81.7 | 75.3 | **67.1** |
| Queretaro | 99.5 | 102.0 | 100.5 | 123.7 | **96.6** | 99.2 | 107.5 | 100.6 | 133.7 | 114.3 | 117.3 | 99.4 | 97.6 |
| Quintana Roo | 69.3 | 69.3 | 67.9 | 64.3 | 61.0 | 61.2 | 64.4 | 66.6 | 63.9 | 135.9 | 78.6 | 62.4 | **54.0** |
| San Luis Potosi | 89.8 | 89.8 | 86.2 | 106.7 | 78.5 | 79.2 | 106.7 | 82.7 | 140.7 | 92.4 | 94.8 | 76.2 | 75.7 |
| Sinaloa | 80.7 | 80.9 | 83.5 | 94.5 | 87.7 | 87.4 | 93.4 | 86.4 | **74.8** | 87.9 | 84.7 | 92.7 | 88.9 |
| Sonora | 90.3 | 91.2 | 90.9 | 137.4 | 94.8 | 95.8 | 137.3 | 94.8 | 127.9 | 505.9 | 279.0 | 106.3 | **90.2** |
| Tabasco | 78.7 | 85.2 | 79.4 | 74.3 | 77.1 | 78.0 | 74.2 | 76.8 | 113.3 | 234.2 | **72.7** | 79.4 | 76.8 |
| Tamaulipas | 90.0 | 89.9 | 89.5 | 112.4 | 78.1 | 79.1 | 112.4 | 89.4 | 103.6 | 144.6 | 104.0 | **76.2** | 82.5 |
| Tlaxcala | - | - | - | - | - | - | - | - | - | - | - | - | - |
| Veracruz | 57.7 | 54.6 | 54.7 | 80.5 | 49.1 | 52.9 | 80.5 | 55.8 | 78.1 | 106.4 | 82.7 | 69.0 | **45.7** |
| Yucatan | 74.2 | 73.2 | 71.4 | 86.5 | 61.3 | 67.8 | 86.9 | 83.2 | 74.7 | 286.4 | 134.5 | 79.5 | **60.1** |
| Zacatecas | 123.1 | 123.9 | 122.3 | 111.7 | 118.3 | 117.1 | 192.9 | 120.7 | 304.8 | 190.9 | 201.2 | **111.2** | 118.8 |

**Table 20:**
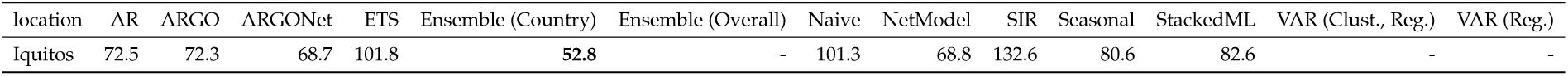
2-month ahead percent absolute errors in Peru for optimized models. Best performing models in each location are bolded. Lower values indicate stronger performance.

| location | AR | ARGO | ARGONet | ETS | Ensemble (Country) | Ensemble (Overall) | Naive | NetModel | SIR | Seasonal | StackedML | VAR (Clust., Reg.) | VAR (Reg.) |
| --- | --- | --- | --- | --- | --- | --- | --- | --- | --- | --- | --- | --- | --- |
| Iquitos | 72.5 | 72.3 | 68.7 | 101.8 | **52.8** | - | 101.3 | 68.8 | 132.6 | 80.6 | 82.6 | - | - |

**Table 21:** 2-month ahead percent absolute errors in Puerto Rico for optimized models. Best performing models in each location are bolded. Lower values indicate stronger performance.

| location | AR | ARGO | ARGONet | ETS | Ensemble (Country) | Ensemble (Overall) | Naive | NetModel | SIR | Seasonal | StackedML | VAR (Clust., Reg.) | VAR (Reg.) |
| --- | --- | --- | --- | --- | --- | --- | --- | --- | --- | --- | --- | --- | --- |
| San Juan | 79.3 | 67.1 | 54.0 | 55.1 | **31.7** | - | 55.1 | 47.5 | 42.0 | 60.6 | 39.7 | - | - |

**Table 22:**
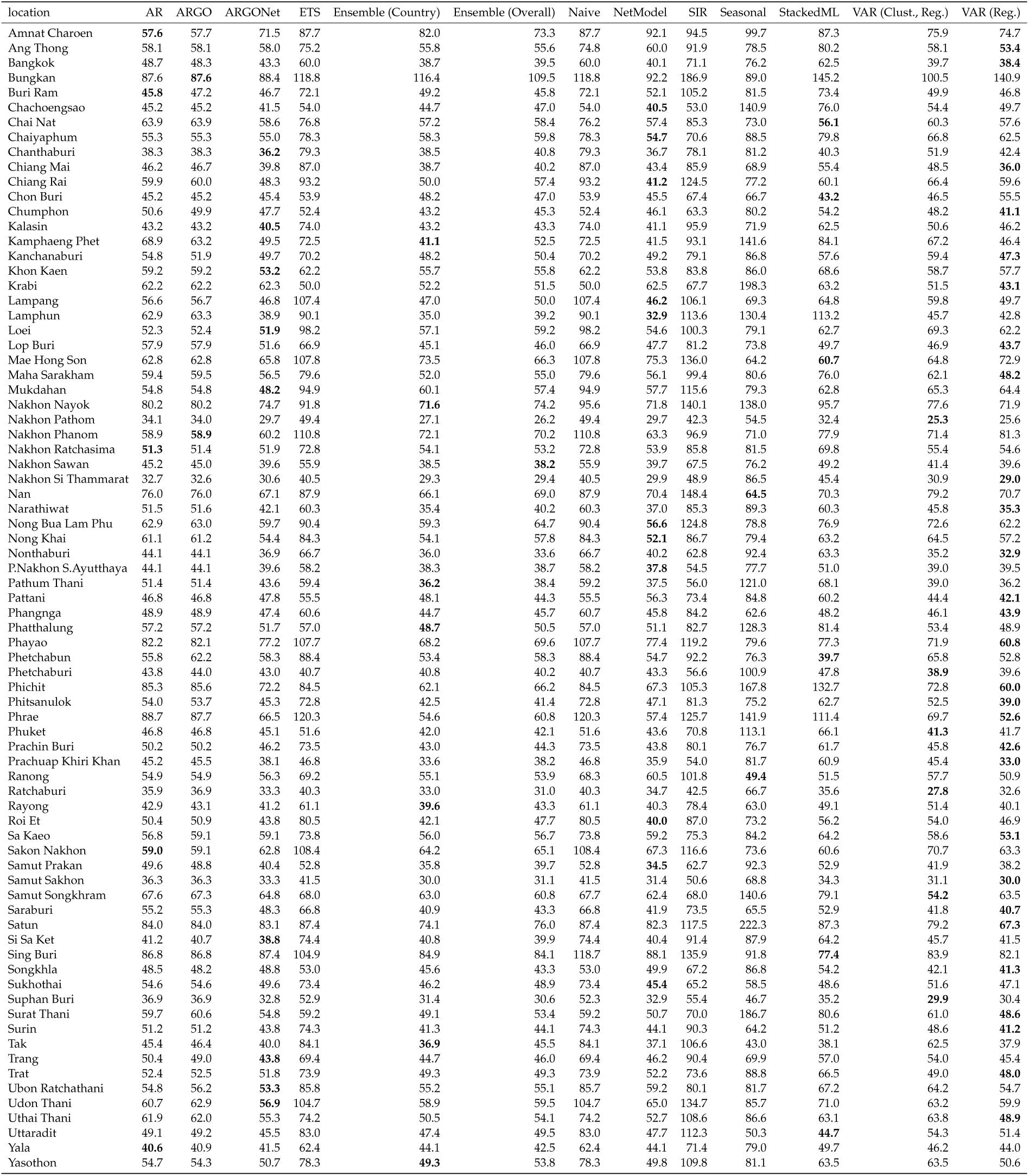
2-month ahead percent absolute errors in Thailand for optimized models. Best performing models in each location are bolded. Lower values indicate stronger performance.

#### 3.4.3 3-Month Ahead

**Table 23:** 3-month ahead percent absolute errors in Brazil for optimized models. Best performing models in each location are bolded. Lower values indicate stronger performance.

| location | AR | ARGO | ARGONet | ETS | Ensemble (Country) | Ensemble (Overall) | Naive | NetModel | SIR | Seasonal | StackedML | VAR (Clust., Reg.) | VAR (Reg.) |
| --- | --- | --- | --- | --- | --- | --- | --- | --- | --- | --- | --- | --- | --- |
| Acre | 63.6 | 65.7 | 63.9 | 104.1 | **60.0** | 62.9 | 104.1 | 64.3 | 101.2 | 97.0 | 74.6 | 68.0 | 69.3 |
| Alagoas | 68.7 | 69.0 | 60.1 | 101.3 | 60.9 | 63.3 | 101.3 | **57.4** | 79.8 | 119.1 | 80.9 | 72.0 | 67.0 |
| Amapa | 93.2 | 93.6 | 100.8 | 68.7 | 97.5 | 111.2 | **68.6** | 111.6 | 111.8 | 544.3 | 264.7 | 114.9 | 117.7 |
| Amazonas | 47.9 | 47.6 | 42.9 | 60.6 | 42.0 | **39.1** | 60.6 | 43.7 | 77.3 | 116.9 | 96.3 | 40.8 | 43.7 |
| Bahia | 47.1 | **44.2** | 53.6 | 97.8 | 54.6 | 57.3 | 97.8 | 66.2 | 106.8 | 51.4 | 44.2 | 67.6 | 61.7 |
| Ceara | **40.2** | 41.2 | 40.8 | 91.2 | 43.1 | 45.6 | 91.2 | 42.3 | 90.5 | 90.2 | 87.1 | 57.7 | 50.5 |
| Distrito Federal | 69.2 | 67.0 | **51.3** | 108.8 | 54.8 | 52.2 | 108.8 | 52.2 | 96.6 | 66.2 | 63.8 | 70.2 | 57.1 |
| Espirito Santo | 61.2 | 60.9 | 60.0 | 87.4 | 60.1 | 61.5 | 87.4 | **59.1** | 90.8 | 122.1 | 94.3 | 59.7 | 64.8 |
| Goias | 38.9 | 33.9 | **28.6** | 88.3 | 31.0 | 30.6 | 88.3 | 36.6 | 92.2 | 58.1 | 47.9 | 43.6 | 35.5 |
| Maranhao | 72.4 | 72.9 | 71.3 | 95.7 | **58.3** | 68.8 | 95.7 | 74.2 | 142.7 | 178.7 | 96.4 | 67.6 | 63.7 |
| Mato Grosso | 54.5 | 54.8 | 67.3 | 86.8 | 56.8 | 59.8 | 86.8 | 86.8 | 104.6 | 52.9 | **46.1** | 54.6 | 55.7 |
| Mato Grosso do Sul | 67.6 | 67.5 | 68.1 | 123.1 | 69.5 | 65.9 | 123.1 | 72.2 | 128.3 | **55.6** | 57.5 | 69.6 | 66.7 |
| Minas Gerais | 71.3 | 70.2 | 69.9 | 128.7 | **68.4** | 74.5 | 128.7 | 76.6 | 120.6 | 86.9 | 80.2 | 73.1 | 76.6 |
| Para | 41.5 | 42.8 | 40.3 | 77.7 | 45.1 | 43.6 | 77.7 | **40.0** | 121.6 | 125.2 | 67.4 | 73.7 | 45.0 |
| Paraiba | 47.4 | 47.1 | 53.7 | 82.9 | 47.9 | 52.9 | 82.9 | 67.9 | 75.5 | 74.6 | 59.6 | **44.4** | 52.3 |
| Parana | 74.0 | 72.9 | 65.6 | 120.0 | 70.0 | 66.3 | 120.0 | 70.1 | 94.5 | 67.0 | **60.8** | 74.0 | 68.2 |
| Pernambuco | 43.5 | 43.4 | **42.7** | 82.2 | 44.9 | 43.1 | 82.2 | 56.8 | 94.9 | 64.3 | 59.0 | 53.8 | 45.6 |
| Piaui | 51.8 | **51.6** | 57.7 | 101.5 | 55.5 | 55.6 | 101.5 | 68.6 | 107.0 | 104.8 | 96.7 | 56.6 | 56.3 |
| Rio Grande do Norte | 65.6 | 65.0 | 62.1 | 81.8 | 59.0 | 56.7 | 81.8 | 66.0 | 82.1 | 106.8 | 107.2 | 60.9 | **55.7** |
| Rio Grande do Sul | 80.0 | 76.2 | 74.1 | 148.0 | 78.6 | 76.5 | 148.0 | 73.5 | 127.8 | **71.0** | 81.8 | 80.7 | 82.5 |
| Rio de Janeiro | 76.6 | 74.2 | 82.3 | 102.2 | 68.3 | 74.3 | 102.2 | 102.8 | 109.4 | 415.6 | 216.8 | **52.7** | 63.3 |
| Rondonia | **45.1** | 46.9 | 50.6 | 71.9 | 47.5 | 49.9 | 71.9 | 58.2 | 81.0 | 140.3 | 69.6 | 54.5 | 47.6 |
| Roraima | 59.2 | 59.3 | 60.6 | **55.7** | 61.4 | 62.6 | 55.7 | 73.4 | 67.9 | 229.7 | 90.8 | 58.3 | 60.3 |
| Santa Catarina | 77.5 | 76.9 | **49.8** | 122.2 | 55.4 | 64.3 | 122.2 | 53.3 | 132.2 | 77.0 | 73.9 | 77.1 | 78.0 |
| Sao Paulo | 67.4 | 68.0 | 66.8 | 113.2 | 64.5 | 62.5 | 113.2 | 73.8 | 123.3 | **44.0** | 53.7 | 64.9 | 62.8 |
| Sergipe | 74.8 | 71.0 | 71.4 | 96.1 | 76.0 | 69.8 | 96.1 | 78.9 | 102.3 | 68.9 | **66.6** | 69.6 | 69.5 |
| Tocantins | **61.9** | 62.3 | 68.8 | 91.9 | 63.0 | 64.5 | 91.9 | 78.0 | 77.6 | 86.0 | 64.4 | 62.2 | 62.4 |

**Table 24:** 3-month ahead percent absolute errors in Colombia for optimized models. Best performing models in each location are bolded. Lower values indicate stronger performance.

| location | AR | ARGO | ARGONet | ETS | Ensemble (Country) | Ensemble (Overall) | Naive | NetModel | SIR | Seasonal | StackedML | VAR (Clust., Reg.) | VAR (Reg.) |
| --- | --- | --- | --- | --- | --- | --- | --- | --- | --- | --- | --- | --- | --- |
| Amazonas | 64.8 | 64.8 | 63.3 | - | 56.8 | 65.6 | 67.3 | 74.5 | 82.1 | 111.8 | 94.5 | **53.1** | 55.8 |
| Antioquia | 43.7 | 43.5 | 42.1 | - | **39.4** | 43.0 | 51.8 | 46.6 | 43.6 | 74.5 | 42.7 | 43.2 | 46.2 |
| Arauca | 43.0 | 43.0 | **41.2** | - | 50.8 | 43.9 | 49.4 | 41.7 | 82.7 | 127.3 | 102.1 | 47.4 | 47.4 |
| Atlantico | 65.9 | 65.9 | 59.9 | - | 69.6 | 62.7 | 96.6 | **58.7** | 112.5 | 85.5 | 67.0 | 70.7 | 69.5 |
| Bogota | 75.6 | 72.1 | 110.0 | - | 63.2 | 91.0 | 80.0 | 148.0 | 118.7 | 230.6 | 210.1 | **52.1** | 59.5 |
| Bolivar | 58.5 | 64.2 | 57.1 | - | **52.0** | 56.8 | 66.8 | 53.5 | 63.5 | 70.7 | 54.7 | 55.0 | 55.5 |
| Boyaca | 54.9 | 55.2 | **53.0** | - | 54.2 | 53.5 | 58.0 | 57.1 | 74.0 | 74.8 | 53.0 | 57.5 | 53.6 |
| Caldas | 37.3 | 37.3 | 35.2 | - | **33.3** | 34.1 | 39.9 | 37.1 | 57.9 | 60.7 | 42.3 | 35.2 | 35.7 |
| Caqueta | 60.4 | 60.4 | **50.9** | - | 63.2 | 53.9 | 74.6 | 52.3 | 105.8 | 72.8 | 57.6 | 62.1 | 58.3 |
| Casanare | 78.0 | 78.0 | 54.8 | - | 48.0 | 59.6 | 62.6 | **38.7** | 89.9 | 137.3 | 84.7 | 43.8 | 65.9 |
| Cauca | 49.8 | 49.8 | 53.9 | - | 50.1 | **48.9** | 49.6 | 59.0 | 68.1 | 64.6 | 52.5 | 51.2 | 50.3 |
| Cesar | 51.1 | 51.2 | 46.5 | - | 43.4 | 46.5 | 57.7 | 49.2 | 81.1 | 80.4 | 51.4 | **41.7** | 49.6 |
| Choco | 48.6 | 48.6 | 44.6 | - | 44.3 | **42.2** | 54.5 | 46.9 | 76.2 | 61.0 | 60.5 | 42.8 | 44.8 |
| Cordoba | 63.1 | 58.8 | 65.7 | - | 53.2 | 56.8 | 62.7 | 72.7 | 63.8 | 64.7 | 95.4 | 52.3 | **51.0** |
| Cundinamarca | 38.2 | 38.2 | **34.7** | - | 38.6 | 35.6 | 47.1 | 37.4 | 54.1 | 48.8 | 39.7 | 40.5 | 40.2 |
| Guainia | 69.7 | 69.7 | **66.1** | - | 72.6 | 72.1 | 105.4 | 66.5 | 109.8 | 94.5 | 102.6 | 76.2 | 72.4 |
| Guajira | 60.3 | 59.0 | 69.3 | - | 55.0 | 66.0 | 66.2 | 80.1 | 86.9 | 110.8 | 92.8 | 58.9 | **54.2** |
| Guaviare | 60.8 | 60.8 | 56.3 | - | 58.6 | 54.7 | 70.9 | 55.1 | 89.9 | 73.5 | 65.4 | 57.4 | **53.8** |
| Huila | 56.2 | 56.1 | 54.1 | - | 51.2 | **47.4** | 51.8 | 54.2 | 80.9 | 69.2 | 56.4 | - | 47.8 |
| Magdalena | 64.7 | 64.1 | 63.1 | - | 61.2 | 64.3 | 76.9 | 67.7 | 90.9 | 100.2 | 94.6 | **59.7** | 63.3 |
| Meta | 46.2 | 46.2 | 33.6 | - | 36.1 | 35.6 | 46.4 | **27.4** | 55.9 | 73.7 | 56.7 | 43.2 | 42.4 |
| Naria | 42.1 | 42.1 | **36.1** | - | 38.4 | 38.5 | 48.6 | 42.7 | 62.6 | 56.9 | 59.0 | 37.5 | 37.9 |
| Norte Santander | 37.3 | 36.5 | 39.0 | - | 35.4 | 40.1 | 37.8 | 45.3 | 54.9 | 63.5 | 41.9 | **32.8** | 37.0 |
| Putumayo | 49.9 | 49.5 | 39.2 | - | **38.2** | 38.6 | 57.1 | 46.5 | 66.8 | 57.2 | 57.8 | 49.8 | 40.5 |
| Quindio | 42.7 | 43.1 | 42.1 | - | 39.1 | 39.3 | 45.0 | 42.7 | 61.5 | 53.7 | 56.3 | 40.1 | **38.2** |
| Risaralda | **46.0** | 46.1 | 55.6 | - | 46.8 | 55.8 | 57.0 | 66.7 | 55.8 | 84.4 | 67.0 | 49.8 | 50.9 |
| San Andres | **74.5** | 78.3 | 81.9 | - | 159.4 | 81.9 | 130.1 | 87.6 | 159.4 | 92.2 | 93.3 | - | - |
| Santander | 30.5 | 30.5 | 28.7 | - | 26.1 | 28.2 | 29.9 | 31.2 | 33.2 | 71.2 | 39.9 | **25.1** | 28.6 |
| Sucre | 62.0 | 62.2 | 51.9 | - | **47.1** | 56.3 | 62.8 | 52.3 | 76.1 | 66.9 | 75.8 | 47.1 | 48.3 |
| Tolima | 36.9 | 36.9 | 37.0 | - | 32.9 | 36.5 | 39.6 | 39.0 | 64.2 | 40.2 | 38.3 | 33.1 | **32.7** |
| Valle | 42.6 | **41.7** | 43.8 | - | 42.8 | 42.3 | 50.6 | 54.4 | 69.4 | 61.4 | 41.9 | 45.2 | 45.1 |
| Vaupes | 174.4 | 174.4 | 207.0 | - | 151.3 | 155.9 | **125.0** | 243.7 | 134.4 | 356.8 | 350.3 | 139.7 | 139.7 |
| Vichada | 70.9 | 70.9 | 77.4 | - | 57.4 | 62.6 | 78.6 | 88.8 | 133.7 | 106.4 | 114.8 | 60.1 | **56.3** |

**Table 25:** 3-month ahead percent absolute errors in Malaysia for optimized models. Best performing models in each location are bolded. Lower values indicate stronger performance.

| location | AR | ARGO | ARGONet | ETS | Ensemble (Country) | Ensemble (Overall) | Naive | NetModel | SIR | Seasonal | StackedML | VAR (Clust., Reg.) | VAR (Reg.) |
| --- | --- | --- | --- | --- | --- | --- | --- | --- | --- | --- | --- | --- | --- |
| Johor | 42.1 | 42.1 | 41.1 | 43.9 | **34.0** | 40.0 | 45.0 | 40.5 | 67.5 | 51.9 | 36.2 | 37.9 | 38.7 |
| Kedah | 41.8 | 41.5 | 39.8 | 43.9 | 43.7 | 39.1 | 52.7 | 39.7 | 71.9 | 50.0 | 46.1 | 38.2 | **37.9** |
| Kelantan | 61.0 | 59.5 | **57.5** | 74.0 | 58.8 | 62.7 | 74.0 | 58.4 | 108.5 | 81.4 | 74.2 | 63.7 | 63.8 |
| Kuala Lumpur and Putrajaya | 39.9 | 42.8 | 38.7 | 42.2 | 35.7 | 38.7 | 42.6 | **35.3** | 66.2 | 57.9 | 36.8 | 38.0 | 37.5 |
| Labuan | 88.9 | 88.3 | 90.5 | **73.8** | 90.7 | 98.4 | 99.9 | 94.8 | 142.0 | 173.3 | 142.1 | 100.6 | 104.0 |
| Malacca | - | - | - | 43.7 | 37.2 | **36.6** | 41.6 | - | - | 55.9 | 40.7 | 37.0 | **36.6** |
| Negeri Sembilan | 33.9 | 34.0 | 32.7 | 34.9 | 30.9 | 32.6 | 35.3 | 33.1 | 51.1 | 44.7 | 32.6 | **30.8** | 31.4 |
| Pahang | 41.7 | 42.0 | 41.3 | 46.6 | 41.1 | 41.2 | 45.8 | 40.5 | 61.9 | 60.9 | 44.9 | 41.0 | **40.5** |
| Perak | 38.6 | 38.6 | 39.4 | 35.6 | 41.8 | 40.6 | **35.1** | 43.2 | 53.0 | 89.3 | 55.9 | 43.3 | 46.2 |
| Perlis | 69.4 | 69.4 | 69.1 | 67.4 | **61.3** | 63.7 | 70.4 | 68.9 | 116.4 | 112.1 | 91.5 | 62.4 | 62.5 |
| Pulau Pinang | - | - | - | 49.8 | **43.3** | 45.1 | 50.0 | - | - | 88.3 | 49.2 | 43.4 | 45.1 |
| Sabah | 38.2 | 38.4 | 40.0 | 42.1 | **36.8** | 43.0 | 48.3 | 43.7 | 85.6 | 49.6 | 37.8 | 40.9 | 42.2 |
| Sarawak | 45.5 | 45.6 | 45.0 | 42.3 | **38.1** | 46.0 | 39.0 | 47.9 | 57.6 | 81.1 | 40.0 | 51.0 | 49.0 |
| Selangor | 37.0 | 37.0 | 36.4 | 36.1 | 35.7 | 37.7 | 38.1 | 36.1 | 63.6 | 54.0 | **35.6** | 35.9 | 36.6 |
| Terengganu | 59.4 | 65.2 | 61.0 | 55.2 | **54.6** | 62.5 | 56.9 | 56.8 | 72.5 | 179.3 | 132.8 | 62.2 | 62.9 |

**Table 26:** 3-month ahead percent absolute errors in Mexico for optimized models. Best performing models in each location are bolded. Lower values indicate stronger performance.

| location | AR | ARGO | ARGONet | ETS | Ensemble (Country) | Ensemble (Overall) | Naive | NetModel | SIR | Seasonal | StackedML | VAR (Clust., Reg.) | VAR (Reg.) |
| --- | --- | --- | --- | --- | --- | --- | --- | --- | --- | --- | --- | --- | --- |
| Aguascalientes | 135.9 | 135.9 | 133.3 | 115.0 | 117.6 | **103.8** | 188.2 | 130.7 | 195.8 | 153.9 | 186.3 | 117.6 | 111.8 |
| Baja California | 521.6 | 521.6 | 530.5 | 223.3 | **117.9** | 202.9 | 255.6 | 635.6 | 281.7 | 2029.3 | 1974.3 | 275.2 | 196.6 |
| Baja California Sur | 67.4 | **67.3** | 77.1 | 161.5 | 124.1 | 102.9 | 161.7 | 92.5 | 174.0 | 1944.7 | 1699.5 | 130.9 | 143.5 |
| Campeche | 91.5 | 91.5 | 95.2 | 100.1 | **83.4** | 90.1 | 104.1 | 99.6 | 127.1 | 420.2 | 132.5 | 89.9 | **83.4** |
| Chiapas | 58.4 | 59.0 | **48.0** | 89.0 | 59.5 | 59.6 | 89.1 | 48.3 | 113.9 | 118.8 | 88.6 | 50.7 | 62.1 |
| Chihuahua | 146.1 | 146.6 | 137.9 | 209.4 | 121.6 | 131.5 | 215.0 | 129.2 | 344.5 | 155.4 | 168.7 | **118.8** | 127.3 |
| Coahuila | 94.4 | 94.1 | 90.1 | 146.6 | 102.9 | 92.3 | 146.7 | 87.7 | 157.7 | **77.0** | 81.3 | 101.2 | 103.0 |
| Colima | 75.8 | 75.8 | 70.2 | 107.9 | 72.2 | 77.4 | 107.9 | 72.3 | 129.8 | 189.6 | 77.1 | **66.1** | 76.9 |
| Durango | 221.4 | 220.8 | 220.0 | 280.8 | 199.3 | 216.6 | 319.5 | 219.3 | 427.6 | 483.8 | 617.6 | **197.4** | 221.3 |
| Guanajuato | 334.1 | 330.5 | 335.1 | 538.9 | 274.7 | 290.8 | 546.7 | 347.7 | 721.4 | 812.1 | 945.2 | 313.7 | **258.9** |
| Guerrero | 86.9 | 81.7 | 78.2 | 94.1 | 79.2 | 75.7 | 94.2 | 82.6 | 118.4 | 146.9 | 118.8 | 78.0 | **74.5** |
| Hidalgo | 94.6 | 94.7 | 92.5 | 147.6 | **83.0** | 89.8 | 147.6 | 92.3 | 196.6 | 101.3 | 110.9 | 97.1 | 83.0 |
| Jalisco | 77.1 | 74.5 | 65.7 | 140.2 | 72.0 | 66.6 | 140.3 | **59.7** | 133.6 | 85.6 | 87.7 | 81.9 | 71.5 |
| Mexico | 113.7 | 114.5 | 111.7 | 122.1 | 113.5 | **104.8** | 156.9 | 112.4 | 242.6 | 142.0 | 163.6 | 112.4 | 106.1 |
| Mexico City | - | - | - | - | - | - | - | - | - | - | - | - | - |
| Michoacan | 68.7 | 70.1 | 76.2 | 97.7 | 74.0 | 75.5 | 97.8 | 86.5 | 100.6 | 74.4 | **61.9** | 75.1 | 72.9 |
| Morelos | 77.9 | 77.7 | 74.2 | 117.5 | 73.1 | 71.6 | 117.5 | 73.1 | 159.5 | **61.7** | 72.6 | 73.6 | 73.3 |
| Nayarit | 81.4 | 81.4 | 80.0 | 111.1 | 72.9 | 76.1 | 112.1 | 81.7 | 170.3 | 93.8 | 91.6 | 71.6 | **70.7** |
| Nuevo Leon | 94.6 | 102.2 | 102.6 | 189.1 | 98.1 | **91.1** | 188.6 | 108.3 | 203.1 | 140.6 | 136.5 | 96.2 | 95.6 |
| Oaxaca | 81.8 | 82.9 | 75.0 | 116.2 | 71.1 | 67.3 | 116.2 | 69.8 | 143.7 | 91.2 | 94.8 | 74.7 | **65.9** |
| Puebla | 94.8 | 94.0 | 94.5 | 144.3 | 88.2 | 87.6 | 144.4 | 97.0 | 177.7 | **81.4** | 92.0 | 91.1 | 86.1 |
| Queretaro | 111.7 | 111.0 | 110.3 | 157.6 | 106.0 | 107.6 | 141.6 | 109.7 | 131.1 | 114.2 | 116.9 | **105.4** | 106.5 |
| Quintana Roo | 85.5 | 85.5 | 74.7 | 92.0 | 71.8 | **66.3** | 92.0 | 77.0 | 69.9 | 134.7 | 88.6 | 68.1 | 67.2 |
| San Luis Potosi | 96.7 | 96.5 | 87.8 | 115.1 | 83.3 | 83.5 | 115.1 | 84.7 | 110.3 | 93.6 | 103.3 | **81.6** | 84.5 |
| Sinaloa | 89.4 | 90.0 | 87.1 | 115.8 | 99.1 | 90.2 | 117.3 | **85.3** | 109.2 | 88.1 | 101.4 | 100.5 | 95.1 |
| Sonora | 113.9 | 113.9 | 112.3 | 167.5 | 117.7 | 113.8 | 167.5 | 111.7 | 162.2 | 535.1 | 160.1 | 133.1 | **108.1** |
| Tabasco | 82.8 | 81.0 | 78.8 | 82.2 | 79.0 | 79.1 | 82.2 | **78.8** | 116.9 | 237.4 | 89.0 | 86.7 | 84.9 |
| Tamaulipas | 99.4 | 99.4 | 102.5 | 139.7 | 88.3 | 94.1 | 139.7 | 105.7 | 145.1 | 143.9 | 106.5 | **87.9** | 94.5 |
| Tlaxcala | - | - | - | - | - | - | - | - | - | - | - | - | - |
| Veracruz | 72.1 | 59.4 | 55.0 | 108.9 | 59.8 | 54.4 | 108.9 | 55.3 | 73.9 | 107.7 | 89.6 | 75.3 | **53.2** |
| Yucatan | 93.8 | 93.8 | 96.8 | 124.4 | **77.0** | 85.6 | 124.4 | 103.8 | 115.8 | 288.2 | 173.1 | 91.8 | 78.4 |
| Zacatecas | 119.3 | 119.3 | 117.2 | 114.3 | 112.2 | 113.4 | 196.4 | 115.4 | 308.5 | 189.6 | 204.0 | **111.0** | 115.7 |

**Table 27:** 3-month ahead percent absolute errors in Peru for optimized models. Best performing models in each location are bolded. Lower values indicate stronger performance.

| location | AR | ARGO | ARGONet | ETS | Ensemble (Country) | Ensemble (Overall) | Naive | NetModel | SIR | Seasonal | StackedML | VAR (Clust., Reg.) | VAR (Reg.) |
| --- | --- | --- | --- | --- | --- | --- | --- | --- | --- | --- | --- | --- | --- |
| Iquitos | 86.3 | 85.0 | 78.1 | 120.9 | **69.7** | - | 120.3 | 82.8 | 165.3 | 81.4 | 86.2 | - | - |

**Table 28:** 3-month ahead percent absolute errors in Puerto Rico for optimized models. Best performing models in each location are bolded. Lower values indicate stronger performance.

| location | AR | ARGO | ARGONet | ETS | Ensemble (Country) | Ensemble (Overall) | Naive | NetModel | SIR | Seasonal | StackedML | VAR (Clust., Reg.) | VAR (Reg.) |
| --- | --- | --- | --- | --- | --- | --- | --- | --- | --- | --- | --- | --- | --- |
| San Juan | 106.6 | 89.7 | 79.2 | 76.8 | **43.9** | - | 76.8 | 74.1 | 67.6 | 59.9 | **43.9** | - | - |

**Table 29:**
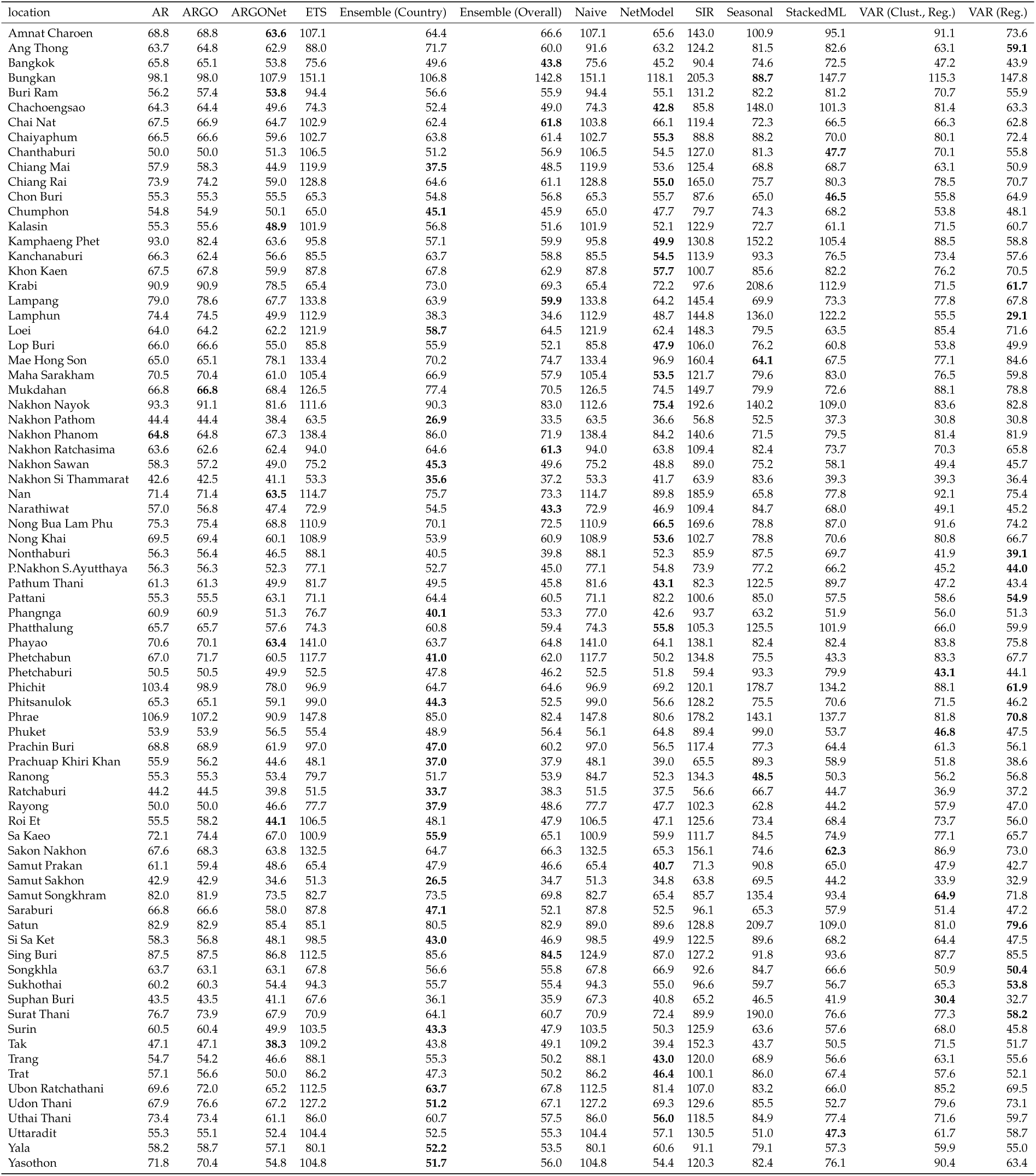
3-month ahead percent absolute errors in Thailand for optimized models. Best performing models in each location are bolded. Lower values indicate stronger performance.

### 3.5 PAE of Optimized Models versus Naive Persistence Baseline

**Table 30:** Number of locations per country where optimized models outperformed the naive persistence baseline at the 1-month ahead horizon, as measured through percent absolute error. Best performing models in each location are bolded. Higher values indicate stronger performance.

| country | no. of locations | AR | ARGO | ARGONet | ETS | Ensemble (Country) | Ensemble (Overall) | NetModel | SIR | Seasonal | StackedML | VAR (Clust., Reg.) | VAR (Reg.) |
| --- | --- | --- | --- | --- | --- | --- | --- | --- | --- | --- | --- | --- | --- |
| Brazil | 27 | **25** | 24 | 24 | 5 | **25** | **25** | 24 | 18 | 2 | 15 | 24 | 24 |
| Colombia | 33 | 26 | 26 | 26 | 0 | **31** | **31** | 25 | 7 | 0 | 7 | 22 | 18 |
| Malaysia | 15 | 5 | 5 | 5 | 3 | **12** | 7 | 5 | 0 | 0 | 0 | 7 | 8 |
| Mexico | 32 | 22 | 23 | 22 | 13 | **26** | 25 | 21 | 6 | 2 | 4 | 23 | 24 |
| Peru | 1 | **1** | **1** | **1** | 0 | **1** | 0 | **1** | 0 | 0 | **1** | 0 | 0 |
| Puerto Rico | 1 | **1** | **1** | **1** | 0 | **1** | 0 | **1** | **1** | 0 | **1** | 0 | 0 |
| Thailand | 77 | 70 | 70 | 74 | 14 | 74 | **75** | 74 | 15 | 3 | 36 | 71 | 69 |
| Overall | 186 | 150 | 150 | 153 | 35 | **170** | 163 | 151 | 47 | 7 | 64 | 147 | 143 |

**Table 31:**
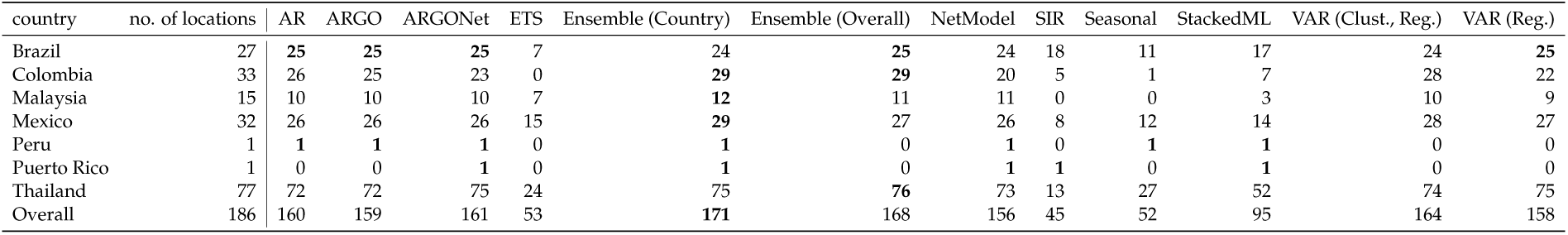
Number of locations per country where optimized models outperformed the naive persistence baseline at the 2-month ahead horizon, as measured through percent absolute error. Best performing models in each location are bolded. Higher values indicate stronger performance.

| country | no. of locations | AR | ARGO | ARGONet | ETS | Ensemble (Country) | Ensemble (Overall) | NetModel | SIR | Seasonal | StackedML | VAR (Clust., Reg.) | VAR (Reg.) |
| --- | --- | --- | --- | --- | --- | --- | --- | --- | --- | --- | --- | --- | --- |
| Brazil | 27 | **25** | **25** | **25** | 7 | 24 | **25** | 24 | 18 | 11 | 17 | 24 | **25** |
| Colombia | 33 | 26 | 25 | 23 | 0 | **29** | **29** | 20 | 5 | 1 | 7 | 28 | 22 |
| Malaysia | 15 | 10 | 10 | 10 | 7 | **12** | 11 | 11 | 0 | 0 | 3 | 10 | 9 |
| Mexico | 32 | 26 | 26 | 26 | 15 | **29** | 27 | 26 | 8 | 12 | 14 | 28 | 27 |
| Peru | 1 | **1** | **1** | **1** | 0 | **1** | 0 | **1** | 0 | **1** | **1** | 0 | 0 |
| Puerto Rico | 1 | 0 | 0 | **1** | 0 | **1** | 0 | **1** | **1** | 0 | **1** | 0 | 0 |
| Thailand | 77 | 72 | 72 | 75 | 24 | 75 | **76** | 73 | 13 | 27 | 52 | 74 | 75 |
| Overall | 186 | 160 | 159 | 161 | 53 | **171** | 168 | 156 | 45 | 52 | 95 | 164 | 158 |

**Table 32:** Number of locations per country where optimized models outperformed the naive persistence baseline at the 3-month ahead horizon, as measured through percent absolute error. Best performing models in each location are bolded. Higher values indicate stronger performance.

| country | no. of locations | AR | ARGO | ARGONet | ETS | Ensemble (Country) | Ensemble (Overall) | NetModel | SIR | Seasonal | StackedML | VAR (Clust., Reg.) | VAR (Reg.) |
| --- | --- | --- | --- | --- | --- | --- | --- | --- | --- | --- | --- | --- | --- |
| Brazil | 27 | **25** | **25** | **25** | 9 | **25** | **25** | 24 | 9 | 16 | 20 | **25** | **25** |
| Colombia | 33 | 27 | 28 | 26 | 0 | 29 | **30** | 21 | 3 | 4 | 12 | 29 | 29 |
| Malaysia | 15 | 10 | 9 | 10 | 11 | **14** | 12 | 11 | 0 | 1 | 9 | 11 | 11 |
| Mexico | 32 | 28 | 29 | 29 | 25 | **30** | **30** | 29 | 8 | 17 | 20 | 28 | 29 |
| Peru | 1 | **1** | **1** | **1** | 0 | **1** | 0 | **1** | 0 | **1** | **1** | 0 | 0 |
| Puerto Rico | 1 | 0 | 0 | 0 | 0 | **1** | 0 | **1** | **1** | **1** | **1** | 0 | 0 |
| Thailand | 77 | 73 | 73 | 75 | 41 | 76 | 75 | 72 | 8 | 52 | 64 | 73 | 77 |
| Overall | 186 | 164 | 165 | 166 | 86 | **176** | 172 | 159 | 29 | 92 | 127 | 166 | 171 |

### 3.6 Standard Models’ PAE by Country

#### 3.6.1 1-Month Ahead

**Table 33:** 1-month ahead percent absolute errors in Brazil for optimized models. Best performing models in each location are bolded. Lower values indicate stronger performance.

| location | AR | ARGO | ARGONet | Ensemble (Country) | Ensemble (Overall) | Naive | NetModel | SIR | Seasonal |
| --- | --- | --- | --- | --- | --- | --- | --- | --- | --- |
| Acre | 42.5 | 60.8 | 60.0 | **41.2** | 46.4 | 42.9 | 60.1 | 79.5 | 98.9 |
| Alagoas | 31.9 | 50.1 | 42.6 | **27.9** | 28.8 | 38.4 | 39.8 | 36.0 | 121.0 |
| Amapa | 109.2 | 328.8 | 326.0 | 45.3 | 46.9 | **39.9** | 326.9 | 84.8 | 539.3 |
| Amazonas | 44.2 | 90.0 | 34.2 | 25.3 | **24.3** | 27.1 | 29.2 | 30.5 | 119.0 |
| Bahia | **30.6** | 38.9 | 38.9 | 31.9 | 33.6 | 42.1 | 40.1 | 33.2 | 51.4 |
| Ceara | 40.3 | 76.4 | 51.0 | 31.2 | 33.2 | 35.6 | 52.3 | **30.1** | 90.5 |
| Distrito Federal | 36.2 | 46.5 | 41.5 | 37.5 | **35.6** | 45.8 | 36.4 | 43.7 | 66.3 |
| Espirito Santo | 39.6 | 47.2 | 46.4 | **31.6** | 35.2 | 35.4 | 56.5 | 39.7 | 125.7 |
| Goiás | 26.6 | 31.5 | 28.6 | **25.4** | 26.8 | 35.8 | 31.1 | 26.9 | 60.0 |
| Maranhao | 58.7 | 84.2 | 82.4 | **37.7** | 42.7 | 41.8 | 76.2 | 58.6 | 183.5 |
| Mato Grosso | **22.9** | 35.2 | 30.6 | 25.5 | 28.0 | 34.1 | 30.2 | 53.6 | 52.5 |
| Mato Grosso do Sul | **36.3** | 64.1 | 62.4 | 36.8 | 43.2 | 49.9 | 62.3 | 67.1 | 57.0 |
| Minas Gerais | 43.9 | 74.7 | 67.7 | **37.5** | 51.6 | 50.8 | 71.8 | 38.1 | 87.5 |
| Para | 43.5 | 52.1 | 50.8 | 34.2 | **33.9** | 35.5 | 49.7 | 40.4 | 132.7 |
| Paraiba | 28.4 | 44.9 | 39.6 | 30.1 | **27.6** | 37.9 | 42.6 | 30.3 | 75.6 |
| Parana | 35.7 | 42.3 | 42.5 | **34.8** | 42.4 | 50.7 | 57.3 | 36.2 | 67.2 |
| Pernambuco | **28.7** | 40.5 | 36.4 | 28.9 | 30.8 | 36.5 | 36.3 | 37.9 | 64.2 |
| Piaui | 53.8 | 74.8 | 67.7 | 38.7 | 37.4 | 44.6 | 65.9 | **34.7** | 109.2 |
| Rio de Janeiro | 129.3 | 168.0 | 117.9 | **36.1** | 79.9 | 45.5 | 116.2 | 44.0 | 420.0 |
| Rio Grande do Norte | 39.7 | 64.3 | 64.2 | **23.0** | 34.3 | 35.0 | 62.5 | 35.6 | 103.5 |
| Rio Grande do Sul | 69.9 | 76.4 | 78.7 | 81.3 | 65.4 | 77.1 | 67.0 | **61.6** | 71.1 |
| Rondonia | 36.7 | 47.0 | 42.5 | **31.1** | 33.2 | 31.2 | 41.6 | 35.2 | 144.8 |
| Roraima | 46.2 | 138.8 | 120.1 | **27.3** | 28.4 | 29.0 | 119.7 | 36.7 | 241.7 |
| Santa Catarina | 45.6 | 38.5 | **37.1** | 51.7 | 41.3 | 53.5 | 40.5 | 49.2 | 77.3 |
| Sao Paulo | 29.7 | 47.6 | 46.8 | **28.0** | 38.6 | 45.6 | 46.6 | 34.3 | 44.4 |
| Sergipe | **36.1** | 43.5 | 43.4 | 38.1 | 39.3 | 42.9 | 43.9 | 38.5 | 69.6 |
| Tocantins | 28.7 | 40.8 | 39.4 | **24.6** | 27.7 | 38.1 | 40.4 | 43.8 | 89.7 |

**Table 34:** 1-month ahead percent absolute errors in Colombia for optimized models. Best performing models in each location are bolded. Lower values indicate stronger performance.

| location | AR | ARGO | ARGONet | Ensemble (Country) | Ensemble (Overall) | Naive | NetModel | SIR | Seasonal |
| --- | --- | --- | --- | --- | --- | --- | --- | --- | --- |
| Amazonas | 44.5 | 62.4 | 63.5 | 46.6 | **38.5** | 42.8 | 71.1 | 60.4 | 148.3 |
| Antioquia | **16.8** | 30.9 | 29.1 | 17.2 | 19.0 | 20.5 | 28.0 | 20.0 | 80.8 |
| Arauca | 45.1 | 64.5 | 48.3 | 37.5 | 37.5 | **33.7** | 51.9 | 44.5 | 126.0 |
| Atlantico | 41.7 | 61.4 | 45.3 | 35.4 | 34.1 | 44.1 | 44.2 | **32.8** | 103.0 |
| Bogota | 252.4 | 428.2 | 328.3 | 107.7 | 104.8 | **103.9** | 337.7 | 158.3 | 612.7 |
| Bolivar | 31.3 | 40.0 | 40.0 | 29.9 | **29.6** | 30.9 | 40.6 | 30.1 | 83.2 |
| Boyaca | 38.1 | 38.0 | 36.8 | **33.5** | 36.4 | 34.4 | 38.3 | 33.7 | 80.9 |
| Caldas | 36.8 | 38.1 | **27.8** | 32.3 | 31.8 | 31.2 | 27.8 | 35.2 | 64.0 |
| Caqueta | 39.8 | 46.3 | 44.3 | 38.5 | 38.3 | **36.5** | 43.6 | 46.9 | 78.6 |
| Casanare | 43.3 | 47.3 | 45.0 | **28.5** | 29.2 | 29.3 | 47.0 | 36.7 | 116.7 |
| Cauca | 26.6 | 45.7 | 45.8 | 24.3 | **24.1** | 25.6 | 45.5 | 29.0 | 67.5 |
| Cesar | 26.1 | 40.7 | 38.9 | **24.5** | 25.5 | 27.4 | 38.7 | 29.6 | 86.4 |
| Choco | **29.2** | 49.9 | 46.4 | 30.4 | 30.0 | 30.0 | 37.1 | 34.3 | 63.7 |
| Cordoba | 36.6 | 54.5 | 52.9 | 32.6 | 33.0 | **31.0** | 52.8 | 34.8 | 81.1 |
| Cundinamarca | **18.0** | 24.4 | 23.0 | 18.8 | 20.1 | 21.1 | 21.4 | 26.1 | 56.5 |
| Guainia | 56.3 | 70.6 | 64.0 | **47.6** | 50.5 | 54.5 | 63.7 | 78.0 | 97.4 |
| Guajira | 45.6 | 77.1 | 75.8 | 41.6 | 43.6 | **38.4** | 75.4 | 41.0 | 145.2 |
| Guaviare | 44.1 | 64.8 | 61.0 | 44.2 | 44.6 | **41.0** | 62.4 | 52.4 | 76.2 |
| Huila | 26.8 | 32.1 | 28.8 | 24.6 | **21.6** | 23.6 | 28.2 | 28.6 | 67.7 |
| Magdalena | 36.8 | 48.7 | 38.0 | **35.8** | 36.1 | 37.5 | 37.9 | 37.2 | 103.6 |
| Meta | 18.3 | 27.4 | 23.5 | **18.0** | 18.8 | 20.7 | 25.5 | 21.5 | 78.8 |
| Naria | 34.1 | 35.6 | 33.7 | 33.6 | 34.5 | **32.0** | 32.9 | 43.7 | 59.0 |
| Norte Santander | 23.1 | 30.0 | 30.1 | 21.1 | 21.8 | **20.6** | 31.4 | 25.8 | 77.3 |
| Putumayo | 24.3 | 34.3 | 21.7 | 23.1 | 21.6 | 24.8 | **21.1** | 29.9 | 56.6 |
| Quindio | 20.6 | 42.1 | 30.7 | 18.3 | **18.3** | 21.4 | 37.4 | 22.0 | 55.4 |
| Risaralda | 31.3 | 52.3 | 37.1 | 26.1 | 25.8 | **25.7** | 41.2 | 27.9 | 90.9 |
| San Andres | **68.3** | 80.2 | 74.6 | 71.7 | 73.1 | 74.1 | 74.3 | 156.2 | 90.8 |
| Santander | 15.4 | 19.4 | 15.7 | **13.3** | 16.5 | 14.2 | 16.3 | 16.4 | 76.6 |
| Sucre | 47.2 | 68.3 | 64.7 | 33.6 | 33.9 | 33.2 | 64.9 | **33.1** | 98.3 |
| Tolima | 19.3 | 21.2 | 19.1 | 20.1 | **17.6** | 19.4 | 17.8 | 28.3 | 46.4 |
| Valle | 20.2 | 25.4 | 20.4 | **19.6** | 19.6 | 22.5 | 19.8 | 28.4 | 61.7 |
| Vaupes | 224.8 | 340.0 | 340.0 | 144.4 | 154.7 | 150.0 | 340.0 | **133.5** | 364.0 |
| Vichada | 76.2 | 119.3 | 119.3 | **51.9** | 55.0 | 54.2 | 119.3 | 62.7 | 117.0 |

**Table 35:** 1-month ahead percent absolute errors in Malaysia for optimized models. Best performing models in each location are bolded. Lower values indicate stronger performance.

| location | AR | ARGO | ARGONet | Ensemble (Country) | Ensemble (Overall) | Naive | NetModel | SIR | Seasonal |
| --- | --- | --- | --- | --- | --- | --- | --- | --- | --- |
| Johor | 25.0 | 31.4 | 31.4 | **19.6** | 21.3 | 21.8 | 31.4 | 28.4 | 52.6 |
| Kedah | **25.5** | 27.8 | 27.8 | 26.9 | 26.1 | 28.2 | 27.2 | 36.4 | 49.9 |
| Kelantan | 58.5 | 85.6 | 85.6 | 49.7 | 45.5 | **41.8** | 85.6 | 51.1 | 82.8 |
| Kuala Lumpur and Putrajaya | 24.4 | 25.8 | 25.1 | **20.8** | 24.0 | 23.4 | 24.5 | 28.3 | 58.9 |
| Labuan | 67.0 | 85.5 | 75.7 | 61.1 | **58.7** | 65.8 | 72.7 | 71.9 | 109.9 |
| Malacca | - | - | - | - | - | **21.0** | - | - | 54.8 |
| Negeri Sembilan | 22.7 | 30.8 | 31.0 | **19.1** | 20.3 | 20.4 | 33.1 | 24.9 | 47.6 |
| Pahang | 25.7 | 36.5 | 33.7 | **19.8** | 21.7 | 20.3 | 33.6 | 30.0 | 64.3 |
| Perak | 36.0 | 73.8 | 70.1 | 19.6 | 21.2 | **19.4** | 70.0 | 23.2 | 103.3 |
| Perlis | 51.5 | 84.5 | 79.4 | 47.8 | **46.0** | 52.5 | 79.1 | 66.6 | 116.8 |
| Pulau Pinang | - | - | - | - | - | 27.4 | - | **23.5** | 106.3 |
| Sabah | 33.9 | 35.2 | 33.5 | **27.7** | 32.2 | 31.6 | 34.0 | 53.0 | 48.7 |
| Sarawak | 26.1 | 51.5 | 51.8 | 20.9 | 22.6 | **20.6** | 56.5 | 27.6 | 85.0 |
| Selangor | 26.8 | 29.9 | 28.0 | 21.8 | 24.6 | **20.8** | 27.9 | 32.5 | 57.6 |
| Terengganu | 63.1 | 112.3 | 112.5 | 42.6 | 44.3 | **39.0** | 116.8 | 45.2 | 172.5 |

**Table 36:** 1-month ahead percent absolute errors in Mexico for optimized models. Best performing models in each location are bolded. Lower values indicate stronger performance.

| location | AR | ARGO | ARGONet | Ensemble (Country) | Ensemble (Overall) | Naive | NetModel | SIR | Seasonal |
| --- | --- | --- | --- | --- | --- | --- | --- | --- | --- |
| Aguascalientes | 145.7 | 181.7 | 149.2 | 120.5 | 105.6 | 100.8 | 142.4 | **74.9** | 153.9 |
| Baja California Sur | 703.2 | 2081.2 | 1594.4 | 88.4 | 85.9 | **78.8** | 1839.6 | 191.0 | 2126.3 |
| Baja California | 739.6 | 1836.8 | 1836.8 | 139.0 | **103.4** | 111.1 | 1880.2 | 221.2 | 1911.7 |
| Campeche | 106.3 | 200.5 | 162.2 | 88.5 | 75.4 | **74.2** | 160.8 | 107.0 | 418.7 |
| Chiapas | 79.7 | 97.2 | 95.3 | 54.6 | 50.0 | **49.7** | 95.2 | 74.9 | 139.8 |
| Chihuahua | 190.9 | 148.5 | 148.5 | **129.1** | 137.0 | 140.7 | 148.5 | 623.6 | 137.3 |
| Coahuila | 77.2 | 117.3 | 110.4 | 73.2 | **72.0** | 90.6 | 110.4 | 77.5 | 76.8 |
| Colima | 77.5 | 96.4 | 77.5 | **54.7** | 61.0 | 62.0 | 80.2 | 95.1 | 195.3 |
| Durango | 278.8 | 355.6 | 355.6 | 122.0 | 116.1 | **98.8** | 355.6 | 130.0 | 282.8 |
| Guanajuato | 122.1 | 321.8 | 239.5 | **77.1** | 89.2 | 86.4 | 326.0 | 148.5 | 308.3 |
| Guerrero | 59.8 | 103.1 | 96.6 | 57.0 | **54.0** | 56.0 | 95.9 | 120.9 | 151.6 |
| Hidalgo | 93.6 | 106.6 | 94.8 | **73.1** | 82.3 | 87.2 | 96.0 | 143.5 | 96.6 |
| Jalisco | 62.0 | 98.8 | 88.2 | **45.6** | 46.4 | 55.2 | 74.1 | 93.0 | 88.1 |
| Mexico City | - | - | - | - | - | - | - | - | - |
| Mexico | 144.4 | 154.9 | 154.9 | **128.6** | 134.2 | 136.3 | 154.9 | 291.9 | 145.6 |
| Michoacan | 38.6 | 55.2 | 60.0 | **36.6** | 40.6 | 39.2 | 72.9 | 67.4 | 77.9 |
| Morelos | 60.7 | 59.9 | 67.4 | **52.0** | 65.4 | 61.3 | 67.1 | 79.9 | 62.6 |
| Nayarit | 72.7 | 93.4 | 72.0 | **58.1** | 61.5 | 64.3 | 59.8 | 95.4 | 95.3 |
| Nuevo Leon | 101.7 | 128.0 | 125.2 | **71.7** | 89.5 | 87.4 | 134.0 | 212.3 | 122.8 |
| Oaxaca | 58.0 | 75.0 | 65.5 | 63.4 | **53.3** | 58.7 | 65.6 | 105.8 | 100.4 |
| Puebla | 57.8 | 66.3 | 69.6 | 55.0 | **55.0** | 58.1 | 75.3 | 66.3 | 78.2 |
| Queretaro | 108.1 | 100.8 | 98.3 | **75.0** | 79.0 | 76.0 | 97.9 | 130.6 | 110.4 |
| Quintana Roo | 52.0 | 62.7 | 65.3 | 43.9 | 42.4 | **41.6** | 91.0 | 46.1 | 142.1 |
| San Luis Potosi | 75.7 | 83.5 | **71.3** | 75.9 | 75.5 | 78.7 | 73.4 | 157.9 | 90.4 |
| Sinaloa | 66.2 | 89.4 | 89.7 | 55.6 | 59.4 | 63.7 | 102.9 | **45.3** | 89.0 |
| Sonora | 264.1 | 280.8 | 339.3 | 90.6 | 145.8 | **72.5** | 457.7 | 123.7 | 507.9 |
| Tabasco | 68.4 | 106.4 | 106.4 | 63.0 | 58.4 | **54.2** | 107.0 | 66.4 | 233.6 |
| Tamaulipas | 70.9 | 133.6 | 125.7 | 57.1 | **55.8** | 62.8 | 133.1 | 96.1 | 147.1 |
| Tlaxcala | - | - | - | - | - | - | - | - | - |
| Veracruz | 67.9 | 81.2 | 78.5 | **51.1** | 51.3 | 52.0 | 75.9 | 53.1 | 104.6 |
| Yucatan | 98.5 | 165.5 | 162.1 | **51.1** | 52.4 | 53.2 | 178.6 | 100.3 | 285.2 |
| Zacatecas | 200.3 | 205.9 | 205.9 | 132.0 | 143.7 | **107.8** | 205.9 | 269.2 | 192.2 |

**Table 37:**
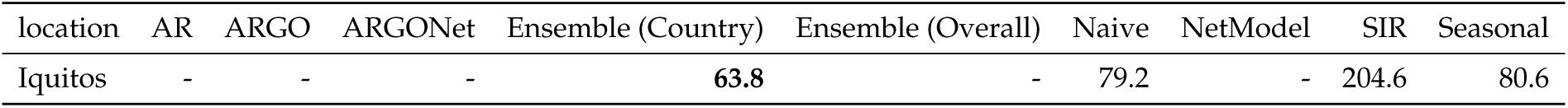
1-month ahead percent absolute errors in Peru for optimized models. Best performing models in each location are bolded. Lower values indicate stronger performance.

**Table 38:** 1-month ahead percent absolute errors in Puerto Rico for optimized models. Best performing models in each location are bolded. Lower values indicate stronger performance.

| location | AR | ARGO | ARGONet | Ensemble (Country) | Ensemble (Overall) | Naive | NetModel | SIR | Seasonal |
| --- | --- | --- | --- | --- | --- | --- | --- | --- | --- |
| San Juan | 23.8 | 34.5 | 34.2 | **20.1** | 23.0 | 30.7 | 36.8 | 22.9 | 60.7 |

**Table 39:**
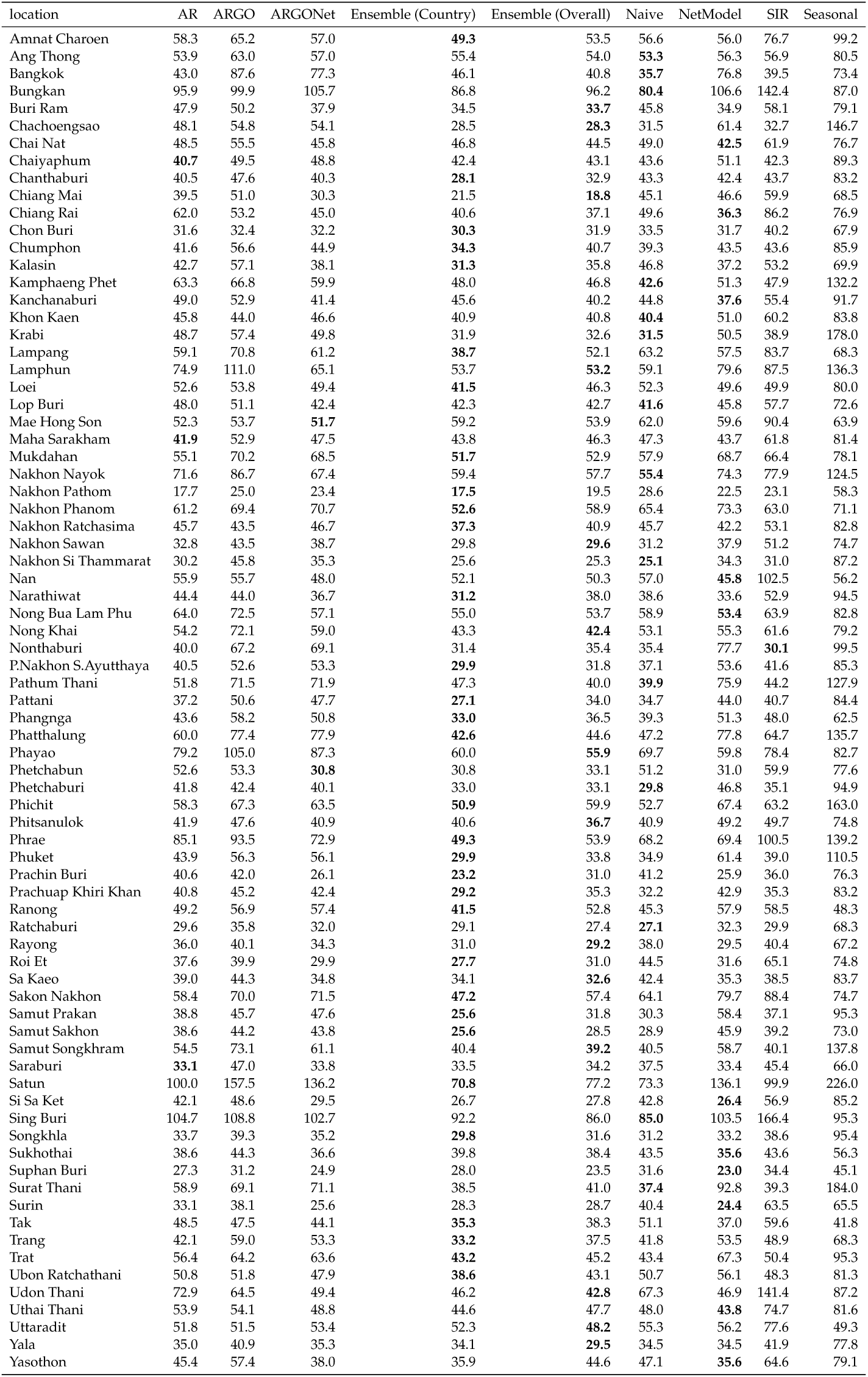
1-month ahead percent absolute errors in Thailand for optimized models. Best performing models in each location are bolded. Lower values indicate stronger performance.

#### 3.6.2 2-Month Ahead

**Table 40:** 2-month ahead percent absolute errors in Brazil for optimized models. Best performing models in each location are bolded. Lower values indicate stronger performance.

| location | AR | ARGO | ARGONet | Ensemble (Country) | Ensemble (Overall) | Naive | NetModel | SIR | Seasonal |
| --- | --- | --- | --- | --- | --- | --- | --- | --- | --- |
| Acre | 69.6 | 97.4 | 97.3 | **67.4** | 68.1 | 76.8 | 97.0 | 211.7 | 96.5 |
| Alagoas | 67.3 | 85.4 | 65.5 | 56.5 | 69.5 | 73.2 | **53.5** | 100.1 | 119.7 |
| Amapa | 179.0 | 475.6 | 394.2 | 68.7 | 108.2 | **58.5** | 390.4 | 187.1 | 540.8 |
| Amazonas | 75.5 | 104.6 | 82.0 | **42.5** | 61.7 | 44.4 | 76.9 | 57.7 | 116.4 |
| Bahia | **51.2** | 69.8 | 58.2 | 64.8 | 54.6 | 73.4 | 57.5 | 80.8 | 51.4 |
| Ceara | 78.8 | 93.7 | 73.4 | 63.3 | 57.1 | 65.6 | **56.1** | 82.0 | 90.7 |
| Distrito Federal | **64.5** | 85.6 | 73.2 | 81.8 | 64.7 | 79.6 | 66.0 | 97.5 | 66.2 |
| Espirito Santo | 78.3 | 92.9 | 91.5 | **59.7** | 80.8 | 64.8 | 103.8 | 68.8 | 123.6 |
| Goiias | 49.3 | 50.0 | 45.0 | 59.2 | **39.3** | 63.6 | 65.7 | 53.1 | 57.8 |
| Maranhao | 125.1 | 163.9 | 149.0 | 75.3 | 100.5 | **72.6** | 141.1 | 229.6 | 180.4 |
| Mato Grosso | **41.8** | 58.1 | 48.2 | 52.9 | 47.1 | 62.1 | 49.3 | 152.8 | 52.7 |
| Mato Grosso do Sul | 66.2 | 89.4 | 90.3 | 79.1 | 56.5 | 92.1 | 86.7 | 256.9 | **56.0** |
| Minas Gerais | 89.5 | 108.8 | 92.4 | 96.9 | **80.0** | 99.1 | 87.5 | 101.7 | 86.9 |
| Para | 75.7 | 91.1 | 66.9 | 68.6 | 70.3 | **59.0** | 61.9 | 92.0 | 127.6 |
| Paraiba | **44.5** | 72.2 | 64.7 | 44.9 | 48.0 | 61.2 | 58.3 | 67.7 | 75.0 |
| Parana | 62.9 | 80.9 | 85.2 | 63.2 | **56.9** | 96.3 | 88.6 | 121.7 | 67.0 |
| Pernambuco | 49.0 | 61.2 | 66.7 | 51.4 | **40.7** | 63.2 | 68.5 | 93.4 | 64.5 |
| Piaui | 108.0 | 132.2 | 56.9 | 69.1 | 63.0 | 73.7 | **48.1** | 78.2 | 104.9 |
| Rio de Janeiro | 300.1 | 420.8 | 284.1 | 88.2 | 213.8 | **81.6** | 212.9 | 104.0 | 410.4 |
| Rio Grande do Norte | 84.8 | 110.4 | 105.8 | **51.6** | 74.3 | 61.5 | 105.7 | 90.8 | 103.9 |
| Rio Grande do Sul | 89.8 | 77.4 | 81.0 | 116.7 | 103.2 | 117.5 | 83.7 | 202.2 | **71.0** |
| Rondonia | 59.4 | 82.3 | 80.6 | 43.9 | **40.6** | 53.3 | 81.2 | 61.2 | 139.7 |
| Roraima | 69.4 | 119.8 | 119.1 | **38.7** | 76.0 | 42.2 | 126.1 | 56.1 | 228.3 |
| Santa Catarina | 74.5 | **71.0** | 73.1 | 73.0 | 75.4 | 100.5 | 73.7 | 168.5 | 77.1 |
| Sao Paulo | 60.3 | 80.5 | 80.0 | 73.0 | 54.6 | 85.6 | 78.8 | 94.3 | **44.0** |
| Sergipe | 62.7 | 66.7 | 66.3 | 67.8 | **59.5** | 70.2 | 63.9 | 88.6 | 69.2 |
| Tocantins | 56.4 | 76.4 | 69.1 | **46.4** | 62.5 | 68.9 | 66.8 | 113.8 | 87.1 |

**Table 41:** 2-month ahead percent absolute errors in Colombia for optimized models. Best performing models in each location are bolded. Lower values indicate stronger performance.

| location | AR | ARGO | ARGONet | Ensemble (Country) | Ensemble (Overall) | Naive | NetModel | SIR | Seasonal |
| --- | --- | --- | --- | --- | --- | --- | --- | --- | --- |
| Amazonas | 67.3 | 105.8 | 88.1 | 56.3 | 76.4 | **55.0** | 78.2 | 68.1 | 119.2 |
| Antioquia | **31.5** | 53.0 | 40.6 | 31.8 | 37.7 | 37.5 | 40.2 | 32.8 | 76.3 |
| Arauca | 69.2 | 87.2 | 56.5 | 55.5 | 67.3 | **46.4** | 62.8 | 71.0 | 128.5 |
| Atlantico | 80.8 | 93.4 | 70.6 | 68.0 | **60.8** | 74.4 | 66.9 | 68.5 | 90.7 |
| Bogota | 214.3 | 367.9 | 257.3 | **87.0** | 142.7 | 96.1 | 235.2 | 163.8 | 431.6 |
| Bolivar | 54.4 | 61.3 | 59.0 | 51.4 | 51.0 | **50.9** | 59.8 | 60.7 | 79.2 |
| Boyaca | 50.3 | 53.6 | 50.2 | 48.7 | 49.4 | **45.9** | 53.9 | 54.7 | 78.0 |
| Caldas | 46.1 | 52.5 | **34.8** | 41.2 | 38.5 | 39.6 | 35.7 | 45.9 | 59.9 |
| Caqueta | 58.9 | 69.1 | 49.5 | 60.2 | 59.5 | 59.4 | **43.4** | 83.1 | 75.8 |
| Casanare | 85.2 | 92.7 | 77.3 | **44.2** | 47.3 | 46.6 | 70.3 | 64.4 | 134.3 |
| Cauca | 43.3 | 63.6 | 56.2 | 42.2 | **38.6** | 39.8 | 53.9 | 55.4 | 65.9 |
| Cesar | 50.8 | 68.7 | 59.1 | 47.8 | 47.1 | **44.7** | 58.5 | 57.8 | 84.6 |
| Choco | 47.4 | 62.2 | 52.1 | 45.6 | 45.0 | **42.3** | 52.9 | 52.1 | 61.5 |
| Cordoba | 65.8 | 71.5 | 64.0 | 54.0 | 47.1 | 49.2 | 65.2 | **46.2** | 77.7 |
| Cundinamarca | 31.7 | 33.8 | 40.3 | 32.2 | **30.9** | 35.7 | 33.5 | 42.6 | 50.0 |
| Guainia | 87.5 | 110.5 | 85.1 | 74.9 | **72.5** | 82.7 | 83.9 | 226.6 | 97.6 |
| Guajira | 69.8 | 98.9 | 81.7 | 63.8 | 65.2 | **58.4** | 74.9 | 67.9 | 129.4 |
| Guaviare | 59.7 | 75.1 | **52.3** | 60.4 | 57.1 | 59.3 | 56.0 | 70.8 | 73.3 |
| Huila | 46.0 | 52.4 | 47.8 | 39.7 | **39.6** | 40.0 | 43.8 | 55.1 | 66.4 |
| Magdalena | 67.9 | 79.7 | 72.3 | **58.4** | 66.6 | 62.0 | 70.8 | 66.1 | 103.3 |
| Meta | 36.6 | 53.4 | 43.0 | 35.3 | 35.5 | **34.6** | 44.3 | 38.7 | 75.7 |
| Naria | 42.6 | 42.9 | 35.4 | 41.2 | 40.7 | 40.9 | **32.8** | 58.4 | 58.9 |
| Norte Santander | 37.7 | 46.9 | 41.4 | 33.6 | **31.7** | 32.1 | 45.8 | 44.1 | 71.8 |
| Putumayo | 47.9 | 61.4 | 40.2 | 40.0 | 41.5 | 43.1 | **33.5** | 52.9 | 58.1 |
| Quindio | 37.8 | 63.8 | 40.2 | 31.1 | **27.4** | 35.2 | 36.2 | 38.9 | 53.2 |
| Risaralda | 50.5 | 86.7 | 53.9 | 43.5 | 44.4 | **42.8** | 45.9 | 46.7 | 86.0 |
| San Andres | 85.4 | 92.1 | 92.8 | **77.1** | 88.1 | 110.4 | 91.0 | 666.6 | 90.8 |
| Santander | 27.2 | 31.8 | **18.9** | 22.2 | 29.4 | 22.3 | 22.4 | 25.7 | 72.6 |
| Sucre | 72.8 | 80.6 | 58.6 | 53.2 | 51.2 | 51.4 | 67.1 | **47.9** | 82.5 |
| Tolima | 30.6 | 31.2 | 32.8 | 30.2 | 29.3 | 30.8 | **27.0** | 50.3 | 41.7 |
| Valle | 35.9 | 41.0 | 37.8 | 37.4 | 36.5 | 37.6 | **35.1** | 59.7 | 61.8 |
| Vaupes | 298.9 | 363.1 | 363.1 | 165.1 | 171.0 | **141.7** | 363.1 | 168.0 | 381.9 |
| Vichada | 104.8 | 111.8 | 96.4 | **65.5** | 75.8 | 66.8 | 116.2 | 99.7 | 109.9 |

**Table 42:** 2-month ahead percent absolute errors in Malaysia for optimized models. Best performing models in each location are bolded. Lower values indicate stronger performance.

| location | AR | ARGO | ARGONet | Ensemble (Country) | Ensemble (Overall) | Naive | NetModel | SIR | Seasonal |
| --- | --- | --- | --- | --- | --- | --- | --- | --- | --- |
| Johor | 40.5 | 42.6 | 42.5 | 40.4 | 43.1 | **38.0** | 42.4 | 51.7 | 53.1 |
| Kedah | **37.0** | 40.5 | 38.5 | 40.5 | 40.3 | 42.3 | 38.7 | 60.6 | 53.9 |
| Kelantan | 77.9 | 85.6 | 85.6 | 57.0 | **47.0** | 62.6 | 85.6 | 94.3 | 82.2 |
| Kuala Lumpur and Putrajaya | 39.6 | 38.2 | 35.6 | 38.5 | **33.3** | 35.8 | 33.9 | 51.0 | 59.1 |
| Labuan | 97.0 | 145.6 | 131.2 | **83.8** | 95.6 | **84.8** | 122.1 | 146.4 | 151.3 |
| Malacca | - | - | - | - | - | **34.6** | - | - | 56.8 |
| Negeri Sembilan | 31.8 | 38.4 | 37.9 | 31.4 | 30.4 | **30.0** | 36.3 | 38.2 | 48.4 |
| Pahang | 37.0 | 48.4 | 47.0 | 38.0 | 40.4 | **35.9** | 47.6 | 54.3 | 62.7 |
| Perak | 45.9 | 77.0 | 77.2 | 31.5 | 33.1 | **29.1** | 81.8 | 39.5 | 95.1 |
| Perlis | 78.3 | 115.5 | 115.6 | 69.4 | 71.6 | **65.7** | 116.2 | 108.1 | 122.4 |
| Pulau Pinang | - | - | - | - | - | **35.5** | - | 38.7 | 95.0 |
| Sabah | 40.1 | 40.3 | 37.0 | 41.7 | 37.9 | 42.6 | **35.4** | 83.4 | 48.8 |
| Sarawak | 38.1 | 55.7 | 56.3 | 30.2 | 34.6 | **27.3** | 59.2 | 44.5 | 81.7 |
| Selangor | 34.7 | 36.4 | 36.4 | **32.5** | 35.4 | 34.0 | 36.5 | 50.4 | 57.5 |
| Terengganu | 103.9 | 168.6 | 168.4 | 50.3 | 53.4 | **47.1** | 168.4 | 57.3 | 171.2 |

**Table 43:** 2-month ahead percent absolute errors in Mexico for optimized models. Best performing models in each location are bolded. Lower values indicate stronger performance.

| location | AR | ARGO | ARGONet | Ensemble (Country) | Ensemble (Overall) | Naive | NetModel | SIR | Seasonal |
| --- | --- | --- | --- | --- | --- | --- | --- | --- | --- |
| Aguascalientes | 182.6 | 184.8 | 184.8 | **100.9** | **100.9** | 169.7 | 184.8 | 142.4 | 153.9 |
| Baja California Sur | 1630.5 | 2262.8 | 2000.3 | 150.4 | 150.4 | **135.3** | 2092.4 | 928.8 | 2255.8 |
| Baja California | 1705.8 | 2148.1 | 2162.0 | 177.5 | 177.5 | **144.4** | 1878.8 | 225.1 | 2130.2 |
| Campeche | 150.2 | 263.0 | 193.5 | 109.6 | 109.6 | **75.2** | 183.0 | 192.5 | 421.1 |
| Chiapas | 107.9 | 127.2 | 111.6 | **47.6** | **47.6** | 70.3 | 104.4 | 135.7 | 128.0 |
| Chihuahua | 183.1 | 161.1 | 161.1 | **122.2** | **122.2** | 185.4 | 161.1 | 4778.4 | 146.6 |
| Coahuila | 117.7 | 120.5 | 117.0 | 86.5 | 86.5 | 143.6 | 113.7 | 208.3 | **76.5** |
| Colima | 112.5 | 148.3 | 128.8 | 90.1 | 90.1 | **89.1** | 118.7 | 224.7 | 194.5 |
| Durango | 404.6 | 407.4 | 407.4 | **146.0** | **146.0** | 188.2 | 407.4 | 230.1 | 317.5 |
| Guanajuato | 435.9 | 639.9 | 639.9 | **200.1** | **200.1** | 267.3 | 639.9 | 1000.9 | 539.7 |
| Guerrero | 85.2 | 127.9 | 84.3 | 82.4 | 82.4 | **76.9** | 87.8 | 259.2 | 143.7 |
| Hidalgo | 107.4 | 110.3 | 110.7 | 101.3 | 101.3 | 128.2 | 110.7 | 437.5 | **96.4** |
| Jalisco | **83.7** | 104.8 | 93.1 | 83.9 | 83.9 | 102.2 | 90.7 | 259.7 | 87.3 |
| Mexico City | - | - | - | - | - | - | - | - | - |
| Mexico | 149.6 | 149.6 | 149.6 | **122.4** | **122.4** | 159.4 | 149.6 | 732.3 | 138.7 |
| Michoacan | **67.7** | 85.2 | 80.6 | 68.4 | 68.4 | 71.0 | 78.6 | 198.0 | 75.6 |
| Morelos | 90.2 | 82.5 | - | 80.9 | 80.9 | 95.8 | 82.5 | 157.9 | **61.8** |
| Nayarit | 102.0 | 112.1 | 80.5 | **72.2** | **72.2** | 90.0 | 74.4 | 213.8 | 95.0 |
| Nuevo Leon | 154.4 | 165.5 | 148.7 | **102.2** | **102.2** | 143.3 | 145.8 | 1032.6 | 128.1 |
| Oaxaca | 84.0 | 133.4 | 106.9 | **64.2** | **64.2** | 92.9 | 87.4 | 310.1 | 96.2 |
| Puebla | 90.7 | 106.3 | - | 97.3 | 97.3 | 110.5 | 104.3 | 201.6 | **79.4** |
| Queretaro | 182.5 | 118.2 | 118.2 | **100.5** | **100.5** | 107.5 | 118.2 | 214.1 | 114.3 |
| Quintana Roo | 74.8 | 94.3 | 81.9 | **63.3** | **63.3** | 64.4 | 77.7 | 84.7 | 135.9 |
| San Luis Potosi | 100.6 | 99.6 | 102.3 | 111.4 | 111.4 | 106.7 | 98.5 | 671.9 | **92.4** |
| Sinaloa | 108.8 | 139.7 | 137.8 | **82.3** | **82.3** | 93.4 | 139.8 | 230.8 | 87.9 |
| Sonora | 488.8 | 552.5 | 552.5 | **123.5** | **123.5** | 137.3 | 552.5 | 540.3 | 505.9 |
| Tabasco | 97.4 | 135.5 | - | 98.9 | 98.9 | **74.2** | 139.3 | 173.0 | 234.2 |
| Tamaulipas | 125.6 | 164.2 | 152.5 | **90.1** | **90.1** | 112.4 | 151.2 | 357.9 | 144.6 |
| Tlaxcala | - | - | - | - | - | - | - | - | - |
| Veracruz | 88.1 | 107.9 | 71.9 | 69.0 | 69.0 | 80.5 | **62.6** | 113.0 | 106.4 |
| Yucatan | 179.9 | 262.2 | 209.0 | 88.1 | 88.1 | **86.9** | 201.6 | 427.9 | 286.4 |
| Zacatecas | 214.8 | 207.3 | 207.3 | **149.8** | **149.8** | 192.9 | 207.3 | 1048.1 | 190.9 |

**Table 44:** 2-month ahead percent absolute errors in Peru for optimized models. Best performing models in each location are bolded. Lower values indicate stronger performance.

| location | AR | ARGO | ARGONet | Ensemble (Country) | Ensemble (Overall) | Naive | NetModel | SIR | Seasonal |
| --- | --- | --- | --- | --- | --- | --- | --- | --- | --- |
| Iquitos | 114.2 | 110.4 | 110.4 | **52.0** | 67.9 | 101.3 | 110.4 | 772.8 | 80.6 |

**Table 45:** 2-month ahead percent absolute errors in Puerto Rico for optimized models. Best performing models in each location are bolded. Lower values indicate stronger performance.

| location | AR | ARGO | ARGONet | Ensemble (Country) | Ensemble (Overall) | Naive | NetModel | SIR | Seasonal |
| --- | --- | --- | --- | --- | --- | --- | --- | --- | --- |
| San Juan | 53.0 | 71.3 | 71.3 | **38.7** | 51.3 | 55.1 | 68.5 | 51.3 | 60.6 |

**Table 46:** 2-month ahead percent absolute errors in Thailand for optimized models. Best performing models in each location are bolded. Lower values indicate stronger performance.

| location | AR | ARGO | ARGONet | Ensemble (Country) | Ensemble (Overall) | Naive | NetModel | SIR | Seasonal |
| --- | --- | --- | --- | --- | --- | --- | --- | --- | --- |
| Amnat Charoen | 99.7 | 109.0 | 103.2 | **69.8** | 81.2 | 87.7 | 113.7 | 174.8 | 99.7 |
| Ang Thong | 75.9 | 77.3 | 73.8 | **61.7** | 68.8 | 74.8 | 72.0 | 117.1 | 78.5 |
| Bangkok | 66.8 | 91.5 | 82.8 | **52.5** | 67.7 | 60.0 | 74.4 | 74.9 | 76.2 |
| Bungkan | 114.2 | 118.2 | 117.0 | **73.4** | 85.0 | 118.8 | 115.7 | 515.8 | 89.0 |
| Buri Ram | 72.0 | 77.7 | 64.5 | 53.0 | **51.9** | 72.1 | 59.3 | 126.5 | 81.5 |
| Chachoengsao | 86.7 | 96.6 | 92.5 | **44.4** | 46.4 | 54.0 | 82.2 | 53.1 | 140.9 |
| Chai Nat | 72.5 | 75.6 | 60.9 | 88.3 | 85.8 | 76.2 | **58.1** | 125.9 | 73.0 |
| Chaiyaphum | 79.2 | **76.9** | 82.6 | 77.9 | 85.1 | 78.3 | 80.4 | 90.6 | 88.5 |
| Chanthaburi | 74.2 | 83.3 | 69.8 | **50.7** | 56.1 | 79.3 | 69.4 | 124.1 | 81.2 |
| Chiang Mai | 78.8 | 115.2 | 54.9 | 56.3 | 57.1 | 87.0 | **50.3** | 210.2 | 68.9 |
| Chiang Rai | 84.2 | 103.3 | 56.7 | 62.3 | 60.0 | 93.2 | **50.0** | 299.2 | 77.2 |
| Chon Buri | 50.1 | 50.3 | 49.9 | 47.4 | 48.5 | 53.9 | **46.0** | 71.3 | 66.7 |
| Chumphon | 63.2 | 65.2 | 62.7 | 45.4 | **42.8** | 52.4 | 60.1 | 63.3 | 80.2 |
| Kalasin | 65.3 | 85.1 | 63.2 | **51.7** | 60.7 | 74.0 | 59.4 | 122.8 | 71.9 |
| Kamphaeng Phet | 121.3 | 132.0 | 93.3 | **66.1** | 69.7 | 72.5 | 89.6 | 93.1 | 141.6 |
| Kanchanaburi | 79.3 | 77.9 | 65.6 | 60.6 | 65.7 | 70.2 | **57.9** | 117.3 | 86.8 |
| Khon Kaen | 64.2 | 82.3 | 72.5 | 71.1 | **64.4** | **62.2** | 72.5 | 105.5 | 86.0 |
| Krabi | 115.3 | 118.3 | 95.1 | 55.9 | 55.2 | **50.0** | 99.2 | 67.7 | 198.3 |
| Lampang | 98.5 | 124.4 | 93.6 | 51.3 | **50.5** | 107.4 | 84.7 | 180.7 | 69.3 |
| Lamphun | 120.9 | 152.9 | 143.2 | 68.0 | **60.6** | 90.1 | 118.6 | 266.9 | 130.4 |
| Loei | 83.4 | 85.3 | 81.2 | **58.4** | 60.6 | 98.2 | 77.3 | 144.1 | 79.1 |
| Lop Buri | 74.9 | 81.6 | 68.9 | **56.5** | 56.9 | 66.9 | 66.9 | 103.8 | 73.8 |
| Mae Hong Son | 82.0 | 88.4 | 87.1 | **64.1** | 73.9 | 107.8 | 80.9 | 307.3 | 64.2 |
| Maha Sarakham | 71.3 | 86.4 | 56.0 | 85.8 | 77.8 | 79.6 | **52.1** | 116.5 | 80.6 |
| Mukdahan | 72.7 | 106.1 | 101.0 | **62.3** | 67.2 | 94.9 | 94.3 | 172.4 | 79.3 |
| Nakhon Nayok | 128.0 | 141.4 | 152.4 | **86.0** | 89.9 | 95.6 | 148.2 | 140.1 | 138.0 |
| Nakhon Pathom | 31.4 | 43.3 | 32.0 | 38.8 | 35.6 | 49.4 | **30.0** | 42.3 | 54.5 |
| Nakhon Phanom | 95.8 | 105.3 | 103.8 | **57.0** | 60.4 | 110.8 | 98.0 | 261.5 | 71.0 |
| Nakhon Ratchasima | 71.3 | 72.8 | 62.9 | 62.4 | 62.7 | 72.8 | **60.5** | 121.4 | 81.5 |
| Nakhon Sawan | 59.5 | 68.5 | 60.9 | **44.7** | 46.9 | 55.9 | 59.1 | 142.4 | 76.2 |
| Nakhon Si Thammarat | 51.7 | 76.9 | 39.5 | 40.4 | 41.1 | 40.5 | **34.4** | 51.7 | 86.5 |
| Nan | 80.8 | 97.0 | 85.2 | 75.7 | 75.9 | 87.9 | 72.0 | 322.3 | **64.5** |
| Narathiwat | 71.1 | 76.3 | 52.7 | 56.9 | 62.4 | 60.3 | **39.5** | 109.1 | 89.3 |
| Nong Bua Lam Phu | 89.0 | 96.8 | 76.8 | **48.5** | 54.3 | 90.4 | 81.8 | 145.8 | 78.8 |
| Nong Khai | 79.2 | 106.8 | 89.4 | **55.8** | 60.6 | 84.3 | 81.1 | 134.8 | 79.4 |
| Nonthaburi | 72.5 | 94.5 | 84.3 | **58.8** | 66.0 | 66.7 | 76.6 | 62.9 | 92.4 |
| P.Nakhon S.Ayutthaya | 59.9 | 69.1 | 61.1 | **42.9** | 43.9 | 58.2 | 63.0 | 71.1 | 77.7 |
| Pathum Thani | 72.6 | 83.4 | 81.5 | **55.0** | 59.2 | 59.2 | 91.0 | 56.0 | 121.0 |
| Pattani | 62.9 | 77.4 | 87.5 | **49.5** | 50.2 | 55.5 | 90.0 | 82.3 | 84.8 |
| Phangnga | 68.7 | 80.7 | 67.9 | **43.4** | 46.4 | 60.7 | 62.0 | 88.0 | 62.6 |
| Phatthalung | 84.4 | 109.2 | 94.0 | **51.6** | 56.8 | 57.0 | 87.5 | 99.6 | 128.3 |
| Phayao | 115.4 | 133.6 | 98.9 | **61.7** | 70.6 | 107.7 | 81.1 | 230.7 | 79.6 |
| Phetchabun | 82.7 | 107.2 | 87.3 | **51.7** | 65.8 | 88.4 | 87.0 | 156.6 | 76.3 |
| Phetchaburi | 65.8 | 68.3 | 73.1 | 45.0 | 49.3 | **40.7** | 83.8 | 56.6 | 100.9 |
| Phichit | 115.2 | 130.7 | 128.7 | 95.9 | **94.7** | **84.5** | 109.5 | 105.3 | 167.8 |
| Phitsanulok | 68.3 | 82.9 | 70.5 | 47.9 | **46.1** | 72.8 | 77.2 | 98.3 | 75.2 |
| Phrae | 166.8 | 187.2 | 136.0 | 85.8 | **71.6** | 120.3 | 130.4 | 468.6 | 141.9 |
| Phuket | 71.8 | 86.7 | 66.3 | **45.5** | 59.1 | 51.6 | 73.5 | 70.8 | 113.1 |
| Prachin Buri | 71.0 | 73.7 | 47.3 | 59.6 | 50.0 | 73.5 | **42.1** | 96.8 | 76.7 |
| Prachuap Khiri Khan | 63.5 | 58.9 | 57.1 | 44.1 | **39.7** | 46.8 | 60.3 | 54.0 | 81.7 |
| Ranong | 63.8 | 65.6 | 67.5 | **47.0** | 58.1 | 68.3 | 66.3 | 101.9 | 49.4 |
| Ratchaburi | 43.8 | 49.7 | 52.1 | **35.3** | 36.8 | 40.3 | 56.6 | 42.5 | 66.7 |
| Rayong | 61.3 | 61.9 | 51.1 | **47.0** | 49.6 | 61.1 | 47.8 | 85.2 | 63.0 |
| Roi Et | 66.5 | 66.9 | 49.9 | 60.1 | 52.5 | 80.5 | **46.0** | 164.3 | 73.2 |
| Sa Kaeo | 65.3 | 73.5 | 57.5 | 72.8 | 66.6 | 73.8 | **54.1** | 89.3 | 84.2 |
| Sakon Nakhon | 91.6 | 106.8 | 105.0 | **58.7** | 65.6 | 108.4 | 103.0 | 213.3 | 73.6 |
| Samut Prakan | 64.8 | 67.6 | 73.8 | 51.3 | **46.0** | 52.8 | 73.9 | 72.2 | 92.3 |
| Samut Sakhon | 54.7 | 61.6 | 40.1 | 33.9 | **32.8** | 41.5 | 38.8 | 50.6 | 68.8 |
| Samut Songkhram | 92.4 | 118.5 | 87.2 | 76.1 | 71.8 | **67.7** | 72.5 | 72.5 | 140.6 |
| Saraburi | 56.3 | 71.5 | 52.0 | 51.9 | 55.5 | 66.8 | **41.5** | 89.3 | 65.5 |
| Satun | 129.1 | 229.1 | 143.5 | 84.7 | **72.1** | 87.4 | 135.9 | 145.5 | 222.3 |
| Si Sa Ket | 75.6 | 89.4 | 82.8 | 49.7 | **47.8** | 74.4 | 69.1 | 134.2 | 87.9 |
| Sing Buri | 124.1 | 93.8 | **89.0** | 117.9 | 113.5 | 118.7 | 91.8 | 521.7 | 91.8 |
| Songkhla | 59.6 | 73.9 | 58.8 | **40.9** | 45.7 | 53.0 | 59.7 | 73.9 | 86.8 |
| Sukhothai | 57.9 | 68.5 | 60.6 | **55.3** | 57.8 | 73.4 | 56.9 | 104.2 | 58.5 |
| Suphan Buri | 39.1 | 44.5 | 41.4 | 40.9 | 42.0 | 52.3 | **37.7** | 63.1 | 46.7 |
| Surat Thani | 110.1 | 136.4 | 117.9 | 72.8 | 62.4 | **59.2** | 115.9 | 70.0 | 186.7 |
| Surin | 60.8 | 78.0 | 57.6 | 55.9 | 55.9 | 74.3 | **45.4** | 151.6 | 64.2 |
| Tak | 65.5 | 75.1 | 69.1 | 46.1 | 56.9 | 84.1 | 57.0 | 162.6 | **43.0** |
| Trang | 73.3 | 92.5 | 79.9 | 45.0 | **43.2** | 69.4 | 76.5 | 116.0 | 69.9 |
| Trat | 82.1 | 84.2 | 107.3 | **51.3** | 56.6 | 73.9 | 104.2 | 100.0 | 88.8 |
| Ubon Ratchathani | 67.5 | 84.2 | 68.5 | 70.7 | **51.8** | 85.7 | 68.9 | 119.6 | 81.7 |
| Udon Thani | 93.4 | 93.7 | 82.0 | 79.3 | **67.4** | 104.7 | 76.3 | 638.0 | 85.7 |
| Uthai Thani | 79.1 | 75.8 | 84.0 | **67.7** | 74.0 | 74.2 | 74.3 | 144.2 | 86.6 |
| Uttaradit | 65.0 | 65.9 | 70.4 | **46.6** | 52.1 | 83.0 | 64.4 | 160.5 | 50.3 |
| Yala | 58.1 | 61.3 | 65.4 | **48.7** | 51.1 | 62.4 | 64.6 | 87.7 | 79.0 |
| Yasothon | 73.0 | 100.3 | 73.1 | **55.0** | 59.6 | 78.3 | 58.9 | 132.1 | 81.1 |

#### 3.6.3 3-Month Ahead

**Table 47:** 3-month ahead percent absolute errors in Brazil for optimized models. Best performing models in each location are bolded. Lower values indicate stronger performance.

| location | AR | ARGO | ARGONet | Ensemble (Country) | Ensemble (Overall) | Naive | NetModel | SIR | Seasonal |
| --- | --- | --- | --- | --- | --- | --- | --- | --- | --- |
| Acre | 94.3 | 98.1 | 98.1 | 89.5 | **86.0** | 104.1 | 98.1 | 547.2 | 97.0 |
| Alagoas | 93.7 | 108.0 | 69.2 | 92.1 | 85.7 | 101.3 | **63.2** | 230.5 | 119.1 |
| Amapa | 261.0 | 506.7 | 508.2 | 71.6 | 81.7 | **68.6** | 503.3 | 239.1 | 544.3 |
| Amazonas | 100.9 | 106.4 | 106.4 | 77.5 | 69.1 | **60.6** | 106.4 | 104.8 | 116.9 |
| Bahia | 60.6 | 72.0 | 68.8 | 59.3 | 65.9 | 97.8 | 69.1 | 151.0 | **51.4** |
| Ceara | 107.0 | 94.3 | 82.3 | 62.9 | **62.1** | 91.2 | 66.2 | 172.8 | 90.2 |
| Distrito Federal | 73.0 | 73.6 | 75.8 | 77.0 | 83.0 | 108.8 | 76.1 | 205.1 | **66.2** |
| Espirito Santo | 107.8 | 129.7 | 129.8 | 97.0 | 99.8 | **87.4** | 127.3 | 120.6 | 122.1 |
| Goiias | 66.0 | 68.6 | 65.1 | **43.2** | 45.9 | 88.3 | 82.1 | 92.1 | 58.1 |
| Maranhao | 184.8 | 190.8 | 178.4 | 110.9 | 110.0 | **95.7** | 192.3 | 1043.6 | 178.7 |
| Mato Grosso | 51.4 | 62.7 | 64.3 | 49.9 | **46.6** | 86.8 | 62.8 | 389.5 | 52.9 |
| Mato Grosso do Sul | 78.8 | 89.3 | 89.3 | 71.5 | 72.3 | 123.1 | 89.3 | 785.8 | **55.6** |
| Minas Gerais | 100.9 | 108.3 | 114.1 | 90.5 | 88.9 | 128.7 | 110.4 | 196.9 | **86.9** |
| Para | 101.5 | 109.1 | 94.3 | 81.5 | **67.6** | 77.7 | 90.6 | 160.9 | 125.2 |
| Paraiba | 59.2 | 72.4 | 74.8 | **57.2** | 62.6 | 82.9 | 73.5 | 135.0 | 74.6 |
| Parana | 72.5 | 82.6 | 83.8 | 79.2 | 80.5 | 120.0 | 81.4 | 303.8 | **67.0** |
| Pernambuco | 58.8 | 60.6 | 61.8 | **54.5** | 55.9 | 82.2 | 79.7 | 182.0 | 64.3 |
| Piaui | 147.2 | 132.2 | 91.2 | 83.0 | **72.2** | 101.5 | 84.1 | 160.9 | 104.8 |
| Rio de Janeiro | 438.4 | 431.0 | 301.3 | 190.4 | 191.5 | **102.2** | 244.7 | 219.4 | 415.6 |
| Rio Grande do Norte | 112.3 | 114.6 | 105.5 | **73.2** | 83.2 | 81.8 | 105.5 | 175.9 | 106.8 |
| Rio Grande do Sul | 83.9 | 84.5 | 84.5 | 100.9 | 103.7 | 148.0 | 86.9 | 623.9 | **71.0** |
| Rondonia | 80.3 | 85.0 | 98.8 | **59.6** | 75.0 | 71.9 | 106.8 | 105.2 | 140.3 |
| Roraima | 85.3 | 124.5 | 163.2 | 68.4 | 90.0 | **55.7** | 198.9 | 74.2 | 229.7 |
| Santa Catarina | 82.0 | 76.5 | 76.4 | 84.8 | 85.8 | 122.2 | **75.7** | 336.2 | 77.0 |
| Sao Paulo | 68.3 | 80.8 | 80.8 | 56.7 | 62.7 | 113.2 | 80.8 | 204.3 | **44.0** |
| Sergipe | 77.0 | 70.0 | 69.2 | 73.8 | 80.8 | 96.1 | 73.7 | 203.4 | **68.9** |
| Tocantins | 76.5 | 97.1 | 96.3 | **69.0** | 79.0 | 91.9 | 97.4 | 265.6 | 86.0 |

**Table 48:**
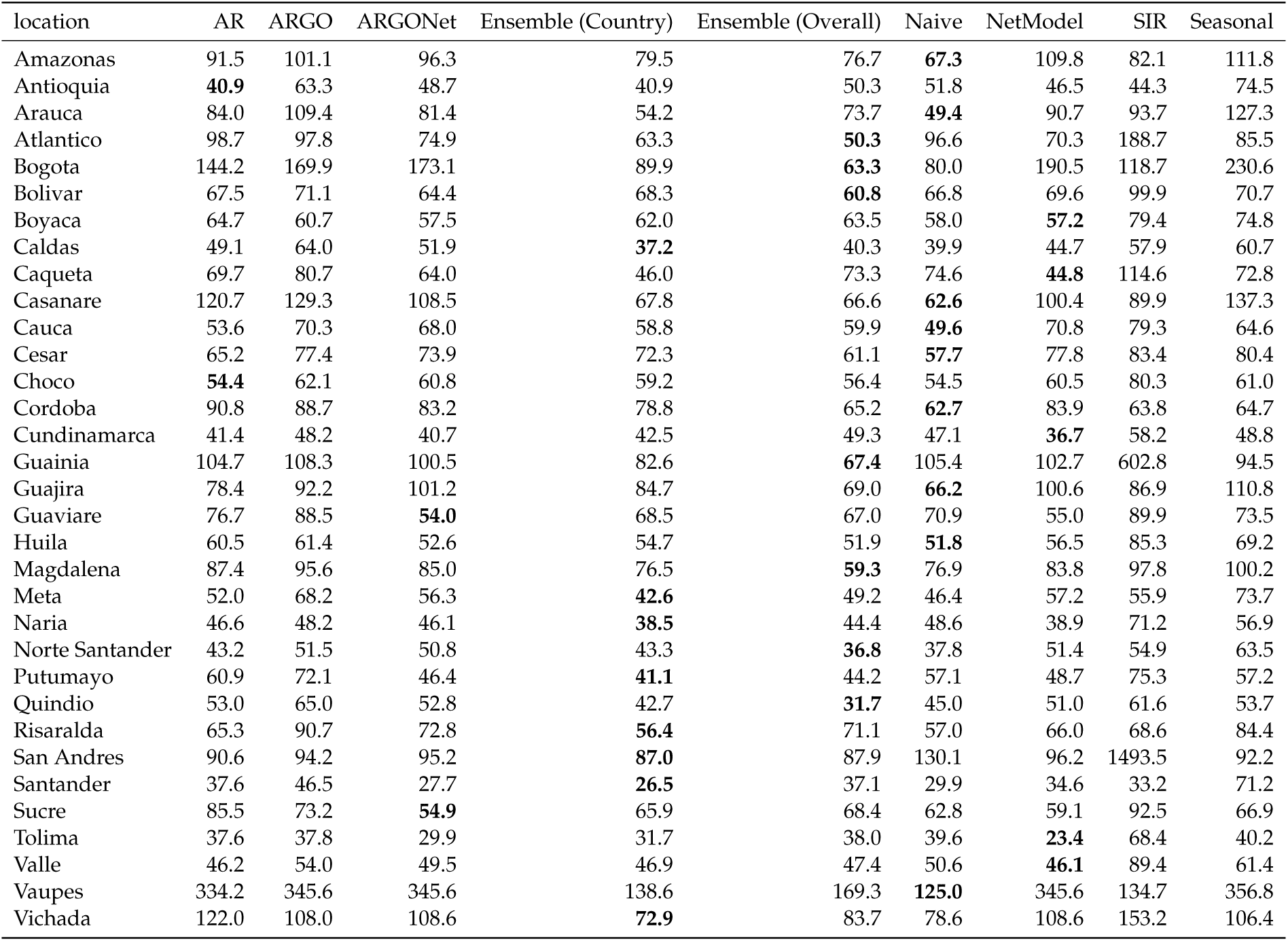
3-month ahead percent absolute errors in Colombia for optimized models. Best performing models in each location are bolded. Lower values indicate stronger performance.

**Table 49:**
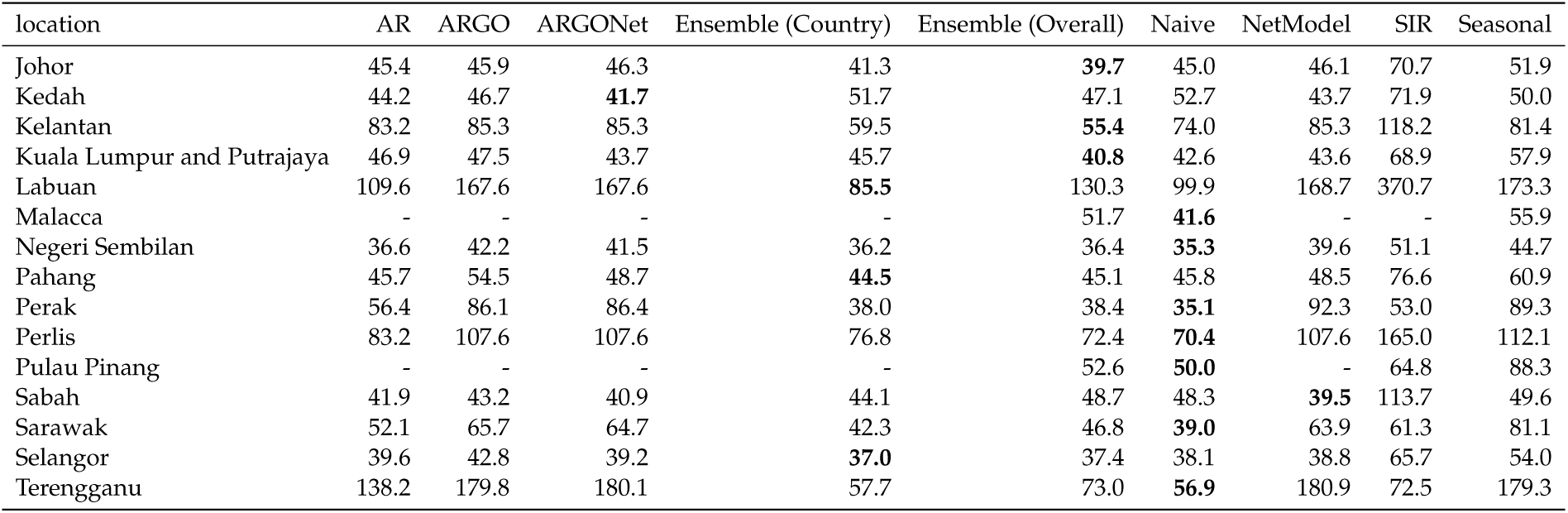
3-month ahead percent absolute errors in Malaysia for optimized models. Best performing models in each location are bolded. Lower values indicate stronger performance.

| location | AR | ARGO | ARGONet | Ensemble (Country) | Ensemble (Overall) | Naive | NetModel | SIR | Seasonal |
| --- | --- | --- | --- | --- | --- | --- | --- | --- | --- |
| Johor | 45.4 | 45.9 | 46.3 | 41.3 | **39.7** | 45.0 | 46.1 | 70.7 | 51.9 |
| Kedah | 44.2 | 46.7 | **41.7** | 51.7 | 47.1 | 52.7 | 43.7 | 71.9 | 50.0 |
| Kelantan | 83.2 | 85.3 | 85.3 | 59.5 | **55.4** | 74.0 | 85.3 | 118.2 | 81.4 |
| Kuala Lumpur and Putrajaya | 46.9 | 47.5 | 43.7 | 45.7 | **40.8** | 42.6 | 43.6 | 68.9 | 57.9 |
| Labuan | 109.6 | 167.6 | 167.6 | **85.5** | 130.3 | 99.9 | 168.7 | 370.7 | 173.3 |
| Malacca | - | - | - | - | 51.7 | **41.6** | - | - | 55.9 |
| Negeri Sembilan | 36.6 | 42.2 | 41.5 | 36.2 | 36.4 | **35.3** | 39.6 | 51.1 | 44.7 |
| Pahang | 45.7 | 54.5 | 48.7 | **44.5** | 45.1 | 45.8 | 48.5 | 76.6 | 60.9 |
| Perak | 56.4 | 86.1 | 86.4 | 38.0 | 38.4 | **35.1** | 92.3 | 53.0 | 89.3 |
| Perlis | 83.2 | 107.6 | 107.6 | 76.8 | 72.4 | **70.4** | 107.6 | 165.0 | 112.1 |
| Pulau Pinang | - | - | - | - | 52.6 | **50.0** | - | 64.8 | 88.3 |
| Sabah | 41.9 | 43.2 | 40.9 | 44.1 | 48.7 | 48.3 | **39.5** | 113.7 | 49.6 |
| Sarawak | 52.1 | 65.7 | 64.7 | 42.3 | 46.8 | **39.0** | 63.9 | 61.3 | 81.1 |
| Selangor | 39.6 | 42.8 | 39.2 | **37.0** | 37.4 | 38.1 | 38.8 | 65.7 | 54.0 |
| Terengganu | 138.2 | 179.8 | 180.1 | 57.7 | 73.0 | **56.9** | 180.9 | 72.5 | 179.3 |

**Table 50:** 3-month ahead percent absolute errors in Mexico for optimized models. Best performing models in each location are bolded. Lower values indicate stronger performance.

| location | AR | ARGO | ARGONet | Ensemble (Country) | Ensemble (Overall) | Naive | NetModel | SIR | Seasonal |
| --- | --- | --- | --- | --- | --- | --- | --- | --- | --- |
| Aguascalientes | 184.3 | 186.7 | 186.7 | 105.8 | **104.3** | 188.2 | 186.7 | 212.2 | 153.9 |
| Baja California Sur | 2079.4 | 2000.1 | 1935.2 | 199.9 | 204.6 | **161.7** | 1827.1 | 2940.3 | 1944.7 |
| Baja California | 2165.9 | 2099.1 | 2099.1 | 296.6 | 284.9 | **255.6** | 2099.1 | 274.3 | 2029.3 |
| Campeche | 206.4 | 343.6 | 311.8 | 142.7 | 165.8 | **104.1** | 312.6 | 321.2 | 420.2 |
| Chiapas | 121.7 | 123.5 | 118.3 | 72.4 | **68.9** | 89.1 | 113.7 | 241.6 | 118.8 |
| Chihuahua | 168.3 | 173.0 | 173.0 | 129.1 | **123.8** | 215.0 | 173.0 | 26487.9 | 155.4 |
| Coahuila | 116.5 | 121.8 | 124.5 | 104.4 | 93.4 | 146.7 | 124.5 | 268.3 | **77.0** |
| Colima | 140.5 | 190.3 | - | - | 111.4 | **107.9** | 128.1 | 829.7 | 189.6 |
| Durango | 688.6 | 633.0 | 633.0 | **241.1** | 251.2 | 319.5 | 633.0 | 427.0 | 483.8 |
| Guanajuato | 942.3 | 983.1 | 983.1 | **389.5** | 624.0 | 546.7 | 983.1 | 7111.9 | 812.1 |
| Guerrero | 101.5 | 142.0 | 98.7 | 100.4 | 101.0 | **94.2** | 107.4 | 344.2 | 146.9 |
| Hidalgo | 109.3 | 116.5 | 116.5 | 115.1 | 105.7 | 147.6 | 116.5 | 1773.2 | **101.3** |
| Jalisco | 95.1 | 102.7 | 99.4 | 91.2 | 100.8 | 140.3 | 98.4 | 752.8 | **85.6** |
| Mexico City | - | - | - | - | - | - | - | - | - |
| Mexico | 155.8 | 154.9 | 154.9 | **118.0** | 123.2 | 156.9 | 154.9 | 576.4 | 142.0 |
| Michoacan | 80.6 | 108.1 | 103.0 | 97.3 | 86.1 | 97.8 | 94.1 | 639.8 | **74.4** |
| Morelos | 95.4 | 94.3 | - | - | 69.1 | 117.5 | 98.9 | 348.4 | **61.7** |
| Nayarit | 108.9 | 111.0 | - | - | **78.1** | 112.1 | 96.9 | 628.4 | 93.8 |
| Nuevo Leon | 181.7 | 182.1 | 192.6 | **134.1** | 140.3 | 188.6 | 200.7 | 5245.7 | 140.6 |
| Oaxaca | 102.4 | 115.1 | 96.3 | 73.6 | **72.1** | 116.2 | 88.0 | 1547.1 | 91.2 |
| Puebla | 101.5 | 109.1 | 109.1 | 114.1 | 82.5 | 144.4 | 109.1 | 690.3 | **81.4** |
| Queretaro | 214.6 | 121.2 | 122.7 | 159.6 | 145.8 | 141.6 | 122.7 | 386.5 | **114.2** |
| Quintana Roo | 106.9 | 132.2 | - | - | 108.7 | **92.0** | 103.8 | 138.8 | 134.7 |
| San Luis Potosi | 95.0 | 99.0 | - | - | 97.7 | 115.1 | 106.3 | 3112.7 | **93.6** |
| Sinaloa | 135.8 | 141.9 | 139.9 | 102.7 | 101.3 | 117.3 | 139.9 | 1113.9 | **88.1** |
| Sonora | 603.4 | 597.7 | 597.7 | **154.5** | 155.9 | 167.5 | 597.7 | 1867.8 | 535.1 |
| Tabasco | 109.8 | 161.1 | 159.6 | 94.0 | 120.7 | **82.2** | 159.6 | 490.1 | 237.4 |
| Tamaulipas | 145.1 | 160.9 | 156.3 | **94.3** | 101.2 | 139.7 | 155.6 | 1127.7 | 143.9 |
| Tlaxcala | - | - | - | - | - | - | - | - | - |
| Veracruz | 92.3 | 117.0 | 98.2 | 96.6 | **91.3** | 108.9 | 91.9 | 244.4 | 107.7 |
| Yucatan | 251.6 | 306.0 | - | - | 128.2 | **124.4** | 154.9 | 740.3 | 288.2 |
| Zacatecas | 208.8 | 207.6 | 207.6 | **149.8** | 186.4 | 196.4 | 207.6 | 2928.5 | 189.6 |

**Table 51:**
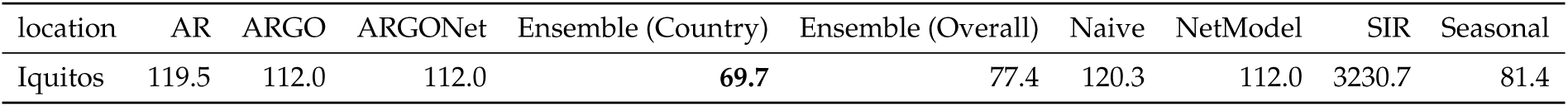
3-month ahead percent absolute errors in Peru for optimized models. Best performing models in each location are bolded. Lower values indicate stronger performance.

| location | AR | ARGO | ARGONet | Ensemble (Country) | Ensemble (Overall) | Naive | NetModel | SIR | Seasonal |
| --- | --- | --- | --- | --- | --- | --- | --- | --- | --- |
| Iquitos | 119.5 | 112.0 | 112.0 | **69.7** | 77.4 | 120.3 | 112.0 | 3230.7 | 81.4 |

**Table 52:**
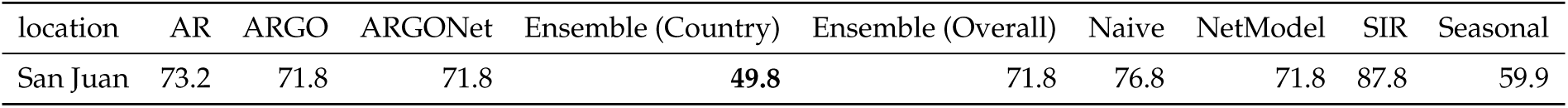
3-month ahead percent absolute errors in Puerto Rico for optimized models. Best performing models in each location are bolded. Lower values indicate stronger performance.

| location | AR | ARGO | ARGONet | Ensemble (Country) | Ensemble (Overall) | Naive | NetModel | SIR | Seasonal |
| --- | --- | --- | --- | --- | --- | --- | --- | --- | --- |
| San Juan | 73.2 | 71.8 | 71.8 | **49.8** | 71.8 | 76.8 | 71.8 | 87.8 | 59.9 |

**Table 53:** 3-month ahead percent absolute errors in Thailand for optimized models. Best performing models in each location are bolded. Lower values indicate stronger performance.

| location | AR | ARGO | ARGONet | Ensemble (Country) | Ensemble (Overall) | Naive | NetModel | SIR | Seasonal |
| --- | --- | --- | --- | --- | --- | --- | --- | --- | --- |
| Amnat Charoen | 117.9 | 121.3 | 152.6 | **80.2** | **80.2** | 107.1 | 145.0 | 434.9 | 100.9 |
| Ang Thong | 91.1 | 83.8 | 86.7 | **65.7** | **65.7** | 91.6 | 85.2 | 216.5 | 81.5 |
| Bangkok | 78.7 | 90.2 | 77.3 | 65.6 | 65.6 | 75.6 | **63.5** | 105.4 | 74.6 |
| Bungkan | 117.5 | 118.4 | 183.2 | **79.2** | **79.2** | 151.1 | 182.4 | 1532.5 | 88.7 |
| Buri Ram | 86.4 | 94.8 | 78.9 | **65.3** | **65.3** | 94.4 | 67.8 | 220.4 | 82.2 |
| Chachoengsao | 128.2 | 133.0 | 135.6 | **56.4** | **56.4** | 74.3 | 122.5 | 85.8 | 148.0 |
| Chai Nat | 81.6 | 73.5 | **67.1** | 77.3 | 77.3 | 103.8 | 67.8 | 254.5 | 72.3 |
| Chaiyaphum | 91.4 | 87.2 | 92.3 | **74.7** | **74.7** | 102.7 | 92.6 | 154.7 | 88.2 |
| Chanthaburi | 99.2 | 102.6 | 109.8 | **56.8** | **56.8** | 106.5 | 102.9 | 266.6 | 81.3 |
| Chiang Mai | 101.1 | 115.7 | 87.5 | 71.5 | 71.5 | 119.9 | 85.6 | 585.7 | **68.8** |
| Chiang Rai | 103.4 | 103.2 | 84.9 | 71.9 | 71.9 | 128.8 | **67.8** | 998.1 | 75.7 |
| Chon Buri | 59.8 | 62.8 | 61.5 | **50.3** | **50.3** | 65.3 | 56.7 | 108.9 | 65.0 |
| Chumphon | 69.0 | 77.8 | 73.0 | **54.3** | **54.3** | 65.0 | 67.7 | 79.6 | 74.3 |
| Kalasin | 77.4 | 89.4 | 85.8 | **64.3** | **64.3** | 101.9 | 79.0 | 241.8 | 72.7 |
| Kamphaeng Phet | 161.1 | 164.6 | 151.1 | 96.2 | 96.2 | **95.8** | 126.3 | 153.7 | 152.2 |
| Kanchanaburi | 99.1 | 100.7 | 87.3 | **75.2** | **75.2** | 85.5 | 85.4 | 242.2 | 93.3 |
| Khon Kaen | 79.5 | 87.7 | 75.4 | 84.5 | 84.5 | 87.8 | **70.9** | 176.9 | 85.6 |
| Krabi | 174.6 | 225.1 | 146.5 | 75.8 | 75.8 | **65.4** | 140.8 | 97.6 | 208.6 |
| Lampang | 115.5 | 125.4 | 120.3 | **59.8** | **59.8** | 133.8 | 110.9 | 384.8 | 69.9 |
| Lamphun | 152.5 | 161.7 | 148.3 | **90.1** | **90.1** | 112.9 | 129.0 | 711.3 | 136.0 |
| Loei | 95.8 | 104.7 | 107.5 | **63.8** | **63.8** | 121.9 | 105.9 | 289.3 | 79.5 |
| Lop Buri | 91.2 | 90.4 | 76.2 | **60.8** | **60.8** | 85.8 | 74.7 | 177.4 | 76.2 |
| Mae Hong Son | 80.3 | 90.6 | 91.3 | 68.7 | 68.7 | 133.4 | 86.1 | 841.6 | **64.1** |
| Maha Sarakham | 83.5 | 97.0 | 80.3 | 75.4 | 75.4 | 105.4 | **66.4** | 197.3 | 79.6 |
| Mukdahan | 93.1 | 109.4 | 107.9 | **66.7** | **66.7** | 126.5 | 119.6 | 353.0 | 79.9 |
| Nakhon Nayok | 144.1 | 143.9 | 148.9 | **101.5** | **101.5** | 112.6 | 150.0 | 192.6 | 140.2 |
| Nakhon Pathom | 45.1 | 52.8 | 43.5 | 47.9 | 47.9 | 63.5 | **38.4** | 62.8 | 52.5 |
| Nakhon Phanom | 104.4 | 106.4 | 135.6 | **61.7** | **61.7** | 138.4 | 134.0 | 894.3 | 71.5 |
| Nakhon Ratchasima | 87.9 | 82.6 | 76.1 | 74.3 | 74.3 | 94.0 | **73.6** | 234.0 | 82.4 |
| Nakhon Sawan | 76.6 | 80.6 | 83.4 | **64.4** | **64.4** | 75.2 | 75.9 | 433.3 | 75.2 |
| Nakhon Si Thammarat | 65.7 | 85.2 | 71.4 | **46.6** | **46.6** | 53.3 | 69.1 | 76.6 | 83.6 |
| Nan | 85.4 | 99.1 | 95.6 | 90.7 | 90.7 | 114.7 | 85.2 | 1047.5 | **65.8** |
| Narathiwat | 77.1 | 86.5 | 71.8 | **63.3** | **63.3** | 72.9 | 66.1 | 217.9 | 84.7 |
| Nong Bua Lam Phu | 99.6 | 104.3 | 81.8 | **56.2** | **56.2** | 110.9 | 97.7 | 275.3 | 78.8 |
| Nong Khai | 97.2 | 106.1 | 95.7 | **70.7** | **70.7** | 108.9 | 84.0 | 268.4 | 78.8 |
| Nonthaburi | 90.5 | 101.7 | 81.8 | **70.0** | **70.0** | 88.1 | 79.4 | 85.9 | 87.5 |
| P.Nakhon S.Ayutthaya | 77.0 | 83.3 | 79.3 | 51.7 | 51.7 | 77.1 | 74.8 | 119.1 | 77.2 |
| Pathum Thani | 94.3 | 104.6 | 102.4 | 75.4 | 75.4 | 81.6 | 95.6 | 82.3 | 122.5 |
| Pattani | 82.4 | 93.1 | 108.1 | **49.1** | **49.1** | 71.1 | 153.0 | 134.9 | 85.0 |
| Phangnga | 79.8 | 81.9 | 80.6 | **45.3** | **45.3** | 77.0 | 105.9 | 130.7 | 63.2 |
| Phatthalung | 106.9 | 132.5 | 105.8 | **55.7** | **55.7** | 74.3 | 103.2 | 159.1 | 125.5 |
| Phayao | 136.6 | 137.7 | 108.5 | **65.7** | **65.7** | 141.0 | 97.1 | 623.7 | 82.4 |
| Phetchabun | 94.7 | 106.2 | 107.3 | **58.4** | **58.4** | 117.7 | 103.1 | 358.0 | 75.5 |
| Phetchaburi | 77.7 | 86.3 | 103.0 | 54.4 | 54.4 | **52.5** | 108.3 | 59.4 | 93.3 |
| Phichit | 164.2 | 177.3 | 176.4 | 121.5 | 121.5 | **96.9** | 179.9 | 136.3 | 178.7 |
| Phitsanulok | 88.7 | 100.7 | 82.4 | **58.9** | **58.9** | 99.0 | 75.4 | 216.1 | 75.5 |
| Phrae | 211.8 | 210.8 | - | **109.9** | **109.9** | 147.8 | 171.3 | 1863.8 | 143.1 |
| Phuket | 83.7 | 102.4 | 69.8 | **54.7** | **54.7** | 56.1 | 80.4 | 94.8 | 99.0 |
| Prachin Buri | 85.7 | 93.4 | 76.3 | 72.9 | 72.9 | 97.0 | **71.6** | 184.2 | 77.3 |
| Prachuap Khiri Khan | 80.8 | 82.4 | 88.8 | 50.0 | 50.0 | **48.1** | 83.5 | 69.6 | 89.3 |
| Ranong | 62.8 | 64.4 | 63.4 | 52.0 | 52.0 | 84.7 | 63.9 | 152.4 | **48.5** |
| Ratchaburi | 57.2 | 67.2 | 65.0 | **44.2** | **44.2** | 51.5 | 68.5 | 56.6 | 66.7 |
| Rayong | 70.2 | 71.9 | 64.3 | **50.1** | **50.1** | 77.7 | 64.8 | 126.2 | 62.8 |
| Roi Et | 78.9 | 90.3 | 68.2 | 73.5 | 73.5 | 106.5 | **63.8** | 406.5 | 73.4 |
| Sa Kaeo | 86.0 | 96.1 | 80.5 | **74.5** | **74.5** | 100.9 | 75.9 | 153.5 | 84.5 |
| Sakon Nakhon | 103.1 | 110.0 | 116.4 | **68.1** | **68.1** | 132.5 | 115.7 | 528.4 | 74.6 |
| Samut Prakan | 78.8 | 81.2 | 74.1 | **62.6** | **62.6** | 65.4 | 72.9 | 101.9 | 90.8 |
| Samut Sakhon | 64.8 | 75.7 | 58.8 | **49.2** | **49.2** | 51.3 | 56.6 | 63.8 | 69.5 |
| Samut Songkhram | 114.8 | 141.4 | 104.2 | **69.5** | **69.5** | 82.7 | 96.5 | 100.3 | 135.4 |
| Saraburi | 73.7 | 82.5 | 62.1 | 60.3 | 60.3 | 87.8 | **58.5** | 154.6 | 65.3 |
| Satun | 151.7 | 229.9 | 169.4 | **86.3** | **86.3** | 89.0 | 164.8 | 213.1 | 209.7 |
| Si Sa Ket | 96.4 | 112.4 | 113.0 | **69.7** | **69.7** | 98.5 | 115.2 | 276.1 | 89.6 |
| Sing Buri | 116.1 | 93.9 | 97.2 | 120.9 | 120.9 | 124.9 | 97.1 | 977.2 | **91.8** |
| Songkhla | 78.6 | 89.6 | 84.3 | **44.4** | **44.4** | 67.8 | 97.0 | 116.9 | 84.7 |
| Sukhothai | 69.3 | 72.8 | 68.2 | **57.8** | **57.8** | 94.3 | 62.0 | 225.2 | 59.7 |
| Suphan Buri | **46.0** | 50.6 | 59.0 | 50.7 | 50.7 | 67.3 | 56.7 | 89.9 | 46.5 |
| Surat Thani | 156.1 | 193.7 | 160.6 | 85.1 | 85.1 | **70.9** | 153.4 | 115.2 | 190.0 |
| Surin | 75.8 | 89.9 | 61.2 | 67.0 | 67.0 | 103.5 | **56.9** | 333.1 | 63.6 |
| Tak | 69.8 | 81.7 | 69.1 | 52.1 | 52.1 | 109.2 | 65.3 | 397.5 | **43.7** |
| Trang | 87.7 | 95.5 | 95.5 | **45.7** | **45.7** | 88.1 | 95.5 | 214.0 | 68.9 |
| Trat | 92.5 | 99.8 | 105.2 | **61.9** | **61.9** | 86.2 | 94.9 | 175.9 | 86.0 |
| Ubon Ratchathani | 87.1 | 93.3 | 90.6 | **79.0** | **79.0** | 112.5 | 89.3 | 227.5 | 83.2 |
| Udon Thani | 99.8 | 97.1 | 99.5 | **78.6** | **78.6** | 127.2 | 97.5 | 2897.3 | 85.5 |
| Uthai Thani | 85.7 | 90.6 | 100.4 | **75.8** | **75.8** | 86.0 | 92.6 | 280.3 | 84.9 |
| Uttaradit | 70.2 | 73.4 | 70.1 | 51.4 | 51.4 | 104.4 | 68.0 | 295.2 | **51.0** |
| Yala | 70.9 | 80.1 | 73.5 | **61.9** | **61.9** | 80.1 | 76.0 | 142.0 | 79.1 |
| Yasothon | 88.5 | 102.3 | 96.7 | **74.3** | **74.3** | 104.8 | 89.1 | 202.9 | 82.4 |

### 3.7 PAE of Standard Models versus Naive Persistence Baseline

**Table 54:**
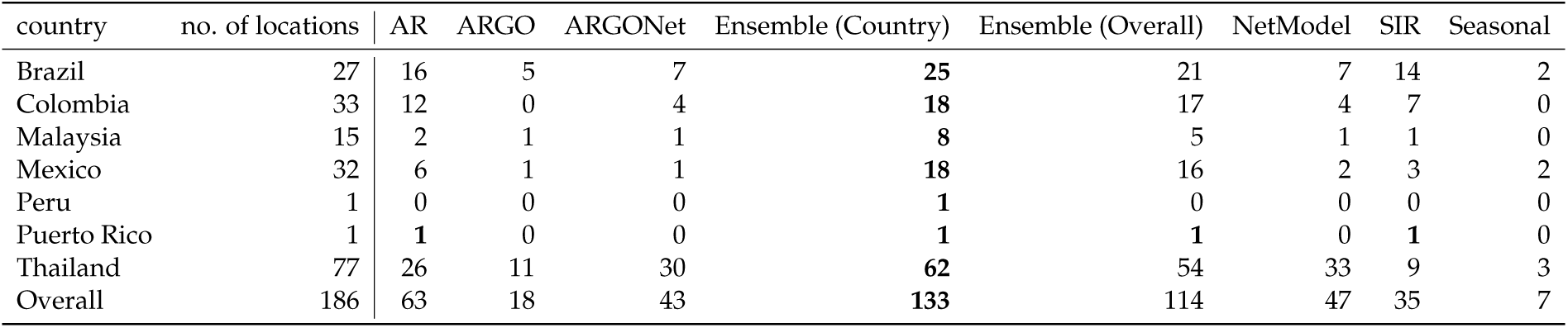
Number of locations per country where standard models outperformed the naive persistence baseline at the 1-month ahead horizon, as measured through percent absolute error. Best performing models in each location are bolded. Higher values indicate stronger performance.

| country | no. of locations | AR | ARGO | ARGONet | Ensemble (Country) | Ensemble (Overall) | NetModel | SIR | Seasonal |
| --- | --- | --- | --- | --- | --- | --- | --- | --- | --- |
| Brazil | 27 | 16 | 5 | 7 | **25** | 21 | 7 | 14 | 2 |
| Colombia | 33 | 12 | 0 | 4 | **18** | 17 | 4 | 7 | 0 |
| Malaysia | 15 | 2 | 1 | 1 | **8** | 5 | 1 | 1 | 0 |
| Mexico | 32 | 6 | 1 | 1 | **18** | 16 | 2 | 3 | 2 |
| Peru | 1 | 0 | 0 | 0 | **1** | 0 | 0 | 0 | 0 |
| Puerto Rico | 1 | **1** | 0 | 0 | **1** | **1** | 0 | **1** | 0 |
| Thailand | 77 | 26 | 11 | 30 | **62** | 54 | 33 | 9 | 3 |
| Overall | 186 | 63 | 18 | 43 | **133** | 114 | 47 | 35 | 7 |

**Table 55:** Number of locations per country where standard models outperformed the naive persistence baseline at the 2-month ahead horizon, as measured through percent absolute error. Best performing models in each location are bolded. Higher values indicate stronger performance.

| country | no. of locations | AR | ARGO | ARGONet | Ensemble (Country) | Ensemble (Overall) | NetModel | SIR | Seasonal |
| --- | --- | --- | --- | --- | --- | --- | --- | --- | --- |
| Brazil | 27 | 16 | 10 | 13 | **22** | 19 | 15 | 1 | 11 |
| Colombia | 33 | 6 | 2 | 8 | 15 | **16** | 10 | 4 | 1 |
| Malaysia | 15 | 2 | 2 | 3 | **5** | 4 | 3 | 0 | 0 |
| Mexico | 32 | 10 | 7 | 8 | **22** | **22** | 11 | 1 | 12 |
| Peru | 1 | 0 | 0 | 0 | **1** | **1** | 0 | 0 | **1** |
| Puerto Rico | 1 | **1** | 0 | 0 | **1** | **1** | 0 | **1** | 0 |
| Thailand | 77 | 36 | 20 | 41 | **69** | 65 | 44 | 4 | 27 |
| Overall | 186 | 71 | 41 | 73 | **135** | 128 | 83 | 11 | 52 |

**Table 56:** Number of locations per country where standard models outperformed the naive persistence baseline at the 3-month ahead horizon, as measured through percent absolute error. Best performing models in each location are bolded. Higher values indicate stronger performance.

| country | no. of locations | AR | ARGO | ARGONet | Ensemble (Country) | Ensemble (Overall) | NetModel | SIR | Seasonal |
| --- | --- | --- | --- | --- | --- | --- | --- | --- | --- |
| Brazil | 27 | 16 | 14 | 17 | **20** | 19 | 17 | 0 | 16 |
| Colombia | 33 | 9 | 3 | 15 | **18** | 15 | 13 | 1 | 4 |
| Malaysia | 15 | 3 | 2 | 2 | **7** | 6 | 2 | 0 | 1 |
| Mexico | 32 | 14 | 13 | 10 | 18 | **20** | 14 | 0 | 17 |
| Peru | 1 | **1** | **1** | **1** | **1** | **1** | **1** | 0 | **1** |
| Puerto Rico | 1 | **1** | **1** | **1** | **1** | **1** | **1** | 0 | **1** |
| Thailand | 77 | 46 | 40 | 43 | **71** | **71** | 47 | 2 | 52 |
| Overall | 186 | 90 | 74 | 89 | **136** | 133 | 95 | 3 | 92 |

## Data Availability

All data produced in the present study are available upon reasonable request to the authors

